# rtestim: Time-varying reproduction number estimation with trend filtering

**DOI:** 10.1101/2023.12.18.23299302

**Authors:** Jiaping Liu, Zhenglun Cai, Paul Gustafson, Daniel J. McDonald

## Abstract

To understand the transmissibility and spread of infectious diseases, epidemiologists turn to estimates of the instantaneous reproduction number. While many estimation approaches exist, their utility may be limited. Challenges of surveillance data collection, model assumptions that are unverifiable with data alone, and computationally inefficient frameworks are critical limitations for many existing approaches. We propose a discrete spline-based approach that solves a convex optimization problem—Poisson trend filtering—using the proximal Newton method. It produces a locally adaptive estimator for instantaneous reproduction number estimation with heterogeneous smoothness. Our methodology remains accurate even under some process misspecifications and is computationally efficient, even for large-scale data. The implementation is easily accessible in a lightweight `R` package `rtestim`.

## 1 Introduction

The effective reproduction number is defined to be the expected number of secondary infections produced by a primary infection where some part of the population is no longer susceptible. The effective reproduction number is a key quantity for understanding infectious disease dynamics including the potential size of an outbreak and the required stringency of control measures (Fraser, 2007; Nishiura and Chowell, 2009). The instantaneous reproduction number is a type of effective reproduction number that tracks the number of secondary infections at time *t* relative to all preceding primary infections. This contrasts with the case reproduction number at *t* which indexes a primary infection at time *t*, tracking the infectiousness of the cohort (Gostic et al., 2020). Tracking the time series of the effective reproduction number quantity is useful for understanding whether or not future infections are likely to increase or decrease from the current state (Anderson and May, 1991). Our focus is on the instantaneous reproduction number at time *t*, which we will denote ℛ (*t*). Practically, as long as ℛ (*t*) *<* 1, infections will decline gradually, eventually resulting in a disease-free equilibrium, whereas when *ℛ* (*t*) *>* 1, infections will continue to increase, resulting in endemic equilibrium. While ℛ (*t*) is fundamentally a continuous time quantity, it can be related to data only at discrete points in time *t* = 1, *…*, *n*. This sequence of instantaneous reproduction numbers over time is not observable, but, nonetheless, is easily interpretable and describes the course of an epidemic. Therefore, a number of procedures exist to estimate ℛ_*t*_ from different types of observed incidence data such as cases, deaths, or hospitalizations, while relying on various domain-specific assumptions, e.g., Goldstein et al. (2023, 2024); Hao et al. (2020); Wallinga and Teunis (2004). Importantly, accurate estimation of instantaneous reproduction numbers relies heavily on the quality of the available data, and, due to the limitations of data collection, such as underreporting and lack of standardization, estimation methodologies rely on various assumptions to compensate. Because model assumptions may not be easily verifiable from data alone, it is also critical for any estimation procedure to be robust to model misspecification.

Many existing approaches for instantaneous reproduction number estimation are Bayesian: they estimate the posterior distribution of ℛ_*t*_ conditional on the observations. One of the first such approaches is the software `EpiEstim` (Cori et al., 2020), described by Cori et al. (2013). This method is prospective, focusing on the instantaneous reproduction number, and using only observations available up to time *t* in order to estimate ℛ_*t*_ for each *i* = 1, *…*, *t*. An advantage of `EpiEstim` is its straightforward statistical model: new incidence data follows the Poisson distribution conditional on past incidence combined with the conjugate gamma prior distribution for ℛ_*t*_ with fixed hyperparameters. Additionally, the serial interval distribution, the distribution of the period between onsets of primary and secondary infections in a population, is fixed and known. For this reason, `EpiEstim` requires little domain expertise for use, and it is computationally fast. Thompson et al. (2019) modified this method to distinguish imported cases from local transmission and simultaneously estimate the serial interval distribution. Nash et al. (2023) further extended `EpiEstim` by using “reconstructed” daily incidence data to handle irregularly spaced observations.

Recently, Abbott et al. (2020) proposed a Bayesian latent variable framework, EpiNow2 (Abbott et al., 2023), which leverages incident cases, deaths or other available streams simultaneously along with allowing additional delay distributions (incubation period and onset to reporting delays) in modelling. Lison et al. (2024) proposed an extension that handles missing data by imputation followed by a truncation adjustment. These modifications are intended to increase accuracy at the most recent (but most uncertain) timepoints, to aid policymakers. Parag (2021) also proposed a Bayesian approach, `EpiFilter`, based on the (discretized) Kalman filter and smoother. `EpiFilter` also estimates the posterior of ℛ_*t*_ given using a Markov model for ℛ_*t*_ and Poisson distributed incident cases. Compared to `EpiEstim`, however, `EpiFilter` estimates ℛ_*t*_ retrospectively using all available incidence data both before and after time *t*, with the goal of being more robust in low-incidence periods. Gressani et al. (2022b) proposed a Bayesian P-splines approach, `EpiLPS`, that assumes negative binomial distributed observations, allowing for overdispersion in the observed incidence. Trevisin et al. (2023) also proposed a Bayesian model estimated with particle filtering to incorporate spatial structures. Bayesian approaches estimate the posterior distribution of the instantaneous reproduction numbers and possess the advantage that credible intervals may be easily computed. They also can incorporate prior knowledge on parameters. Another potential advantage is that a relatively large prior on the mean of ℛ_*t*_ can be used to guard against erroneously concluding that an epidemic is shrinking (Thompson et al., 2019). However, a downside is that the induced bias can persist for long periods of time. Bayesian approaches that do not use conjugate priors, or that incorporate multilevel modelling, can be computationally expensive, especially when observed data sequences are long or hierarchical structures are complex, e.g., Abbott et al. (2020).

There are also frequentist approaches for ℛ_*t*_ estimation. Abry et al. (2020) proposed regularizing the smoothness of ℛ_*t*_ through penalized regression with second-order temporal regularization, additional spatial penalties, and with Poisson loss. Pascal et al. (2022) extended this procedure by adding a penalty on outliers. Pircalabelu (2023) proposed a spline-based model relying on the assumption of exponential-family distributed incidence. Ho et al. (2023) estimated ℛ_*t*_ while monitoring the time-varying level of overdispersion. There are other spline-based approaches such as Azmon et al. (2014); Gressani et al. (2022a), autoregressive models with random effects (Jin et al., 2023) that are robust to low incidence, and generalized autoregressive moving average models (Hettinger et al., 2023) that are robust to measurement errors in incidence data.

We propose an instantaneous reproduction number estimator, that requires only incidence data. Our model makes the conditional Poisson assumption, similar to much of the prior work described above, but is empirically more robust to misspecification. This estimator is defined by a convex optimization problem with Poisson loss and *𝓁*_1_ penalty on the temporal evolution of log(ℛ_*t*_) to impose smoothness over time. As a result, it generates discrete splines, and the estimated curves (on the logarithmic scale) appear to be piecewise polynomials of an order selected by the user. Importantly, the estimates are locally adaptive, meaning that different time ranges may possess heterogeneous smoothness. Because we penalize the logarithm of ℛ_*t*_, we naturally accommodate the positivity requirement, in contrast to related methods (Abry et al., 2020; Pascal et al., 2022), can handle large or small incidence measurements, and are automatically (reasonably) robust to outliers without additional constraints (a feature of the *𝓁*_1_ penalty). A small illustration using three years of Covid-19 case data in Canada (Berry et al., 2021) is shown in Figure 1, where we use a time-varying serial interval distribution. The implementation is easily accessible in a lightweight `R` called `rtestim`.

**Figure 1:**
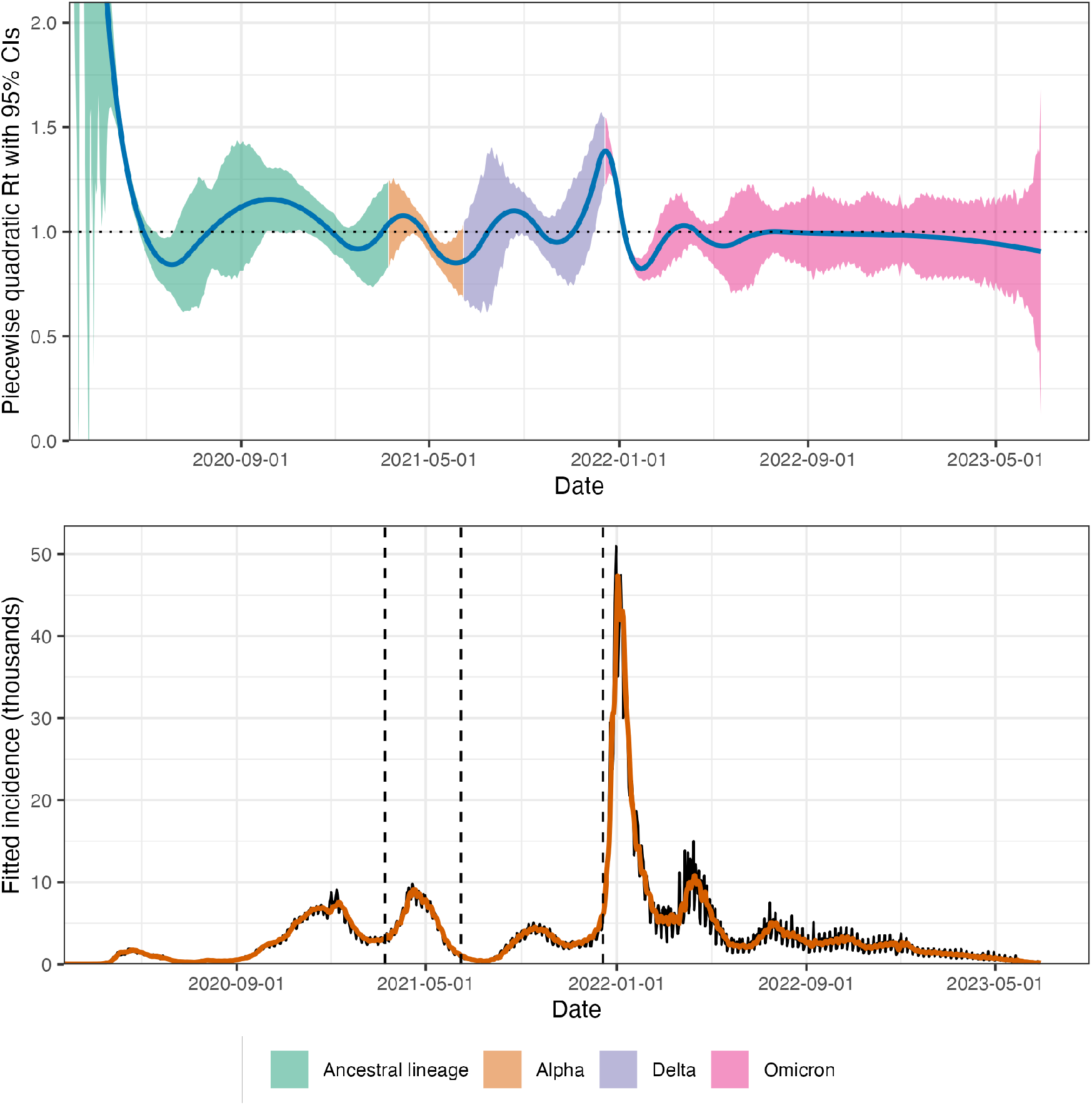
A demonstration of instantaneous reproduction number estimation by rtestim and the corresponding predicted incident cases. The example is the Covid-19 epidemic in Canada during the period from January 23, 2020 to June 28, 2023. In the top panel, the blue curve is the estimated piecewise quadratic ℛ_*t*_ and the colorful ribbon is the corresponding 95% confidence band. The colors represent the variants whose serial interval distributions are used to estimate ℛ_*t*_. The dominant circulating variants are based on a multinomial logistic regression model with variant probabilities from CAMEO Group (2023). The time-varying serial interval distributions are based on results from Xu et al. (2023). In the bottom panel, the black curve is the observed Covid-19 daily confirmed cases, and the orange curve on top of it is the predicted incident cases corresponding to the estimated ℛ_*t*_. The three vertical dashed lines represent the beginning of a new dominant variant.

While our approach is straightforward and requires little domain knowledge for implementation, we also implement a number of refinements:

- the algorithm solves over a range of tuning parameters simultaneously, using warm starts to speed up subsequent solutions;
- cross-validation is built in (and used in all analyses below) to automatically select tuning parameters;
- parametric (gamma), non-parametric (any discretized delay), and time-varying delay distributions are allowed;
- irregularly spaced incidence data are easily accommodated;
- approximate confidence intervals for ℛ_*t*_ and the observed incidence are available;
- the estimated log ℛ_*t*_ can be mathematically described as an element of a well-known function space depending on user choice (Tibshirani, 2022).

We use a proximal Newton method to solve the convex optimization problem along with warm starts to produce estimates efficiently, typically in a matter of seconds, even for long sequences of data. In a number of simulation experiments, we show empirically that our approach is more accurate than existing methods at estimating the true instantaneous reproduction numbers and robust to some degrees of misspecification of incidence distribution, serial interval distribution, and the order of graphical curvature.

The manuscript proceeds as follows. We first introduce our ℛ_*t*_ estimation methodology including the renewal equation and the development of Poisson trend filtering estimator. We explain how this method could be interpreted from the Bayesian perspective, connecting it to previous work in this context. We provide illustrative experiments comparing our estimator to other Bayesian alternatives. We then apply our methodology to the Covid-19 pandemic in Canada and the 1918 influenza pandemic in the United States. Finally, we conclude with a discussion of the advantages and limitations of our approach and describe some practical considerations for instantaneous reproduction number estimation.

## 2 Methods

### 2.1 Renewal model for incidence data

The instantaneous reproduction number ℛ (*t*) is defined to be the expected number of secondary infections at time *t* produced by a primary infection sometime in the past. To make this precise, denote the number of new infections at time *t* as *y*(*t*). Then the total primary infectiousness can be written as 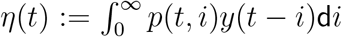, where *p*(*t, i*) is the probability that a new secondary infection at time *t* is the result of a primary infection that occurred *i* time units in the past. The instantaneous reproduction number is then given as the value that equates

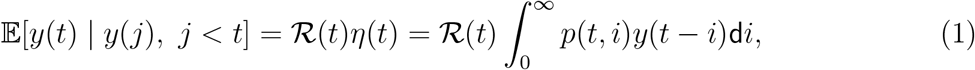

otherwise known as the renewal equation. The period between primary and secondary infections is exactly the generation time of the disease, but given real data, observed at discrete times (say, daily), this delay distribution must be discretized into contiguous time intervals, say, (0, 1], (1, 2], *…*, resulting in the sequence 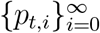 corresponding to observations *y*_*t*_ for each *t* and yields the discretized version of Equation (1),

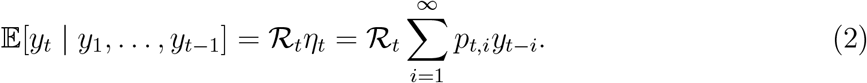

Many approaches to estimating ℛ_*t*_ rely on Equation (2) as motivation for their procedures, among them, `EpiEstim` (Cori et al., 2013) and `EpiFilter` (Parag, 2021).

In most cases, it is safe to assume that infectiousness disappears beyond *τ* timepoints (*p*(*t, i*) = 0 for *i > τ*), resulting in the truncated integral of the generation interval distribution 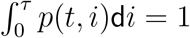 or each *t*. Generation time, however, is usually unobservable and tricky to estimate, so common practice is to approximate it by the serial interval: the period between the symptom onsets of primary and secondary infections. If the infectiousness profile after symptom onset is independent of the incubation period (the period from the time of infection to the time of symptom onset), then this approximation is justifiable: the serial interval distribution and the generation interval distribution share the same mean. However, other properties may not be similarly shared, and, in general, the generation interval distribution is a convolution of the serial interval distribution with the distribution of the difference between independent draws from the delay distribution from infection to symptom onset. See, for example, Gostic et al. (2020) for a fuller discussion of the dangers of this approximation. Nonetheless, treating these as interchangeable is common (Cori et al., 2013; Park et al., 2021), and doing otherwise is beyond the scope of this work. We will allow the delay distribution to be either constant over time—the probability *p*(*i*) depends only on the gap between primary and secondary infections and not on the time *t* when the secondary infection occurs—or to be time-varying: *p*(*t, i*) also depends on the time of the secondary infection. For our methods, we assume that the serial interval can be accurately estimated from auxiliary data (say by contact tracing, or previous epidemics) and we take it as fixed, as is common in existing studies (Abry et al., 2020; Cori et al., 2013; Pascal et al., 2022).

The renewal equation in Equation (2) relates observable data streams (incident cases) occurring at different timepoints to the instantaneous reproduction number given the serial interval. The fact that it depends only on the observed incident counts makes it reasonable to estimate ℛ_*t*_. However, data collection idiosyncrasies can obscure this relationship. Diagnostic testing targets symptomatic individuals, omitting asymptomatic primary infections which can lead to future secondary infections. Testing practices, availability, and uptake can vary across space and time (Hitchings et al., 2021; Pitzer et al., 2021). Finally, incident cases as reported to public health are subject to delays due to laboratory confirmation, test turnaround times, and eventual submission to public health (Pellis et al., 2021). For these reasons, reported cases are lagging indicators of the course of the pandemic. Furthermore, they do not represent the actual number of new infections that occur on a given day, as indicated by exposure to the pathogen. The assumptions described above (homogeneous mixing, similar susceptibility and social behaviours, etc.) are therefore consequential. That said, Equation (2) also provides some comfort about deviations from these assumptions. Under certain conditions, failing to account for the reporting behaviours will minimally impact the accuracy of any ℛ_*t*_ estimator that is based on Equation (2). We discuss three types of deviation here. First, if *y*_*t*_ is scaled by a constant *a* describing the reporting ratio, then, because it appears on both sides of Equation (2), ℛ_*t*_ will be unchanged. Second, if such a scaling *a*_*t*_ varies in time, as long as it varies slowly relative to *p*_*i*_ —that is, if 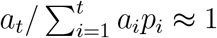 —then ℛ_*t*_ can still be estimated well from reported incidence data. Finally, if a sudden change in reporting ratio occurs at time *t*_1_, it would only result in large errors in ℛ_*t*_ at times near *t*_1_ (where the size of this neighbourhood is determined indirectly by the effective support of 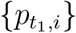. On the other hand, time-varying reporting delays would be much more detrimental (Eales and Riley, 2024; Park et al., 2024).

### 2.2 Poisson trend filtering estimator

We use the daily confirmed incident cases *y*_*t*_ on day *t* to estimate the observed infectious cases under the model that *y*_*t*_, given previous incident cases *y*_*t−*1_, *…*, *y*_1_ and a constant serial interval distribution, follows a Poisson distribution with mean Λ_*t*_. That is,

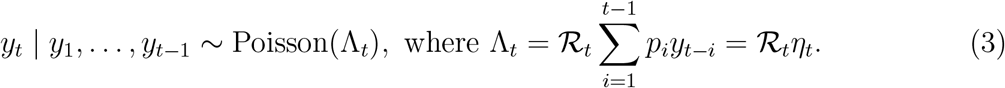

We will write *p*_*i*_ as constant in time for simplicity, although this is not required. Given a history of *n* confirmed incident counts **y** = (*y*_1_, *…*, *y*_*n*_)^T^, our goal is to estimate *ℛ*_*t*_ for each *t* = 1, *…*, *n*. A natural approach is to maximize the likelihood, producing the maximum likelihood estimator (MLE):

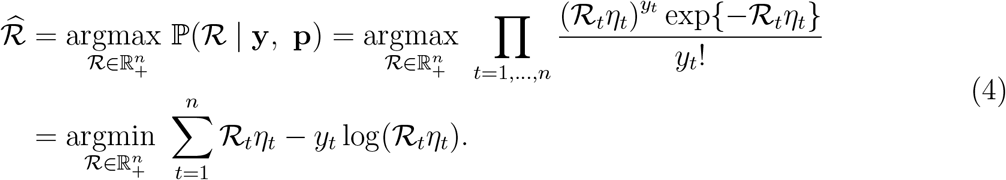

This optimization problem, however, is easily seen to yield a one-to-one correspondence between the observation and the estimated instantaneous reproduction number, i.e., 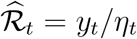, so that the estimated sequence 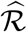 will have no significant smoothness.

The MLE is an unbiased estimator of the true parameter ℛ_*t*_, but unfortunately has high variance: changes in *y*_*t*_ result in proportional changes in 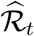. To avoid this behaviour, and to match the intuition that ℛ_*t*_ ≈ *ℛ*_*t−*1_, we advocate enforcing smoothness of the instantaneous reproduction numbers. This constraint will decrease the estimation variance, and hopefully lead to more accurate estimation of ℛ, as long as the smoothness assumption is reasonable. Smoothness assumptions are common (see e.g., Gostic et al. (2020); Parag (2021)), but the type of smoothness assumption is critical. Cori et al. (2013) imposes smoothness indirectly by estimating ℛ_*t*_ with moving windows of past observations. The Kalman filter procedure of Parag (2021) would enforce *𝓁*_2_ -smoothness 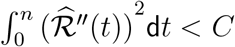 for some constant *C*), although the computational implementation results in 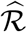 taking values over a discrete grid. Pascal et al. (2022) produces piecewise linear 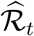, which turns out to be closely related to a special case of our methodology. Smoother estimated curves will provide high-level information about the entire epidemic, obscuring small local changes in ℛ (*t*), but may also remove the ability to detect large sudden changes, such as those resulting from lockdowns or other major containment policies.

To enforce smoothness of 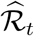, we add a trend filtering penalty (Kim et al., 2009; Sadhanala et al., 2024; Tibshirani, 2014, 2022) to Equation (5). Because ℛ_*t*_ *>* 0, we explicitly penalize the divided differences (discrete derivatives) of neighbouring values of log(ℛ_*t*_). Let *θ* := log(ℛ) ∈ *ℝ*^*n*^, so that Λ_*t*_ = *η*_*t*_ exp(*θ*_*t*_), and log(*η*_*t*_ ℛ_*t*_) = log(*η*_*t*_)+*θ*_*t*_. For evenly spaced incidence data, we write our estimator as the solution to the optimization problem

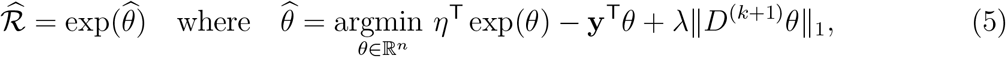

where exp(*·*) applies elementwise and 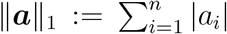 is the *𝓁*_1_ norm. Here, *D*^(*k*+1)^ ∈ ℤ ^(*n−k−*1)*×n*^ is the (*k* + 1)^th^ order divided difference matrix for any *k* ∈ *{*0, *…*, *n −* 1*}* with the convention that *D*^(0)^ = **0**_*n×n*_. The divided difference matrix for *k* = 0, *D*^(1)^ ∈ *{−*1, 0, 1*}*^(*n−*1)*×n*^, is a sparse matrix with diagonal band of the form:

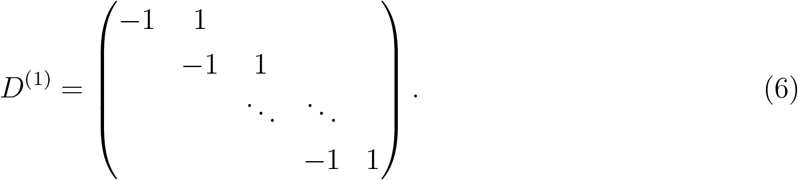

For *k* ≥ 1, *D*^(*k*+1)^ can be defined recursively as *D*^(*k*+1)^ := *D*^(1)^*D*^(*k*)^, where *D*^(1)^ ∈ *{−*1, 0, 1*}*^(*n−k−*1)*×*(*n−k*)^ has the form Equation (6) but with modified dimensions.

The tuning parameter (hyperparameter) *λ* balances data fidelity with desired smoothness. When *λ* = 0, the problem in Equation (5) reduces to the MLE in Equation (4). Larger tuning parameters privilege the regularization term and yield smoother estimates. Finally, there exists *λ*_max_ such that any *λ* ≥ *λ*_max_ will result in 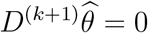 and 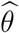 will be the Kullback-Leibler projection of **y** onto the null space of *D*^(*k*+1)^ (see Section 2.3 for more details).

The solution to Equation (5) will result in piecewise polynomials, specifically called discrete splines. For example, 0^th^-degree discrete splines are piecewise constant, 1^st^-degree curves are piecewise linear, and 2^nd^-degree curves are piecewise quadratic. For *k* ≥ 1, *k*^th^-degree discrete splines are continuous and have continuous discrete differences up to degree *k −* 1 at the knots (i.e., changing points between segments). This penalty results in more flexibility compared to the homogeneous smoothness that is created by the squared *𝓁*_2_ norm. Using different orders of the divided differences results in estimated instantaneous reproduction numbers with different smoothness properties.

For unevenly spaced data, the spacing between neighbouring parameters varies with the time between observations, and thus, the divided differences must be adjusted by the times that the observations occur. Given observation times **x** = (*x*_1_, *…*, *x*_*n*_)^T^, for *k* ≥ 1, define a *k*^th^-order diagonal matrix

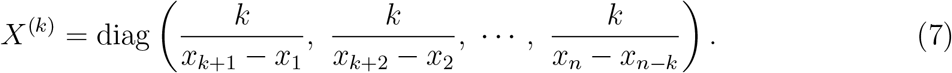

Letting *D*^(**x**,1)^ := *D*^(1)^, then for *k* ≥ 1, the (*k* + 1)^th^-order divided difference matrix for unevenly spaced data can be created recursively by *D*^(**x**,*k*+1)^ := *D*^(1)^*X*^(*k*)^*D*^(**x**,*k*)^. No adjustment is required for *k* = 0.

Due to the penalty structure, this estimator is locally adaptive, meaning that it can potentially capture local changes such as the initiation of control measures, becoming more wiggly in regions that require it. In contrast, Abry et al. and Pascal et al. considered only the 2^nd^-order (*k* = 1) divided difference of *ℛ*_*t*_ rather than its logarithm (Abry et al., 2020; Pascal et al., 2022). In comparison to their work, our estimator (i) allows for arbitrary degrees of temporal smoothness and (ii) avoids the potential numerical issues of penalizing/estimating positive real values. Nonetheless, as we will describe below, our procedure is computationally efficient for estimation over an entire sequence of hyperparameters *λ* and provides methods for choosing how smooth the final estimate should be.

### 2.3 Solving over a sequence of tuning parameters

We can solve the Poisson trend filtering estimator over an arbitrary sequence of *λ* that produces different levels of smoothness in the estimated curves. We consider a candidate set of M *λ*-values, 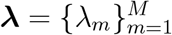, that is strictly decreasing.

Let *D* := *D*^(*k*+1)^ for simplicity in the remainder of this section. As *λ* → ∞, the penalty term *λ*∥*Dθ*∥_1_ dominates the Poisson loss, so that minimizing Equation (5) is asymptotically equivalent to minimizing the penalty term, which results in ∥*Dθ*∥_1_ = 0. In this case, the divided differences of *θ* with order *k* + 1 is always 0, and thus, *θ* must lie in the null space of *D*, that is, *θ* ∈ *𝒩* (*D*). The same happens for any *λ* beyond this threshold, so define *λ*_max_ to be the smallest *λ* that produces *θ* ∈ *𝒩* (*D*). It turns out that this value can be written explicitly as *λ*_max_ = ∥(*D*^*†*^)^T^ (*η − y*)∥_∞_, where *D*^*†*^ is the (left) generalized inverse of *D* satisfying *D*^*†*^*D* = *I* and ∥*a*∥_∞_ := max_*i*_|*a*_*i*_| is the infinity norm. Explicitly, for any *λ* ≥ *λ*_max_, the solution to Equation (5) will be identical to the solution with *λ*_max_. Therefore, we use *λ*_1_ = *λ*_max_ and choose the minimum *λ*_*M*_ to be *rλ*_max_ for some *r* ∈ (0, 1) (typically *r* = 10^*−*4^). Given any *M* ≥ 3, we generate a sequence of *λ* values to be equally spaced on the log-scale between *λ*_1_ and *λ*_*M*_ .

To compute the sequence of solutions efficiently, the model is estimated sequentially by visiting each *λ*_*m*_ in order, from largest to smallest. The estimates produced for a larger *λ* are used as the initial values (warm starts) for the next smaller *λ*. By solving through the entire sequence of tuning parameters, we improve computational efficiency and also enable one to trade between bias and variance, resulting in improved accuracy relative to procedures using a single fixed tuning parameter.

### 2.4 Choosing a final *λ*

We estimate model accuracy over the candidate set through *V* -fold cross validation (CV) to choose the best tuning parameter. Specifically, we divide **y** (except the first and last observations) roughly evenly and randomly into *V* folds, estimate *ℛ*_*t*_ for all ***λ*** leaving one fold out, and then predict the held-out observations. Alternativly, one could use regular splitting, assigning every *v*^th^ observation into the same fold. Note that our approach is most closely related to non-parametric regression rather than time series forecasting. That said, under some conditions, one can guarantee that *V* -fold remains valid for risk estimation in time series. The sufficient conditions are quite strong, but the guarantees are also stronger than would be required for model selection consistency (Bergmeir et al., 2018).

Model accuracy can be measured by multiple metrics such as mean squared error 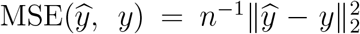 or mean absolute error 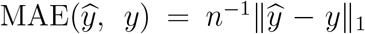, but we prefer to use the (average) deviance, to mimic the likelihood in Equation (4): 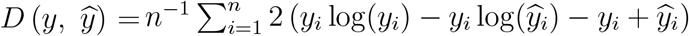, with the convention that 0 log(0) = 0. Note that for any *V* and any *M*, we will end up estimating the model (*V* + 1)*M* times rather than once.

### 2.5 Approximate confidence bands

We also provide empirical confidence bands of the estimators with approximate coverage. Consider the related estimator 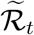 defined as

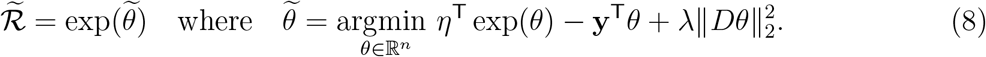

Letting 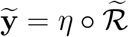 (where ∘ denotes the elementwise product), it can be shown (for example, Theorem 2 in Vaiter et al. (2017)) that an estimator for 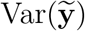 is given by 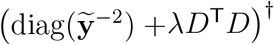. Finally, an application of the delta method shows that 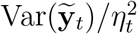 is an estimator for 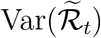 for each *t* = 1, *…*, *n*. We therefore use 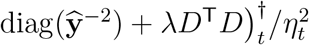 as an estimator for 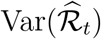. An approximate (1 *− α*)% confidence interval then can be written as 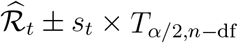, where *s*_*t*_ is the square-root of 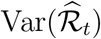 for each *t* = 1, *…*, *n* and df is the number of changepoints in 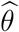 plus *k* + 1 (Tibshirani, 2014). An approximate confidence interval of 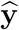 can be computed similarly.

### 2.6 Bayesian perspective

Unlike many other methods for ℛ_*t*_ estimation, our approach is frequentist rather than Bayesian. Nonetheless, it has a corresponding Bayesian interpretation: as a state-space model with Poisson observational noise, autoregressive transition equation of degree *k* ≥ 0, e.g., *θ*_*t*+1_ = 2*θ*_*t*_ *− θ*_*t−*1_ + *ε*_*t*+1_ for *k* = 1, and Laplace transition noise *ε*_*t*+1_ ∼ Laplace(0, 1*/λ*). Compared to `EpiFilter` (Parag, 2021), we share the same observational assumptions, but our approach has a different transition noise. `EpiFilter` estimates the posterior distribution of *ℛ*_*t*_, and thus it can provide credible interval estimates as well. Our approach produces the maximum *a posteriori* estimate via an efficient convex optimization, obviating the need for MCMC sampling. But the associated confidence bands are created differently.

## 3 Results

Implementation of our approach is provided in the `R` package `rtestim`. All computational experiments are conducted on the Cedar cluster provided by the Digital Research Alliance of Canada with `R` 4.3.1. The `R` packages used for simulation and real-data application are `EpiEstim` 2.2-4 (Cori et al., 2022), `EpiLPS` 1.2.0 (Gressani, 2021), and `rtestim` 0.0.4. The `R` scripts for `EpiFilter` are used (Parag, 2020).

### 3.1 Synthetic experiments

#### 3.1.1 Design for the synthetic data

We simulate four scenarios of the instantaneous reproduction number, intended to mimic different epidemics. The first two scenarios are rapidly controlled by intervention, where the ℛ (*t*) consists of one discontinuity and two segments. Scenario 1 has constant ℛ (*t*) before and after an intervention, while Scenario 2 grows exponentially, then decays. The other two scenarios are more complicated, where more waves are involved. Scenario 3 has four linear segments with three discontinuities, which reflect the effect of an intervention, resurgence to rapid transmission, and finally suppression of the epidemic. Scenario 4 involves sinusoidal waves throughout the epidemic. The first three scenarios and the last scenario are motivated by Parag (2021) and Gressani et al. (2022b) respectively. We name the four scenarios as (1) piecewise constant, (2) piecewise exponential, (3) piecewise linear, and (4) periodic.

In all cases, the times of observation are regular, and epidemics are of length *n* = 300. Specifically, in Scenario 1, ℛ_*t*_ = 2 for *t* ≤ 120 and 0.8 for *t >* 120. In Scenario 2, *ℛ*_*t*_ increases and decreases exponentially with rates 0.01 for *t* ≤ 100 and 0.005 for *t >* 100. In Scenario 3, *ℛ*_*t*_ is piecewise linear with four discontinuous segments,

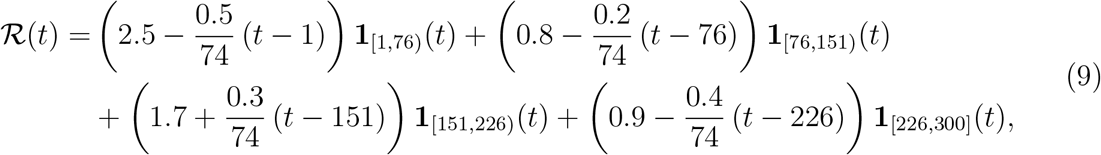

where **1**_*A*_(*t*) = 1, if *t* ∈ *A*, and **1**_*A*_(*t*) = 0 otherwise. In Scenario 4, ℛ_*t*_ is realization of the continuous, periodic curve generated by the function

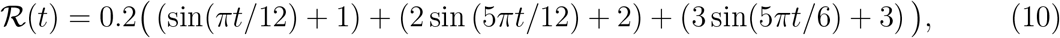

evaluated at equally spaced points *t* ∈ [0, 10]. These ℛ_*t*_ scenarios are illustrated in Figure 2. We compute the expected incidence Λ_*t*_ using the renewal equation, and generate the incident infections from the Poisson distribution with mean 𝔼 [*y*_*t*_ | *y*_*s*_, *s < t*] = Λ_*t*_. To verify the performance of our model under violations of the model’s distributional assumptions, we also generate incident cases using the negative binomial distribution with dispersion parameter *ρ* = 5. Here, the negative binomial is parameterized such that the mean is 𝔼 [*y*_*t*_ | *y*_*s*_, *s < t*] = Λ_*t*_ and the variance is Var[*y*_*t*_ | *y*_*s*_, *s < t*] = Λ_*t*_(1 + Λ_*t*_*/ρ*) (following, for example, Gressani et al. (2022b)). Because (1+Λ_*t*_*/ρ*) *>* 1 for 0 ≤ *ρ <* ∞, this parameterization results in overdispersion relative to the Poisson distribution, with smaller values of *ρ* leading to greater overdispersion. For context on the observed dispersion of these synthetic experiments, Figure A.2.1 in the Supplement displays the ratio of the time-varying standard deviation to the mean.

**Figure 2:**
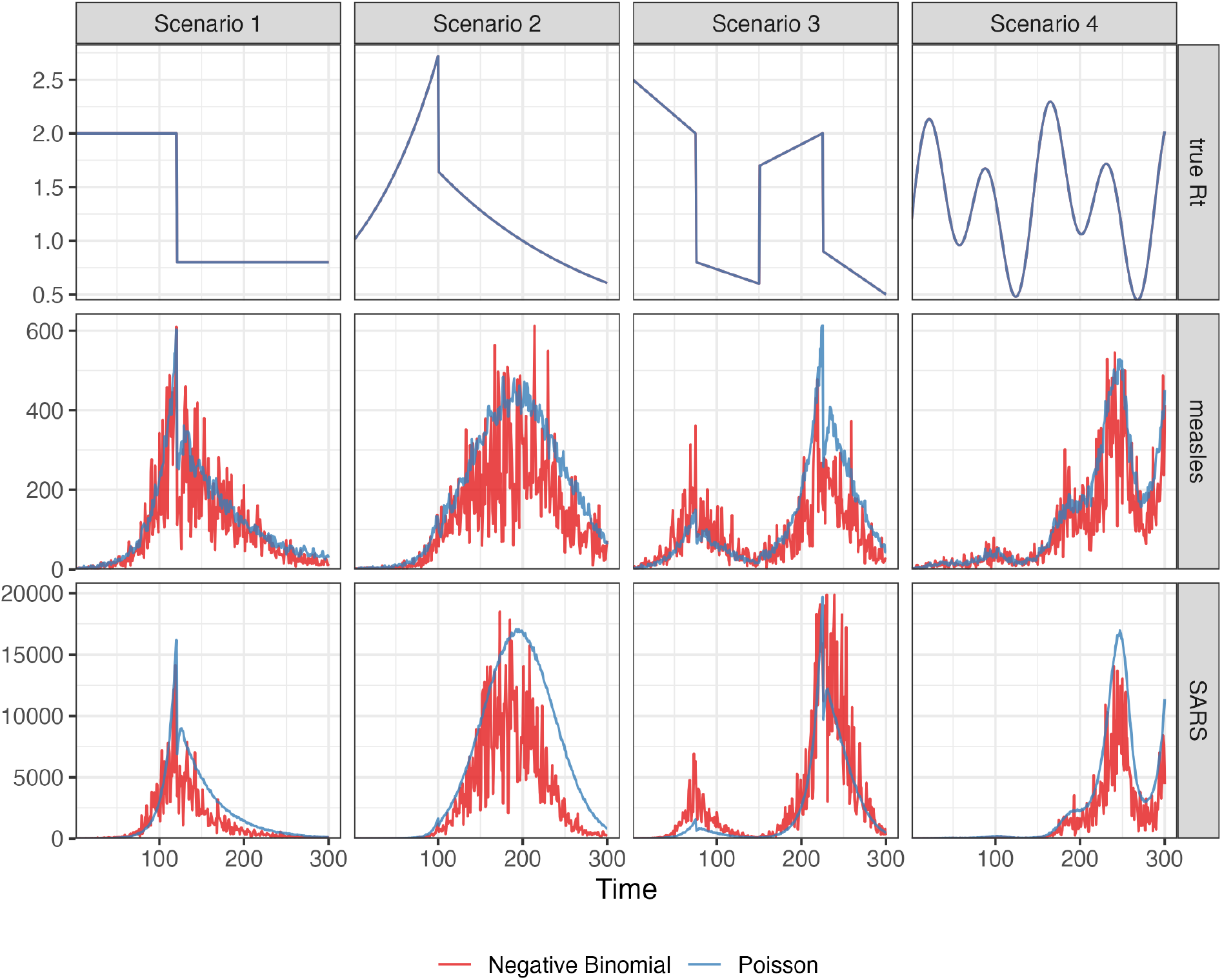
This figure displays example realizations for each ℛ_*t*_ setting. Top row: the instantaneous reproduction numbers. Middle row: synthetic measles incidence (Poisson in blue, negative binomial in red) incidence. Bottom row: synthetic SARS incidence. The corresponding ℛ_*t*_ scenarios are shown in the columns.

We use serial interval (SI) distributions of measles (with mean 14.9 and standard deviation 3.9) in Hagelloch, Germany in 1861 (Groendyke et al., 2011) and SARS (with mean 8.4 and standard deviation 3.8) in Hong Kong in 2003 (Lipsitch et al., 2003), inspired by Cori et al. (2013), to generate synthetic epidemics. We initialize all epidemics with *y*_1_ = 2 cases and generate for *t* = 2, *…*, 300. The synthetic measles epidemics have smaller incident cases in general, and the SARS epidemics have larger incidence. Essentially, the smaller mean of the serial interval for SARS with a similar standard deviation leads to shorter expected delays between onsets of primary and secondary infections, resulting in faster growth of incidence within the same period of time. We also consider shorter flu epidemics with 50 timepoints with piecewise linear ℛ_*t*_ (Scenario 3) considering both incidence distributional assumptions. The motivation is to compare our method and other alternatives with EpiNow2 which takes much longer to compute for long epidemics (nearly 2 hours to converge for a measles epidemic with 300 timepoints) than other methods. Besides using the correct SI distributions to estimate ℛ_*t*_, we also consider the scenarios where the SI is misspecified. More details on experimental settings and results for shorter epidemics and misspecification of SI distributions are given in Sections A.2.1 and A.3 in the supplementary document respectively.

For each problem setting (including SI distribution, an ℛ_*t*_ scenario, and an incidence distribution), we generate 50 random samples, resulting in 800 total synthetic epidemics. Example realizations for measles and SARS with each instantaneous reproduction number scenario is displayed in Figure 2.

#### 3.1.2 Algorithmic choices

We compare `rtestim` to `EpiEstim`, `EpiLPS`, and `EpiFilter`. `EpiEstim` estimates the posterior distribution of the instantaneous reproduction number given a Gamma prior and Poisson distributed observations over a trailing window, under the assumption that the instantaneous reproduction number is constant during that window. A larger window averages out more fluctuations, leading to smoother estimates, whereas a shorter window is more responsive to sudden spikes or declines. We used weekly sliding and monthly windows, however, since neither considerably outperforms the other across all scenarios, we defer the monthly results to the supplementary document. `EpiLPS` is another Bayesian approach that estimates P-splines based on the Laplace approximation to the conditional posterior with negative binomial likelihood. It should more easily handle the negative binomial scenarios as it matches the data generating process. `EpiFilter` uses a particle filtering procedure on a discrete grid of possible ℛ_*t*_ values.

In each setting, we apply `rtestim` with four choices of *k* = 0, 1, 2, 3 resulting in different shapes of the estimated ℛ_*t*_—piecewise constant, piecewise linear, piecewise quadratic, and piecewise cubic—respectively. We use 10-fold cross validation (CV) to choose the parameter *λ* that minimizes out-of-sample prediction risk from a candidate set of size 50, i.e., ***λ*** = *{λ*_1_, *…*, *λ*_50_*}*, for long epidemics, and 5-fold CV for short epidemics (results for this case are deferred to Sections A.3.2 and A.4.2 in the Supplement). We select the tuning parameter that gives the lowest deviance between the estimated incidence and the held-out samples averaged over all folds.

For the alternative methods, we generally use the set of tuning parameters that were applied to their own experimental settings. We consider both weekly and monthly sliding windows in `EpiEstim`. `EpiLPS` uses 40 *P* -spline basis functions and optimizes using the Nelder-Mead procedure. For `EpiFilter`, we specify a grid with 2000 cells, use 0.1 for the size of the diffusion noise, and use the “smoothed” ℛ_*t*_ (conditional on all data) as the final estimate.

For the ℛ_*t*_ estimation using all models for each problem, we use the same serial interval distribution, that was used to generate synthetic data. Taking different hyperparameters into consideration, we solve each problem using 8 methods including `EpiEstim` with weekly or monthly sliding windows, `EpiLPS`, `EpiFilter`, and `rtestim` with piecewise constant, linear, quadratic, or cubic curves. We have not made any effort to tune these (and other choices) more carefully.

For `rtestim`, the choice of *k* explicitly controls the function space to which the solution will belong (Tibshirani, 2022), providing the analyst with a mathematical understanding of the result. When faced with real data, the choice of *k* for `rtestim` should be done either (1) based on the analyst’s preference for the resulting structure (e.g., “I want to find large jumps, so *k* = 0”) or (2) in a data-driven manner, as a component of the estimation process. Our software enables both cases: the second case can be implemented by simply fitting different *k* and choosing the set *k, λ* that has smallest CV score. Thus, all necessary choices can be accomplished based solely on the data, a departure from existing methods in that we both allow this choice and provide simple data-driven methods to accomplish it.

#### 3.1.3 ccuracy measurement

To measure estimation accuracy, we compare the estimated 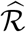 to the true ℛ using the Kullback-Leibler (KL) divergence. KL is useful in this context for a few reasons. First, it correctly handles the non-negativity constraint on ℛ. Second, KL matches the negative log-likelihood used in Equation (4). Third, it captures the curved geometry of the probability spaces implied by the Poisson distribution accurately. And fourth, as in the equation below, it has a convenient functional form depending only on ℛ and *η*. For the Poisson distribution the KL divergence is defined as

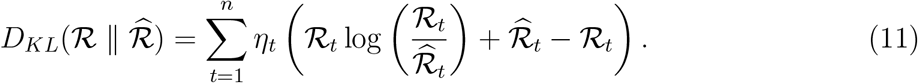

We use the average KL divergence: 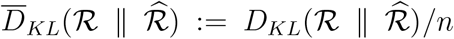. Details on the derivation of Equation (11) is provided in Section A.1 of the Supplement. KL divergence is more appropriate for measuring accuracy because it connects directly to the Poisson likelihood used to generate the data, whereas standard measures like the mean-squared error correspond to Gaussian likelihood. Using Poisson likelihood has the effect of increasing the relative cost of mistakes when Λ_*t*_ is small.

To fairly compare across methods, we omit the first week of data (and estimates) for a few reasons. Estimates from `EpiEstim` are not available until *t* = 8 when using a weekly sliding window. Additionally, some procedures purposely impose strong priors that ℛ_1_ is much larger than 1 to avoid over confidently asserting that an epidemic is under control. The effect of these priors will persist for days or weeks, but one would hope for accurate estimates as early in the outbreak as possible. Other details of the experimental settings are deferred to the supplementary document.

### 3.2 Results for synthetic data

In general, `rtestim` performs at least as well as the other competitors in the experimental study. Figure 3 and Figure 4 visualize the KL divergence across the seven methods. For low incidence in measles epidemics, `rtestim` is the most accurate for all ℛ_*t*_ scenarios given both Poisson and negative binomial incidence. The best performance of `rtestim` has the lowest median and has little or no overlap with other methods. For Scenario 1 with Poisson incidence, `EpiFilter` has a similar median to that of `rtestim` and small variability, making it a competitive alternative. However for negative binomial incidence, `EpiFilter` loses its advantage and has the largest medians of any method in Scenarios 1 and 2. The large incidence in SARS epidemics is more difficult for all methods. For Poisson incidence, results are similar to the previous setting. However, for negative binomial incidence, `EpiLPS` performs at least as well if not better than `rtestim`, especially in Scenarios 2 and 4. Nonetheless, `rtestim` is largely similar, with simulation uncertainty suggesting comparable performance. We will examine a single realization of each experiment to investigate these global conclusions in more detail.

**Figure 3:**
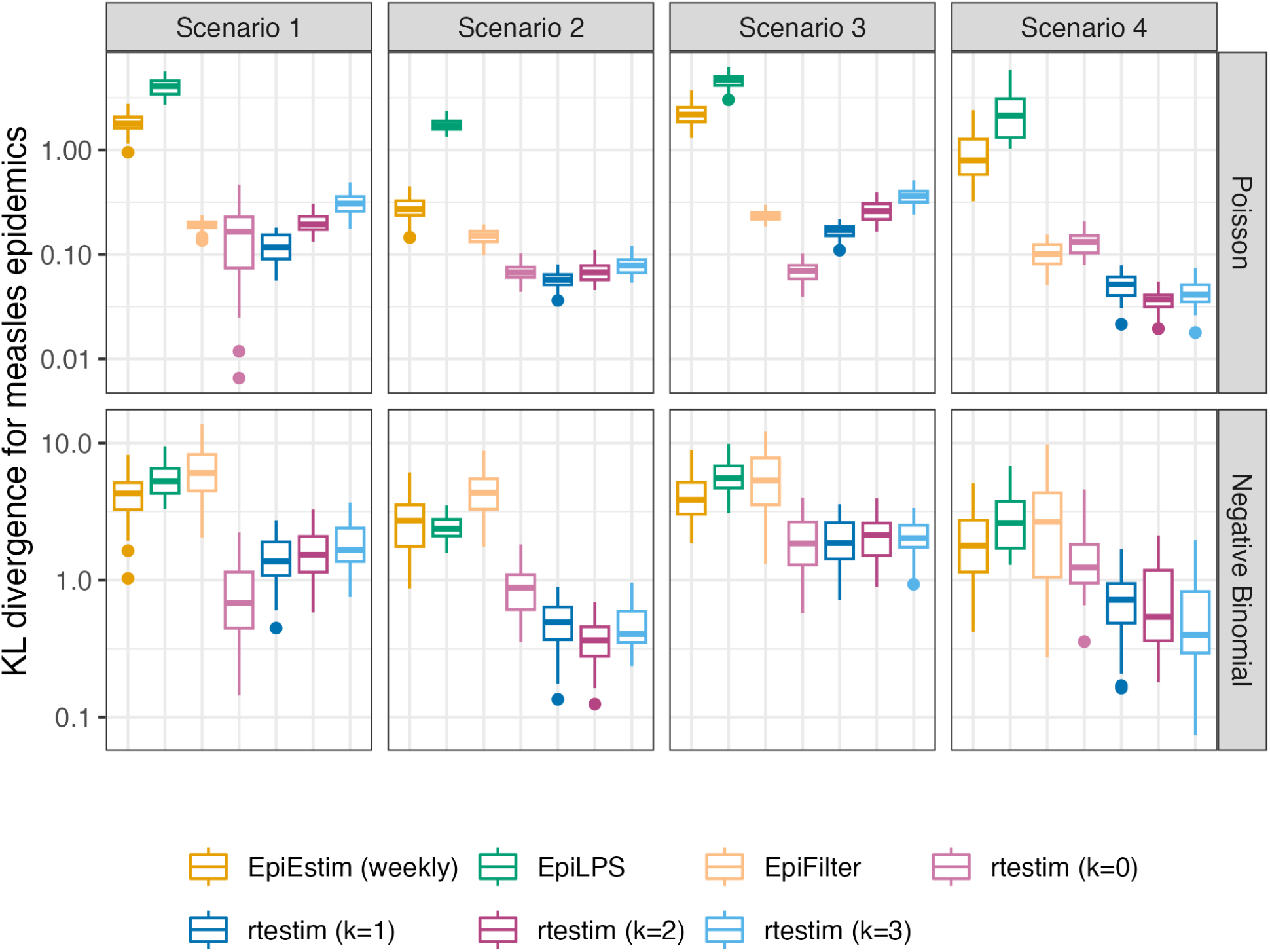
Boxplot of mean KL divergence between 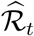 and true ℛ_*t*_ across 50 synthetic measles epidemics. Performance of each approach given Poisson incidence is in top panels and negative binomial incidence is in bottom panels. The average excludes the first week in all settings, since `EpiEstim` with a weekly sliding window does not provide estimates for the first week. Outliers beyond 1.5*×*IQR of each box are excluded for the sake of comparison with full range of the *y*-axis deferred to Figure A.3.1 in the Supplement.

**Figure 4:**
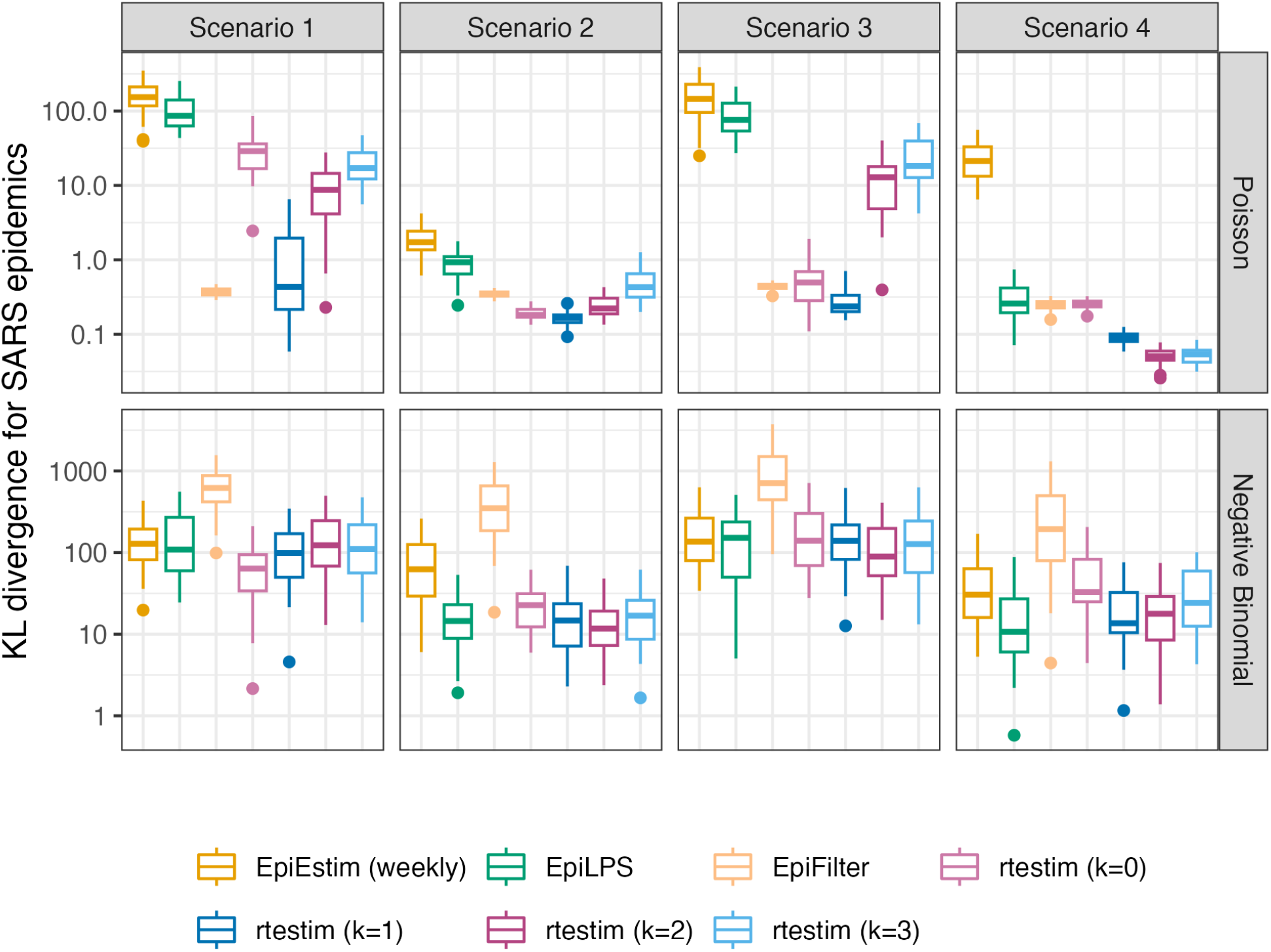
Boxplot of mean KL divergence between 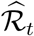 and true ℛ_*t*_ across 50 synthetic SARS epidemics. Performance of each approach given Poisson incidence is in top panels and negative binomial incidence is in bottom panels. The average excludes the first week in all settings, since `EpiEstim` with a weekly sliding window does not provide estimates for the first week. Outliers beyond 1.5*×*IQR of each box are excluded for the sake of comparison with full range of the *y*-axis deferred to Figure A.3.1 in the Supplement.

Figure 5 shows one realization for the estimated instantaneous reproduction number under the Poisson generative model in measles synthetic epidemics for all four scenarios. An expanded visualization with each estimated ℛ_*t*_ curve displayed in a separate panel is provided in Figure A.6.1 in the Supplement. Ignoring the start of the epidemics, all methods look accurate and recover the underlying curves well, except `EpiEstim` with monthly sliding windows, where the trajectories are shifted to the right. Compared to `EpiEstim` and `EpiLPS`, which have rather severe difficulties at the beginning of the period, `rtestim` and `EpiFilter` estimates are more accurate without suffering from the initialization problem. The edge problem in `EpiEstim` and `EpiLPS` may be due to their priors, with the bias persisting for many days. A similar edge problem is possible for `rtestim` though it tends to be less severe for smaller *k*. Besides the edge problem, `EpiEstim` (especially, with the monthly sliding window) and `EpiLPS` produce “smooth” estimated curves that are continuous at the changepoints in Scenarios 1–3, resulting in large errors for a long period. Since the piecewise constant `rtestim` estimator does not enforce any smoothness, it easily captures the sharp change and nearly overlaps with the true values in Scenario 1. For larger *k*, `rtestim` can work nearly as well due to the *𝓁*_1_ penalty’s ability to allow heterogenous smoothness. However, similar to other methods, `rtestim` has some difficulty with the first few timepoints, especially in the periodic scenario, where all methods fail to capture the first peak with much accuracy. `EpiFilter` recovers the ℛ_*t*_ curves well in general, but tends to be more wiggly than other methods.

**Figure 5:**
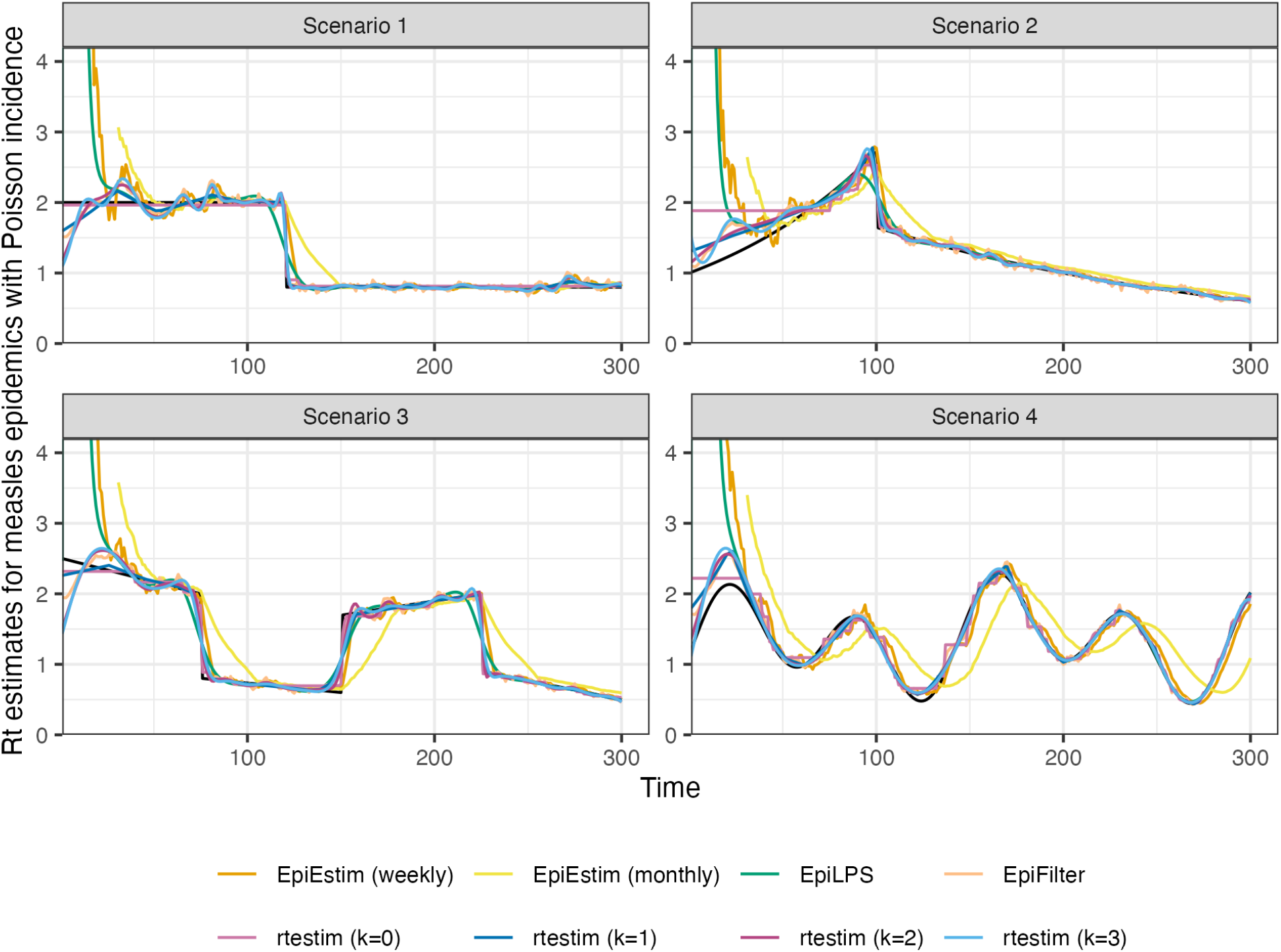
ℛ_*t*_ estimates for realizations of a measles epidemic with Poisson observations. An expanded visualization with each estimated ℛ_*t*_ curve displayed in a separate panel is provided in Figure A.6.1 in the Supplement.

Figure 6 is similar to Figure 5 but shows estimated ℛ_*t*_ given negative binomial incidence in SARS epidemics for each setting. An expanded visualization with each estimated ℛ_*t*_ curve displayed in a separate panel is provided in Figure A.6.4 in the Supplement. Compared to the Figure 5, all methods perform worse overall for two main reasons: larger incidence and overdispersed data. All methods are worse at the start of the epidemics. `EpiFilter` is dramatically wiggly. The performance of `rtestim` is among the best in the first three ℛ_*t*_ scenarios, though it has significant difficulties in the periodic scenario.

**Figure 6:**
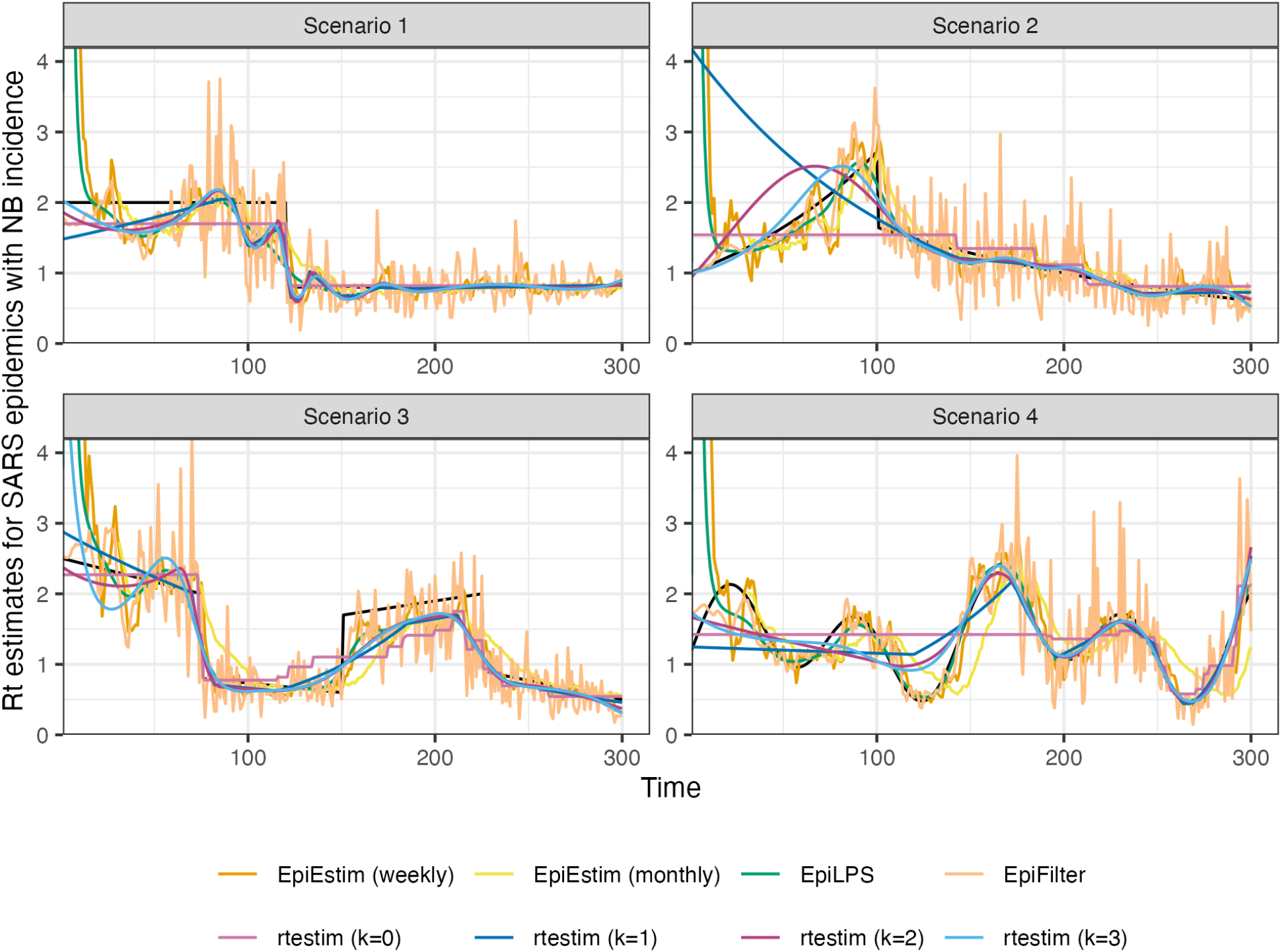
ℛ_*t*_ estimates for realizations of a SARS epidemic with negative binomial observations. An expanded visualization with each estimated ℛ_*t*_ curve displayed in a separate panel is provided in Figure A.6.4 in the Supplement.

Finally, it is important to provide a brief comparison of the running times of all three models across the 8 experimental settings. We find that almost all models across all experiments operate within 10 seconds. Generally, `rtestim` takes the longest due to a relatively large number of estimates—50 values of *λ* and 10 folds of cross validation require 550 estimates— while other models run only a single time for a fixed setting of their hyperparameters per experiment.

### 3.3 Real-data results: Covid-19 incident cases in Canada

We return to the data for Covid-19 confirmed incident cases in Canada examined in Section 1. In this section, we use the weighted average of the serial interval distributions for the four dominant variants (shown in Figure 1) for the purposes of comparison with other methods, none of which allow time-varying delays. The estimates produced by `rtestim` are displayed in Figure 7 while the estimates of all competitors are deferred to Figures A.8.1 and A.8.2 in the Supplement.

**Figure 7:**
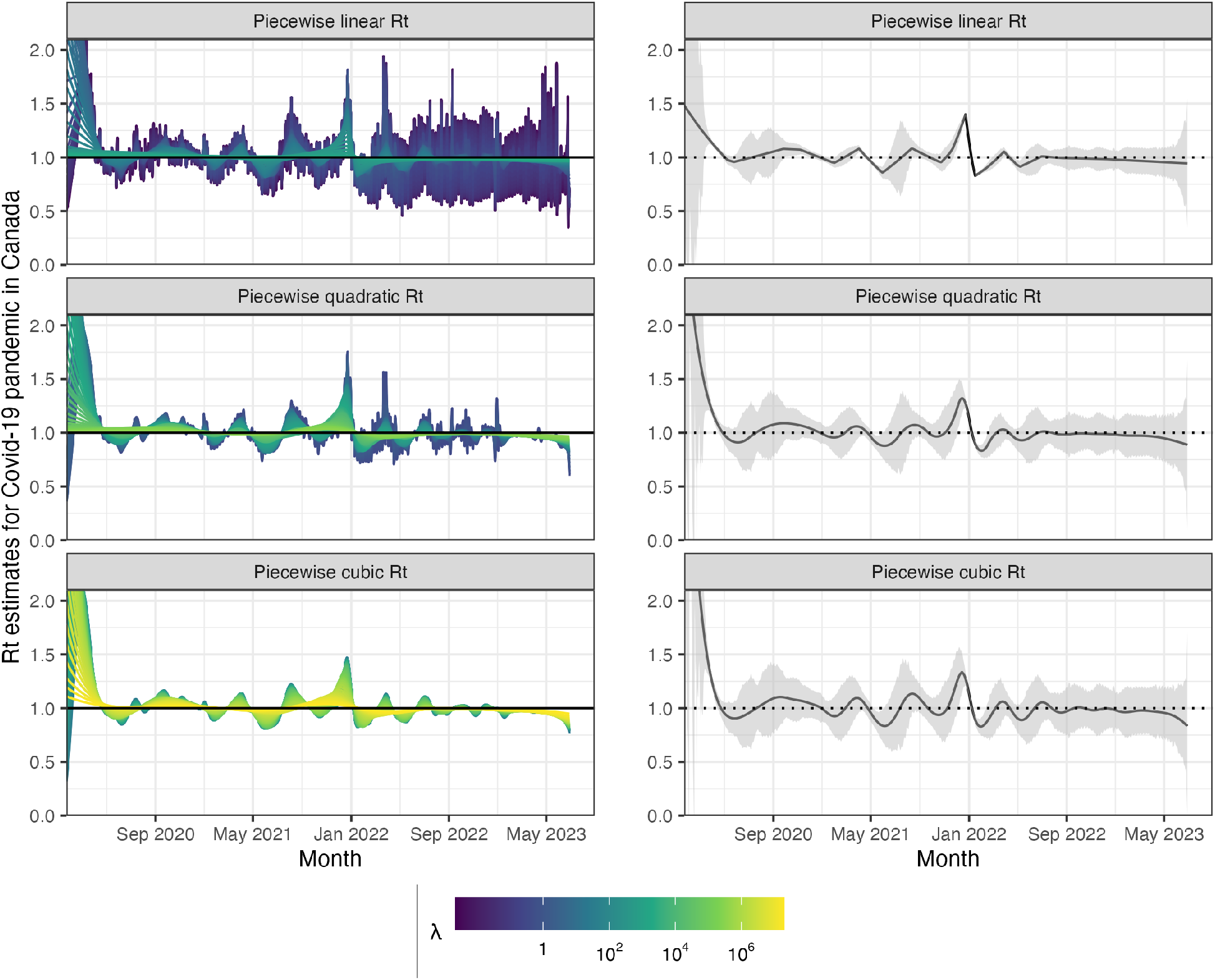
Estimated instantaneous reproduction number based on Covid-19 daily confirmed incident cases. The epidemic is between January 23rd, 2020 and June 28th, 2023 in Canada. The left panels show estimates corresponding to 50 tuning parameters. The right panels show the CV-tuned estimate along with approximate 95% confidence bands. The top, middle and bottom panels show the estimated ℛ_*t*_ using the Poisson trend filtering in Equation (5) with degrees *k* = 1, 2, 3 respectively. All estimates use a constant serial interval distribution, which is the weighted sum of probabilities of the 4 dominant variants used in Figure 1.

Considering *k* = 1, 2 and 3, *ℛ*_*t*_ for Covid-19 in Canada is always less than 2 except at the very early stage, which means that one distinct infected individual on average infects less than two other individuals in the population. Examining three different settings for *k*, the temporal evolution of 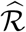 (across all regularization levels *λ*) are similar near the highest peak around the end of 2021 before dropping shortly thereafter. Throughout the estimated curves, the peaks and troughs of the instantaneous reproduction numbers precede the growth and decay cycles of confirmed cases, as expected. We also visualize 95% confidence bands for the point estimates with *λ* chosen by minimizing cross-validated KL divergence in Figure 7.

The estimated instantaneous reproduction numbers are relatively unstable before April, 2022. The highest peak coincides with the emergence and global spread of the Omicron variant. The estimated instantaneous reproduction numbers fall below 1 during a few time periods, where the most obvious troughs are roughly from April 2021 to July 2021 and from January, 2022 to April 2022. The first trough coincides with the introduction of Covid-19 vaccines in Canada. The second trough, shortly after the largest peak may be due to variety of factors resulting in the depletion of the susceptible population such as increased self-isolation in response to media coverage of the peak or immunity incurred via recent infection. Since April 2022, the estimated instantaneous reproduction number has remained relatively stable (fluctuating around one) corresponding to low reported cases, though reporting behaviours also changed significantly since the Omicron wave.

### 3.4 Real-data results: influenza in Baltimore, Maryland, 1918

We also apply `rtestim` to daily reported influenza cases in Baltimore, Maryland occurring during the world-wide pandemic of 1918 from September to November (Frost and Sydenstricker, 1919). The data, shown in Figure 8, is included in the `EpiEstim` `R` package, along with the serial interval distribution. The 1918 influenza outbreak, caused by the H1N1 influenza A virus, was unprecedentedly deadly with case fatality rate over 2.5%, infecting almost one-third of the population across the world (Taubenberger and Morens, 2006). The CV-tuned piecewise cubic estimates in Figure 9 better capture the growth at the beginning of the pandemic in Figure 8. The estimated ℛ_*t*_ curve suggests that the transmissibility of the pandemic grew rapidly over the first 30 days before declining below one after 50 days. However, it also suggests an increase in infectiousness toward the end of the period. With this data, it is difficult to determine if there is a second wave or a steady decline ahead. The CV-tuned piecewise constant and linear estimates in Figure 9 both suggest a steady decline. This conclusion is supported by the fact that incident cases decline to zero at the end of the period, matching *ℛ*_*t*_ estimates in Cori et al. (2013), which are all lower than one. Results from alternative software is deferred to Figures A.8.3 and A.8.4 in the Supplement.

**Figure 8:**
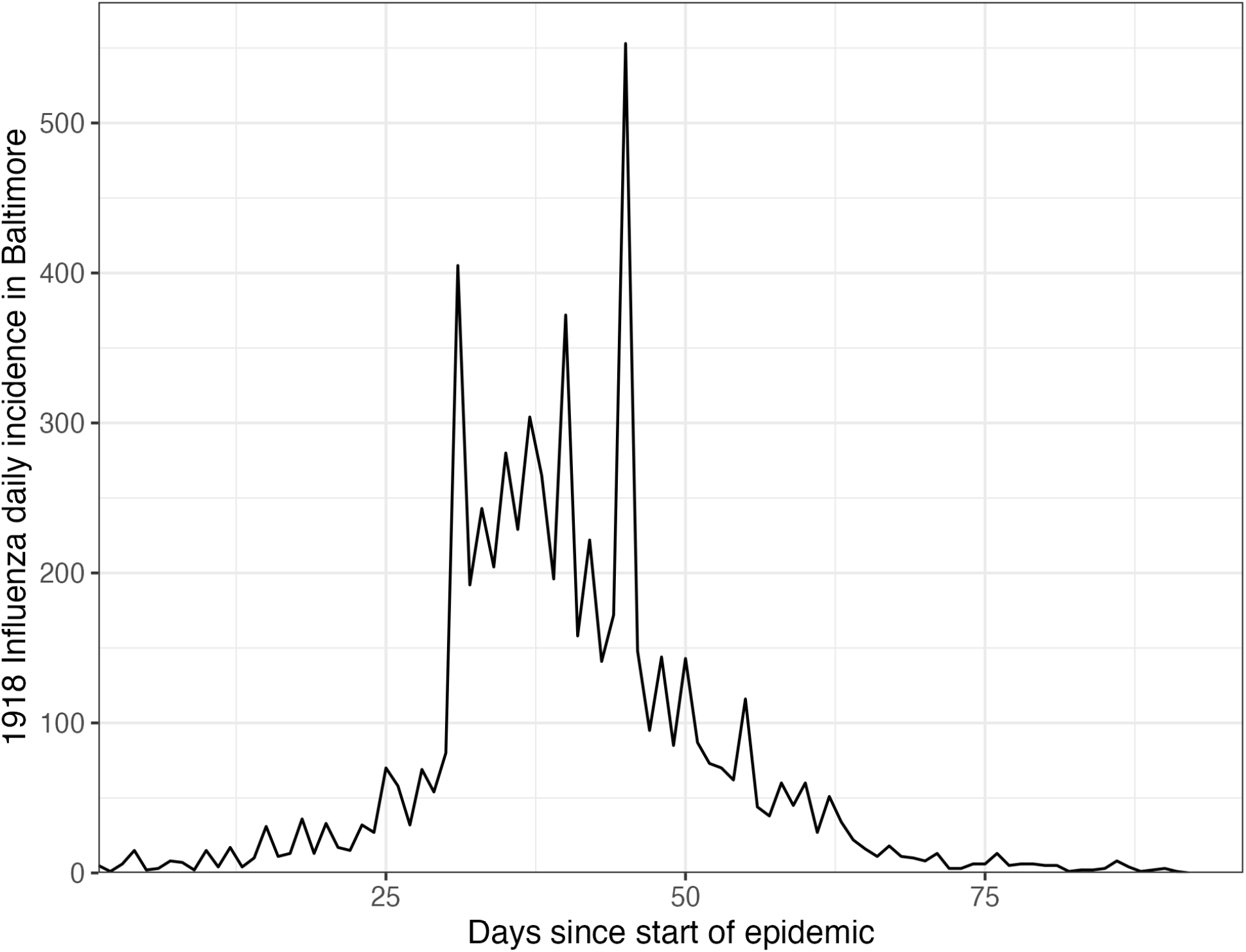
Daily incident influenza cases in Baltimore, Maryland between September and November 1918.

**Figure 9:**
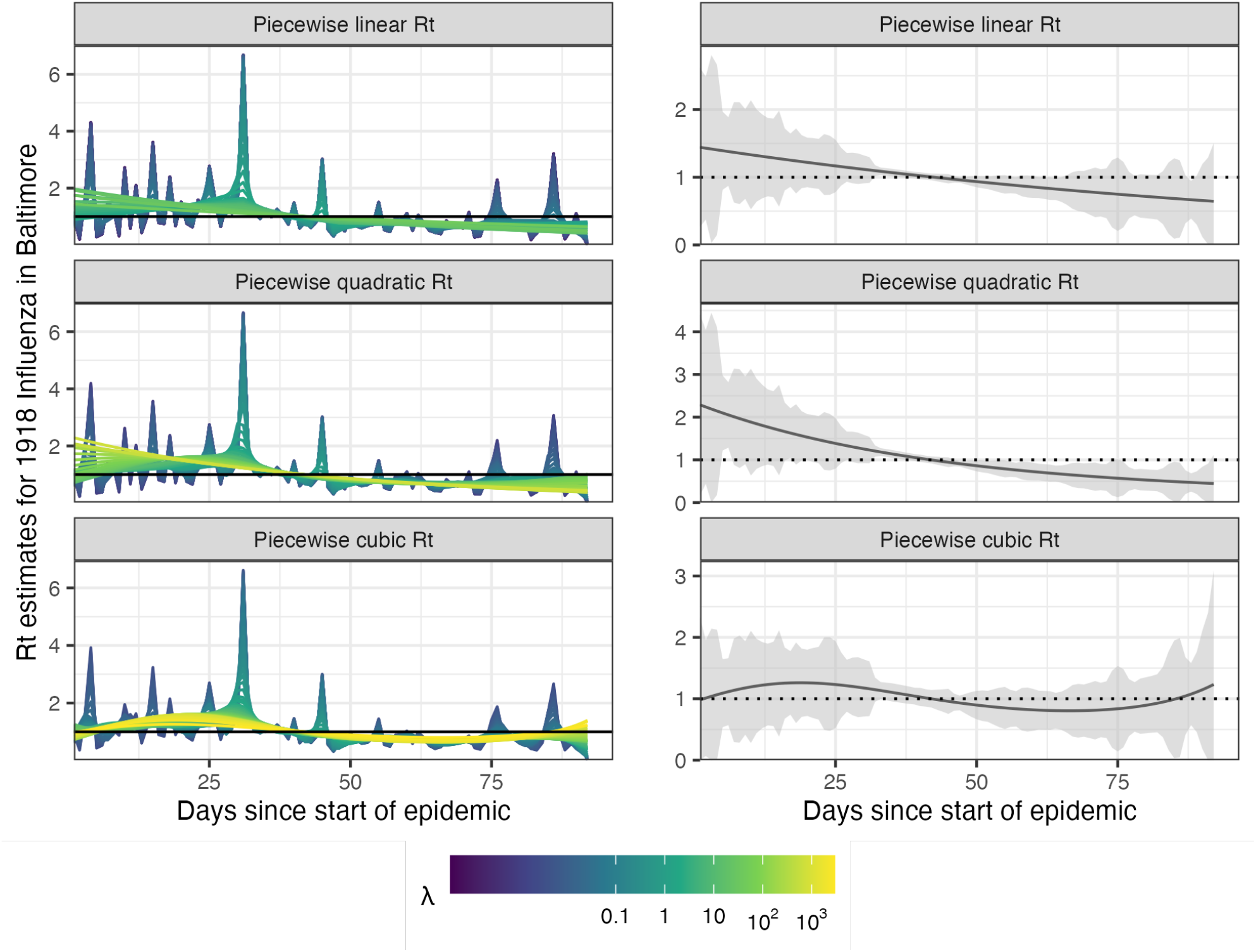
Estimated instantaneous reproduction numbers for influenza in Baltimore, Maryland in 1918. The left panels show estimates for a set of 50 tuning parameters. The right column displays the CV-tuned estimates with approximate 95% confidence bands. The rows (top to bottom) show 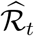 using Poisson trend filtering with *k* = 1, 2, 3 respectively.

## 4 Discussion

Our methodology provides a locally adaptive estimator using Poisson trend filtering. It captures the heterogeneous smoothness of instantaneous reproduction numbers given observed incidence data rather than resulting in global smoothness. This is a nonparametric regression model which can be written as a convex optimization problem. Minimizing the negative logliklihood of observations guarantees data fidelity while the penalty on divided differences between pairs of neighbouring parameters imposes smoothness. The *𝓁*_1_-regularization results in sparsity of the divided differences, leading to heterogeneous smoothness across time.

The property of local adaptivity (heterogenous smoothness) is useful to automatically distinguish, for example, seasonal outbreaks from outbreaks driven by other factors (behavioural changes, foreign introduction, etc.). Given a well-chosen polynomial degree, the growth rates can be quickly detected, potentially advising public health authorities to implement policy changes. The instantaneous reproduction numbers can be estimated retrospectively to examine the efficacy of such policies, whether they result in ℛ_*t*_ falling below 1 or the speed of their effects. The smoothness of ℛ_*t*_ curves (including the polynomial degrees and tuning parameters) should be chosen based on the purpose of the study in practice or with data-driven risk estimation by cross validation.

Our method provides a natural way to deal with missing data, for example, on weekends and holidays or due to changes in reporting frequency. While solving the convex optimization problem, our method can easily handle uneven spacing or irregular reporting. Computing the total primary infectiousness is also easily generalized to irregular reporting through automatic modifications of the discretization of the serial interval distribution. However, there are many other aspects to be considered in choosing the delay distribution to improve accuracy (Park et al., 2024). Additionally, because the *𝓁*_1_ penalty introduces sparsity (operating like a median rather than a mean), this procedure is relatively insensitive to spurious outliers compared to *𝓁*_2_ regularization.

There are a number of limitations that may influence the quality of ℛ_*t*_ estimation. While our model is generic for incidence data rather than tailored to any specific disease, it does assume that the generation interval is short relative to the period of data collection. More specialized methodologies would be required for diseases with long incubation periods such as HIV or Hepatitis. Our approach, does not explicitly model imported cases, nor distinguish between subpopulations that may have different mixing behaviour. However, a natural extension to handle imported cases is to follow the suggested procedure of Thompson et al. (2019). By including imported cases only in *η*_*t*_ rather than in both *y*_*t*_ and *η*_*t*_, we exclude individuals who were infected elsewhere, lowering ℛ_*t*_, but correctly reflecting the number of new primary infectees.

While the Poisson assumption is common, it does not handle overdispersion (observation variance larger than the mean). The negative binomial distribution is a good alternative, but more difficult to estimate in this context. As described in Section 1, the expression for ℛ assumes that a relatively constant proportion of true infections is reported. However, if this proportion varies with time (say, due to changes in surveillance practices or testing recommendations), the estimates may be biased over this window. A good example is in early January 2022, during the height of the Omicron wave, Canada moved from testing all symptomatic individuals to testing only those in at-risk groups. The result was a sudden change that would render ℛ_*t*_ estimates on either side of this timepoint incommensurable.

Our implementation in the software package `rtestim` can accommodate a fixed serial interval throughout the period of study (as implemented in simulation and in the real epidemics) or use time-varying serial interval distributions (as implemented in Figure 1 for Covid-19 data in Canada). In reality, the serial interval may vary due to changes in the factors such as population immunity (Nash et al., 2023). One issue regarding the serial interval distribution relates to equating serial and generation intervals (also mentioned above). The serial interval distribution is generally wider than that of the generation interval, because the serial interval involves the convolution of two distributions, and is unlikely to actually follow a named distribution like gamma, though it may be reasonably well approximated by one. Our implementation allows for an arbitrary distribution to be used, but requires the user to specify the discretization explicitly, requiring more nuanced knowledge than is typically available. Pushing this analysis further, to accommodate other types of incidence data (hospitalizations or deaths), a modified generation interval distribution would be necessary, and further assumptions would be required as well. Or else, one would first need to deconvolve deaths to infection onset before using our software.

Accurate statistical coverage of a function is a difficult problem, and the types of (frequentist) guarantees that can be made are not always what one would want (Genovese and Wasserman, 2008). We examine the coverage of our approximate confidence interval in simulation, with details are deferred to Section A.6 in the Supplement. Empirically, our observations for our method, as well as all others we have seen, follow a similar (undesirable) pattern: when ℛ_*t*_ is stable, they over cover dramatically (even implausibly narrow intervals have 100% coverage); but when ℛ_*t*_ changes abruptly, they under cover. Theoretically, whether these intervals should be expected to provide (1 *− α*)% coverage simultaneously over all time while being narrow enough to provide useful uncertainty quantification is neither easy nor settled. An alternative to our approximation in Section 2.5, which we defer to future work, is to use the data fission method proposed by Leiner et al. (2023), which provides post-selection inference for trend filtering.

Nonetheless, our methodology is implemented in a lightweight `R` package `rtestim` and computed efficiently, especially for large-scale data, with a proximal Newton solver coded in C++. Given available incident case data, prespecified serial interval distribution, and a choice of degree *k*, `rtestim` is able to produce accurate estimates of instantaneous reproduction number and provide efficient tuning parameter selection via cross validation.

## Data Availability

All data produced are available online at the "jiapivialiu/rt-est-manuscript" Github repository with the following url: https://github.com/jiapivialiu/rt-est-manuscript/tree/main

## Acknowledgments

This research was enabled in part by support provided by BC DRI group who manages Cedar cloud (https://docs.alliancecan.ca/wiki/Cedar) and the Digital Research Alliance of Canada (alliancecan.ca).

## References

Abbott, S., Funk, S., Hickson, J., Badr, H. S., Monticone, P., Ellis, P., Azam, J., Munday, J., Allen, J., Johnson, A., Pearson, C. A. B., actions user, Chapman, L., DeWitt, M., Bosse, N. and Meakin, S. (2023) epiforecasts/EpiNow2: 1.4.0 release. 10.5281/zenodo.8380568.

Abbott, S., Hellewell, J., Thompson, R. N., Sherratt, K., Gibbs, H. P., Bosse, N. I., Munday, J. D., Meakin, S., Doughty, E. L., Chun, J. Y. et al. (2020) Estimating the time-varying reproduction number of SARS-CoV-2 using national and subnational case counts. Wellcome Open Research, 5, 112. 10.12688/wellcomeopenres.16006.2.

Abry, P., Pustelnik, N., Roux, S., Jensen, P., Flandrin, P., Gribonval, R., Lucas, C.-G., Guichard, É., Borgnat, P. and Garnier, N. (2020) Spatial and temporal regularization to estimate COVID-19 reproduction number R(t): Promoting piecewise smoothness via convex optimization. PLoS ONE, 15, e0237901. 10.1371/journal.pone.0237901.

Anderson, R. M. and May, R. M. (1991) Infectious diseases of humans: dynamics and control. Oxford university press.

Azmon, A., Faes, C. and Hens, N. (2014) On the estimation of the reproduction number based on misreported epidemic data. Statistics in Medicine, 33, 1176–1192. 10.1002/sim.6015.

Bergmeir, C., Hyndman, R. J. and Koo, B. (2018) A note on the validity of cross-validation for evaluating autoregressive time series prediction. Computational Statistics & Data Analysis, 120, 70–83. URL: https://www.sciencedirect.com/science/article/pii/S0167947317302384.

Berry, I., O’Neill, M., Sturrock, S. L., Wright, J. E., Acharya, K., Brankston, G., Harish, V., Kornas, K., Maani, N., Naganathan, T., Obress, L., Rossi, T., Simmons, A. E., Camp, M. V., Xie, X., Tuite, A. R., Greer, A. L., Fisman, D. N. and Soucy, J.-P. R. (2021) A sub-national real-time epidemiological and vaccination database for the COVID-19 pandemic in canada. Scientific Data, 8. URL: 10.1038/s41597-021-00955-2.

CAMEO Group, C. V. R. R. N. (2023) Covarr-net/duotang: Release for zenodo archive. URL: 10.5281/zenodo.10367461.

Cori, A., Cauchemez, S., Ferguson, N. M., Fraser, C., Dahlqwist, E., Demarsh, P. A., Jombart, T., Kamvar, Z. N., Lessler, J., Li, S. et al. (2020) EpiEstim: estimate time varying reproduction numbers from epidemic curves. Version 2.2-4. https://cran.r-project.org/package=EpiEstim.

Cori, A., Ferguson, N. M., Fraser, C. and Cauchemez, S. (2013) A new framework and software to estimate time-varying reproduction numbers during epidemics. American Journal of Epidemiology, 178, 1505–1512. 10.1093/aje/kwt133.

Cori, A., Kamvar, Z., Stockwin, J., Jombart, T., Dahlqwist, E., FitzJohn, R., Thompson, R., Nash, R., Wardle, J. and Bhatia, S. (2022) EpiEstim v2.2-4: A tool to estimate time varying instantaneous reproduction number during epidemics. https://github.com/mrc-ide/EpiEstim.

Eales, O. and Riley, S. (2024) Differences between the true reproduction number and the apparent reproduction number of an epidemic time series. Epidemics, 46, 100742.

Fraser, C. (2007) Estimating individual and household reproduction numbers in an emerging epidemic. PloS one, 2, e758.

Frost, W. H. and Sydenstricker, E. (1919) Influenza in Maryland: preliminary statistics of certain localities. Public Health Reports (1896-1970), 491–504.

Genovese, C. and Wasserman, L. (2008) Adaptive confidence bands. The Annals of Statistics, 36, 875–905. URL: 10.1214/07-AOS500.

Goldstein, I. H., Parker, D. M., Jiang, S. and Minin, V. M. (2023) Semiparametric inference of effective reproduction number dynamics from wastewater pathogen surveillance data. arXiv preprint 2308.15770.

Goldstein, I. H., Wakefield, J. and Minin, V. M. (2024) Incorporating testing volume into estimation of effective reproduction number dynamics. Journal of the Royal Statistical Society Series A: Statistics in Society, 187, 436–453.

Gostic, K. M., McGough, L., Baskerville, E. B., Abbott, S., Joshi, K., Tedijanto, C., Kahn, R., Niehus, R., Hay, J. A., De Salazar, P. M. et al. (2020) Practical considerations for measuring the effective reproductive number, Rt. PLoS Computational Biology, 16, e1008409. 10.1371/journal.pcbi.1008409.

Gressani, O. (2021) EpiLPS: A Fast and Flexible Bayesian Tool for Estimating Epidemiological Parameters. https://epilps.com/.

Gressani, O., Faes, C. and Hens, N. (2022a) An approximate Bayesian approach for estimation of the instantaneous reproduction number under misreported epidemic data. Biometrical Journal, 65, 2200024. 10.1002/bimj.202200024.

Gressani, O., Wallinga, J., Althaus, C. L., Hens, N. and Faes, C. (2022b) Epilps: A fast and flexible Bayesian tool for estimation of the time-varying reproduction number. PLoS Computational Biology, 18, e1010618. 10.1371/journal.pcbi.1010618.

Groendyke, C., Welch, D. and Hunter, D. R. (2011) Bayesian inference for contact networks given epidemic data. Scandinavian Journal of Statistics, 38, 600–616.

Hao, X., Cheng, S., Wu, D., Wu, T., Lin, X. and Wang, C. (2020) Reconstruction of the full transmission dynamics of covid-19 in wuhan. Nature, 584, 420–424.

Hettinger, G., Rubin, D. and Huang, J. (2023) Estimating the instantaneous reproduction number with imperfect data: a method to account for case-reporting variation and serial interval uncertainty. arXiv preprint 2302.12078. 10.48550/arXiv.2302.12078.

Hitchings, M. D., Dean, N. E., García-Carreras, B., Hladish, T. J., Huang, A. T., Yang, B. and Cummings, D. A. (2021) The usefulness of the test-positive proportion of severe acute respiratory syndrome coronavirus 2 as a surveillance tool. American Journal of Epidemiology, 190, 1396–1405. 10.1093/aje/kwab023.

Ho, F., Parag, K. V., Adam, D. C., Lau, E. H., Cowling, B. J. and Tsang, T. K. (2023) Accounting for the potential of overdispersion in estimation of the time-varying reproduction number. Epidemiology, 34, 201–205. 10.1097/ede.0000000000001563.

Jin, S., Dickens, B. L., Lim, J. T. and Cook, A. R. (2023) EpiMix: A novel method to estimate effective reproduction number. Infectious Disease Modelling, 8, 704–716. 10.1016/j.idm.2023.06.002.

Kim, S.-J., Koh, K., Boyd, S. and Gorinevsky, D. (2009) 𝓁1 trend filtering. SIAM Review, 51, 339–360. 10.1137/070690274.

Leiner, J., Duan, B., Wasserman, L. and Ramdas, A. (2023) Data fission: splitting a single data point. Journal of the American Statistical Association, 1–12.

Lipsitch, M., Cohen, T., Cooper, B., Robins, J. M., Ma, S., James, L., Gopalakrishna, G., Chew, S. K., Tan, C. C., Samore, M. H. et al. (2003) Transmission dynamics and control of severe acute respiratory syndrome. science, 300, 1966–1970.

Lison, A., Abbott, S., Huisman, J. and Stadler, T. (2024) Generative Bayesian modeling to nowcast the effective reproduction number from line list data with missing symptom onset dates. PLoS Computational Biology, 20, e1012021.

Nash, R. K., Bhatt, S., Cori, A. and Nouvellet, P. (2023) Estimating the epidemic reproduction number from temporally aggregated incidence data: A statistical modelling approach and software tool. PLoS Computational Biology, 19, e1011439. 10.1371/journal.pcbi.1011439.

Nishiura, H. and Chowell, G. (2009) The effective reproduction number as a prelude to statistical estimation of time-dependent epidemic trends. Mathematical and statistical estimation approaches in epidemiology, 103–121.

Parag, K. V. (2020) EpiFilter. https://github.com/kpzoo/EpiFilter?tab=readme-ov-file.

Parag, K. V. (2021) Improved estimation of time-varying reproduction numbers at low case incidence and between epidemic waves. PLoS Computational Biology, 17, e1009347. 10.1371/journal.pcbi.1009347.

Park, S. W., Akhmetzhanov, A. R., Charniga, K., Cori, A., Davies, N. G., Dushoff, J., Funk, S., Gostic, K., Grenfell, B., Linton, N. et al. (2024) Estimating epidemiological delay distributions for infectious diseases. medRxiv, 2024–01.

Park, S. W., Sun, K., Champredon, D., Li, M., Bolker, B. M., Earn, D. J., Weitz, J. S., Grenfell, B. T. and Dushoff, J. (2021) Forward-looking serial intervals correctly link epidemic growth to reproduction numbers. Proceedings of the National Academy of Sciences, 118, e2011548118.

Pascal, B., Abry, P., Pustelnik, N., Roux, S., Gribonval, R. and Flandrin, P. (2022) Nonsmooth convex optimization to estimate the Covid-19 reproduction number space-time evolution with robustness against low quality data. IEEE Transactions on Signal Processing, 70, 2859–2868. 10.1109/TSP.2022.3180926.

Pellis, L., Scarabel, F., Stage, H. B., Overton, C. E., Chappell, L. H., Fearon, E., Bennett, E., Lythgoe, K. A., House, T. A., Hall, I. et al. (2021) Challenges in control of COVID-19: short doubling time and long delay to effect of interventions. Philosophical Transactions of the Royal Society B, 376, 20200264. 10.1098/rstb.2020.0264.

Pircalabelu, E. (2023) A spline-based time-varying reproduction number for modelling epidemiological outbreaks. Journal of the Royal Statistical Society Series C: Applied Statistics, 72, 688–702. 10.1093/jrsssc/qlad027.

Pitzer, V. E., Chitwood, M., Havumaki, J., Menzies, N. A., Perniciaro, S., Warren, J. L., Weinberger, D. M. and Cohen, T. (2021) The impact of changes in diagnostic testing practices on estimates of COVID-19 transmission in the United States. American Journal of Epidemiology, 190, 1908–1917. 10.1093/aje/kwab089.

Sadhanala, V., Bassett, R., Sharpnack, J. and McDonald, D. J. (2024) Exponential family trend filtering on lattices. Electronic Journal of Statistics, 18, 1749–1814.

Taubenberger, J. K. and Morens, D. M. (2006) 1918 Influenza: the mother of all pandemics. Emerging Infectious Diseases, 17, 69–79. 10.3201%2Feid1201.050979.

Thompson, R. N., Stockwin, J. E., van Gaalen, R. D., Polonsky, J. A., Kamvar, Z. N., Demarsh, P. A., Dahlqwist, E., Li, S., Miguel, E., Jombart, T. et al. (2019) Improved inference of time-varying reproduction numbers during infectious disease outbreaks. Epidemics, 29, 100356. 10.1016/j.epidem.2019.100356.

Tibshirani, R. J. (2014) Adaptive piecewise polynomial estimation via trend filtering. The Annals of Statistics, 42, 285–323. 10.1214/13-AOS1189.

Tibshirani, R. J. (2022) Divided differences, falling factorials, and discrete splines: Another look at trend filtering and related problems. Foundations and Trends® in Machine Learning, 15, 694–846. 10.1561/2200000099.

Trevisin, C., Bertuzzo, E., Pasetto, D., Mari, L., Miccoli, S., Casagrandi, R., Gatto, M. and Rinaldo, A. (2023) Spatially explicit effective reproduction numbers from incidence and mobility data. Proceedings of the National Academy of Sciences, 120, e2219816120. 10.1073/pnas.2219816120.

Vaiter, S., Deledalle, C., Fadili, J., Peyré, G. and Dossal, C. (2017) The degrees of freedom of partly smooth regularizers. Annals of the Institute of Statistical Mathematics, 69, 791–832. 10.1007/s10463-016-0563-z.

Wallinga, J. and Teunis, P. (2004) Different epidemic curves for severe acute respiratory syndrome reveal similar impacts of control measures. American Journal of epidemiology, 160, 509–516.

Xu, X., Wu, Y., Kummer, A. G., Zhao, Y., Hu, Z., Wang, Y., Liu, H., Ajelli, M. and Yu, H. (2023) Assessing changes in incubation period, serial interval, and generation time of sars-cov-2 variants of concern: a systematic review and meta-analysis. BMC medicine, 21, 374.

