## supplement for "rtestim: Time-varying reproduction number estimation with trend filtering"

**A.8 Application of `rtestim` and all competitors on real epidemics** **38**

**References** **44**

#### A.1 Derivation of Kullback Leibler divergence

We provide the detailed derivation of the Kullback Leibler (KL) divergence in Eq. (11) in the manuscript that is used to compare the accuracy of the estimated time-varying instantaneous reproduction number with the true ones. Given the total infectiousness $\eta$, we compare the distance between the Poisson distributions $y \sim \text{Pois}(\eta\widehat{\mathcal{R}})$ and $y \sim \text{Pois}(\eta\mathcal{R})$, where $y, \mathcal{R} \in \mathbb{N}_0^n$ are natural numbers including 0, $\eta \in \mathbb{R}^n$, and $f_0(y; \eta, \mathcal{R}) =$

$\prod_{t=1}^{n} \frac{(\eta_t \mathcal{R}_t)^{y_t} e^{-\eta_t \mathcal{R}_t}}{y_t!}$, $f_1(y; \eta, \widehat{\mathcal{R}}) = \prod_{t=1}^{n} \frac{(\eta_t \widehat{\mathcal{R}}_t)^{y_t} e^{-\eta_t \widehat{\mathcal{R}}_t}}{y_t!}$ are the corresponding density mass functions for independent $y_t, t = 1, \cdots, n$. Because this is a natural exponential family with log-partition function $\exp(\cdot)$ and parameter $\log(\eta_t \mathcal{R}_t)$, then, the KL divergence between them can be written in terms of the Bregman divergence for exp, e.g. Wainwright and Jordan (2008),

$$
\begin{aligned}
&D_{KL}(\mathcal{R} \parallel \widehat{\mathcal{R}}) \\
&= D_{KL}(f_0(y; \eta, \mathcal{R}) \parallel f_1(y; \eta, \widehat{\mathcal{R}})) \\
&= D_{KL}\left( \prod_{t=1}^{n} f_0(y_t; \eta_t, \mathcal{R}_t) \parallel \prod_{t=1}^{n} f_1(y_t; \eta_t, \widehat{\mathcal{R}}_t) \right) \\
&= \sum_{t=1}^{n} D_{KL}\left( f_0(y_t; \eta_t, \mathcal{R}_t) \parallel f_1(y_t; \eta_t, \widehat{\mathcal{R}}_t) \right), \ (y_t \text{ are independent, conditional on } \mathcal{R}_t, \widehat{\mathcal{R}}_t, \eta_t) \\
&= \sum_{t=1}^{n} \exp\left(\log(\eta_t \widehat{\mathcal{R}}_t)\right) - \exp\left(\log(\eta_t \mathcal{R}_t)\right) + \exp\left(\log(\eta_t \mathcal{R}_t)\right) \log \frac{\eta_t \mathcal{R}_t}{\eta_t \widehat{\mathcal{R}}_t}, \ (\text{definition of Bregman divergence}) \\
&= \sum_{t=1}^{n} \eta_t \widehat{\mathcal{R}}_t - \eta_t \mathcal{R}_t + \eta_t \mathcal{R}_t \log \frac{\mathcal{R}_t}{\widehat{\mathcal{R}}_t} \\
&= \sum_{t=1}^{n} \eta_t \left( \mathcal{R}_t \log \frac{\mathcal{R}_t}{\widehat{\mathcal{R}}_t} + \widehat{\mathcal{R}}_t - \mathcal{R}_t \right).
\end{aligned}
$$

We use mean KL divergence (denoted, $\overline{D}_{KL}(\mathcal{R} \parallel \widehat{\mathcal{R}}) := D_{KL}(\mathcal{R} \parallel \widehat{\mathcal{R}})/N$, which is the KL divergence divided by the sequence length) in experiments for accuracy comparison.

#### A.2 Additional details on experimental settings

We compare the accuracy of the estimated instantaneous reproduction numbers using the average Kullback Leibler (KL) divergence under Poisson incidence (we say mean KL divergence for short when clear) in (11) across `rtestim` and several alternative methods, including `EpiEstim` with weekly and monthly sliding windows, `EpiLPS`, `EpiFilter`, `EpiNow2`, and `rtestim` with degrees k=0,1,2,3, resulting in 9 methods in total. We consider two durations of epidemics with $n = 50$ or $n = 300$ timepoints respectively. Since `EpiNow2` may take too long to converge (for example, a long `measles` epidemic requires nearly 2 hours to converge on the Cedar cluster provided by Digital Research Alliance of Canada), we only compare it with other methods for short `flu` epidemics.

We use the serial interval (SI) distributions of `measles` and `SARS` to generate long synthetic epidemics, and `flu` for short epidemics, inspired by Cori et al. (2013) which used the SI from real epidemics to illustrate the performance of their method. The means and standard deviations of the SI distributions are based on existing studies; specifically, $(14.9, 3.9)$ for `measles` (Groendyke, Welch, and Hunter (2011)), $(8.4, 3.8)$ for `SARS` (Lipsitch et al. (2003)), and $(2.6, 1.5)$ for `flu` (Ferguson et al. (2005), Boëlle et al. (2011)). Incident cases in synthetic `measles` epidemics are relatively low (generally $< 1000$ at the peak), while `SARS` incident cases are relatively large (between 15000 and 20000 at the peak).

We consider reasonably large overdispersion for negative binomial incidence with size 5. Figure A.2.1 displays the ratio of the variance to the mean across different settings using the same set of sample epidemics in Fig 5 and Fig 6, and all figures in Section A.6.1. For Poisson, this ratio is constant at 1. However, the negative binomial incidence results in significant overdispersion.

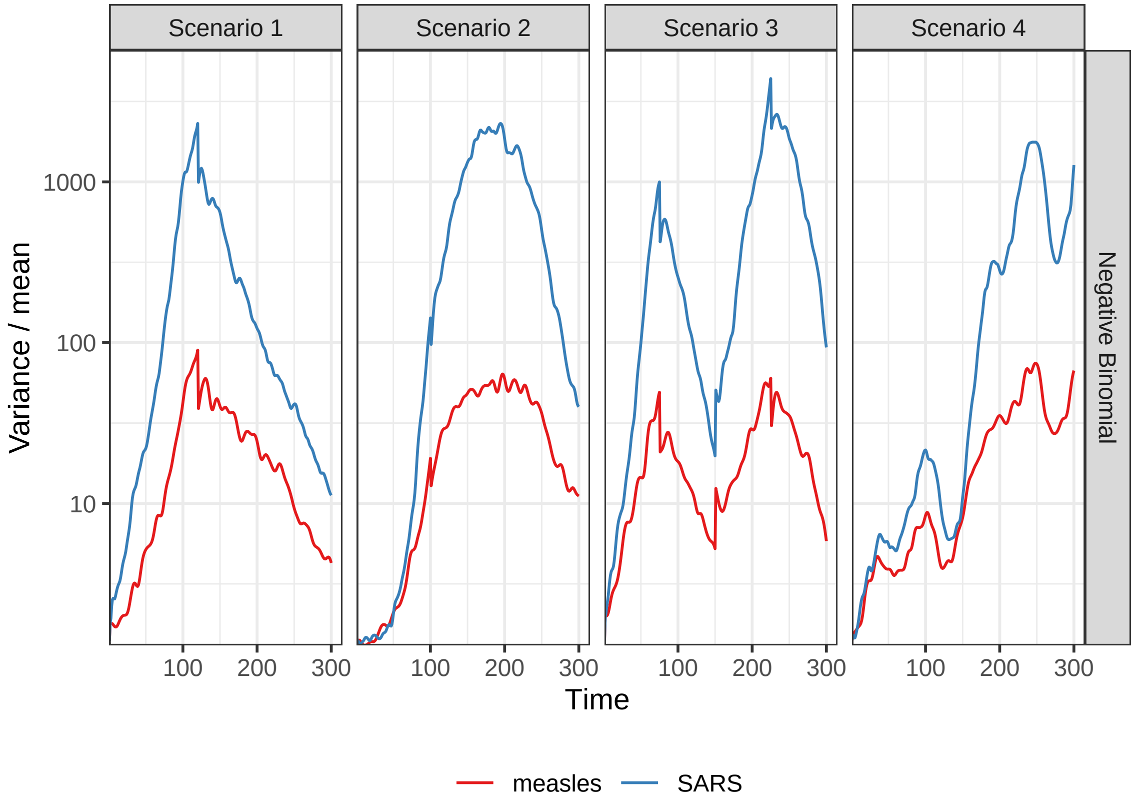

Figure A.2.1: Dispersion level of incidence for sample negative binomial epidemics.

In model fitting, we use both true and misspecified serial interval (SI) distributions to test the robustness of our method, compared with alternatives. The misspecification of serial interval distributions are either "mild" or "major", where, in the major misspecification, we use a completely different pair of SI parameters, e.g., we use the SI of `SARS` for generated `measles` epidemics, and the SI of `measles` for generated `flu` epidemics. In the mild SI misspecification, we consider slightly adjusted parameters for both `measles` and `flu` epidemics, where the mean is decreased by 2 for `measles` and increased by 2 for `flu` and the standard deviation is increased by 10%, denoted as `adj_flu` and `adj_measles` respectively. These settings result in 7 pairs of SI distributions (for epidemic generating, model fitting): (`measles`, `measles`), (`SARS`, `SARS`), (`measles`, `adj_measles`), (`measles`, `SARS`) for long epidemics and (`flu`, `flu`), (`flu`, `adj_flu`), (`flu`, `measles`) for short epidemics. Figure A.2.2 displays all SI distributions (`measles`, `adj_measles`, `SARS`, `flu`, and `adj_flu`) used in the experiments.

Table 1 summarizes the aforementioned experimental setting for accuracy comparison: Poisson and negative binomial (NB) distributions for incidence and four $\mathcal{R}_t$ scenarios are used for all long epidemics. We only consider one $\mathcal{R}_t$ scenario (Scenario 3: piecewise linear $\mathcal{R}_t$) for short epidemics. Each experimental setting is replicated 50 times, which yields 12800 experiments for long epidemics and 2700 for short epidemics.

We visualize the selected key results of the accuracy comparison using long synthetic epidemics in Section 3.2 in the manuscript. Other main experimental results are displayed in Section A.3.

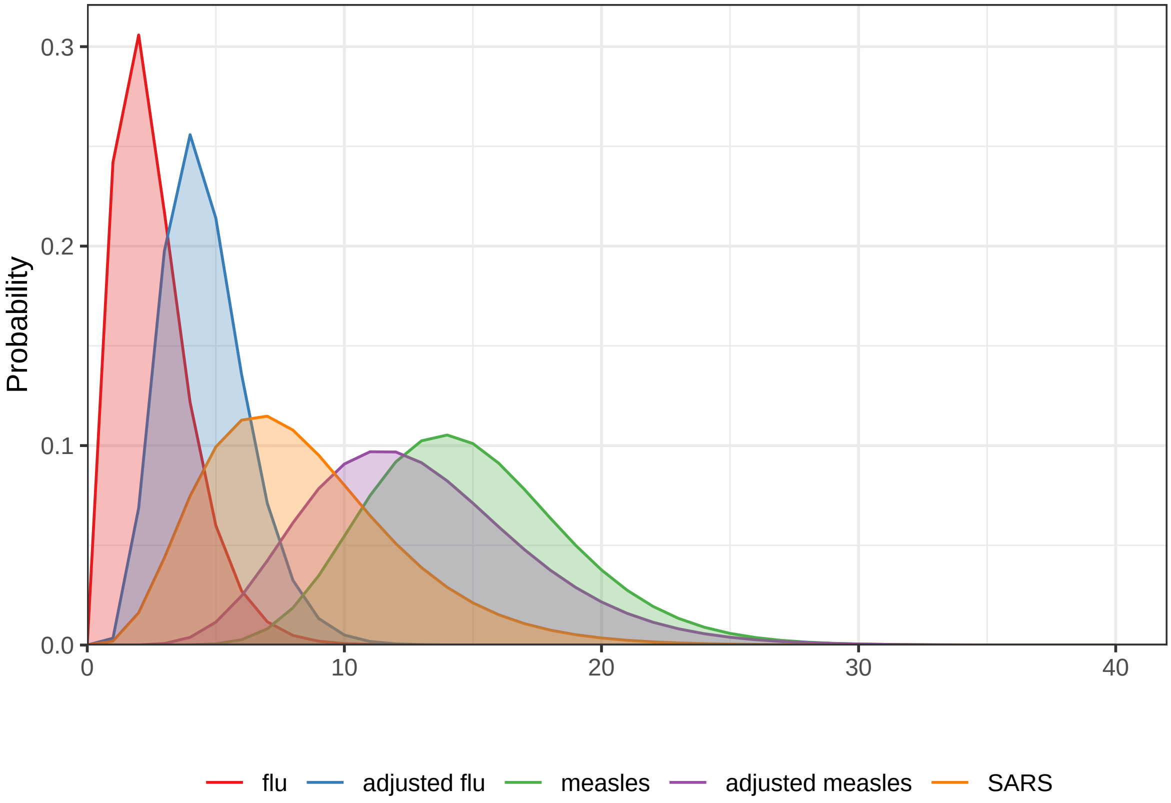

Figure A.2.2: Probability densities of serial interval distributions used in the experiments.

Table 1: Summary of experimental settings on accuracy comparison.

| Length | SI | Rt scenario | Incidence | SI for modelling | Method |
|---|---|---|---|---|---|
| 300 | measles | 1-4 | Poisson, NB | measles, adj-measles, SARS | 8 methods |
| 300 | SARS | 1-4 | Poisson, NB | SARS | 8 methods |
| 50 | flu | 3 | Poisson, NB | flu, adj-flu, measles | 9 methods |

#### A.3 Additional accuracy comparisons

##### A.3.1 Long epidemics

We have displayed the accuracy of all methods (where `EpiEstim` uses a weekly sliding window) for `measles` and `SARS` sample epidemics using KL divergence excluding the first week since `EpiEstim` does not provide estimates in the first week in Fig 3 and Fig 4 in the manuscript, where we exclude the outliers. A full visualization including the outliers is in Figure A.3.1.

Figure A.3.2 compares `EpiEstim` with *monthly* sliding windows with other methods. We average the KL divergence per coordinate excluding the timepoints in the first month for all approaches, since `EpiEstim` estimates with monthly sliding windows are not available until the second month. The $y$-axis is displayed on a logarithmic scale for a better visualization.

The relative performance of `EpiEstim` with monthly sliding windows, in general, is not as good as its weekly sliding window based on the relative positions of its boxes and the counterparts of the other methods. This is likely because `EpiEstim` with longer sliding windows assumes similarity of neighbouring $\mathcal{R}_t$ across longer periods, and thus, is smoother and less accurate compared to the one with shorter sliding windows.

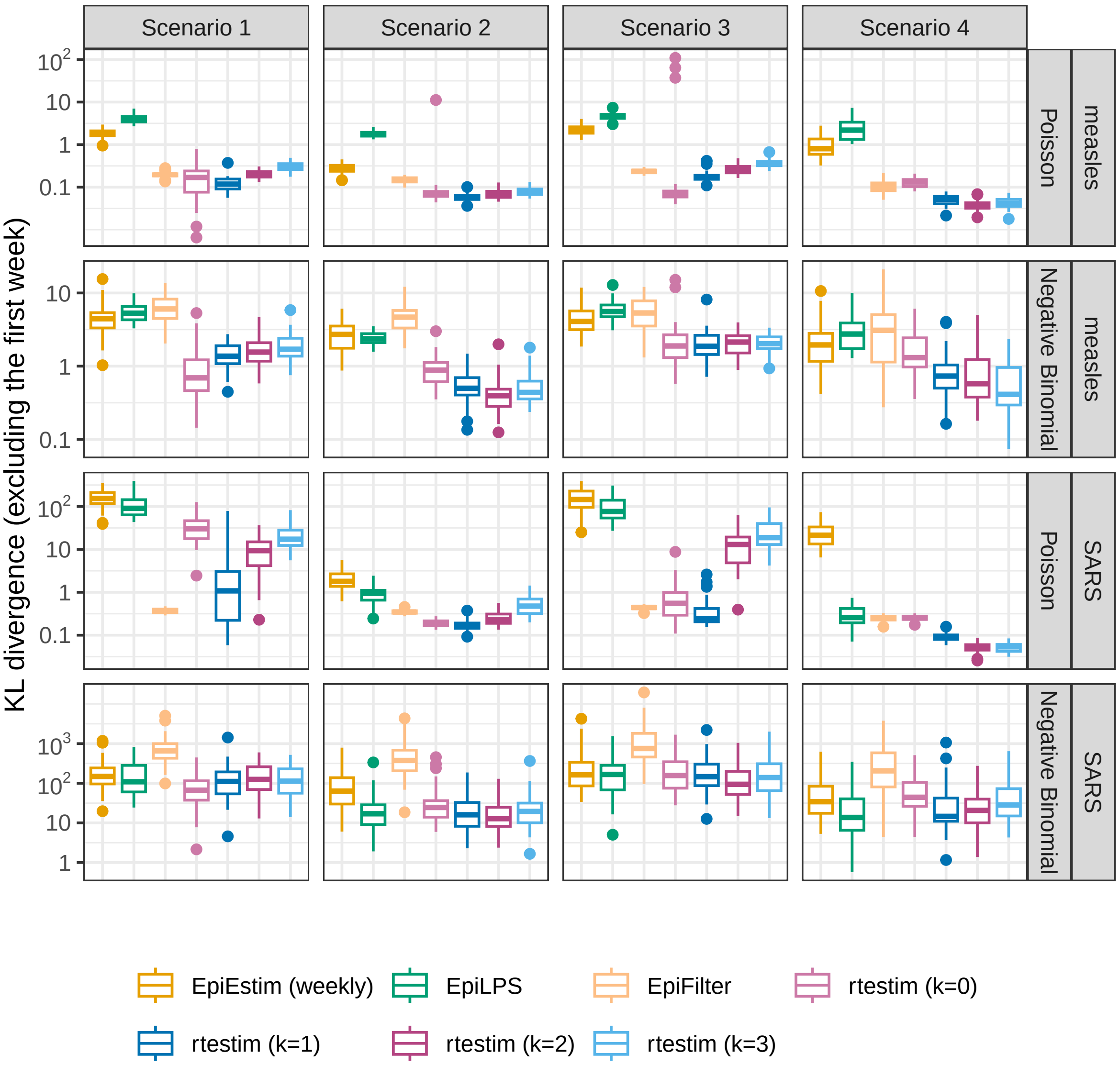

Figure A.3.1: The mean KL divergence excluding the first week for measles and SARS epidemics, since `EpiEstim`with the weekly sliding window does not provide estimates for the first week. Y-axes are on a logarithmic scale.

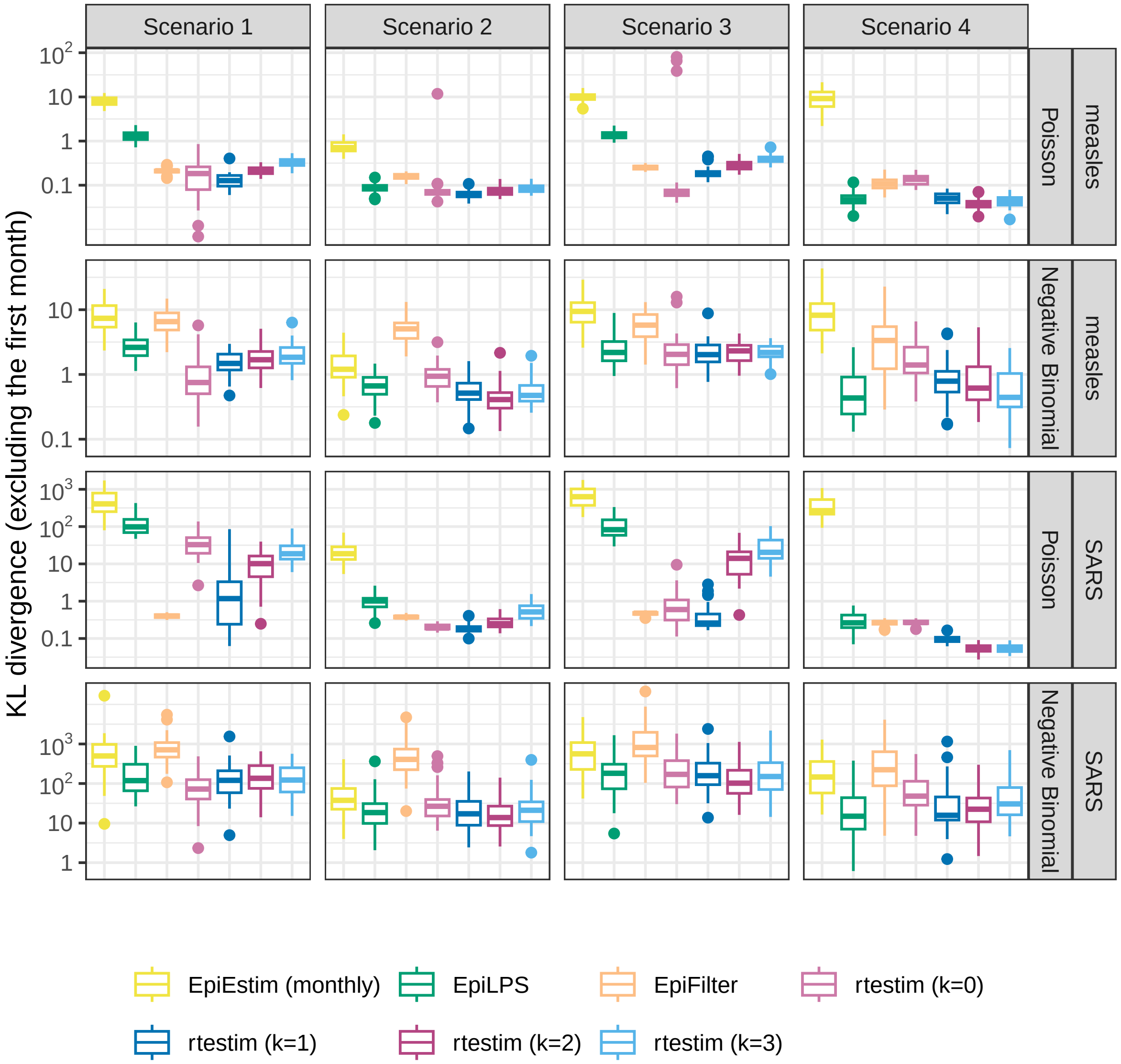

Figure A.3.2: The mean KL divergence excluding the first month for measles and SARS epidemics, since `EpiEstim`with the monthly sliding window does not provide estimates for the first month. Y-axes are on a logarithmic scale.

##### A.3.2 Short epidemics

Figures A.3.3 and A.3.4 display the KL divergence for short epidemics aggregated over time excluding the first week and month respectively to compare `EpiEstim` with weekly and monthly sliding windows with other methods including `EpiNow2`. The difference in accuracy is more obvious for Poisson incidence. To estimate true piecewise linear $\mathcal{R}_t$, piecewise constant and linear `rtestim` (with $k = 0, 1$) are the most accurate for Poisson incidence, `rtestim` ($k = 2, 3$), `EpiLPS` and `EpiFilter` are accurate as well with median KL estimates around 1. For negative binomial incidence, the advantage of `rtestim` is less obvious, but `rtestim` with all degrees still has the lowest median with a small IQR.

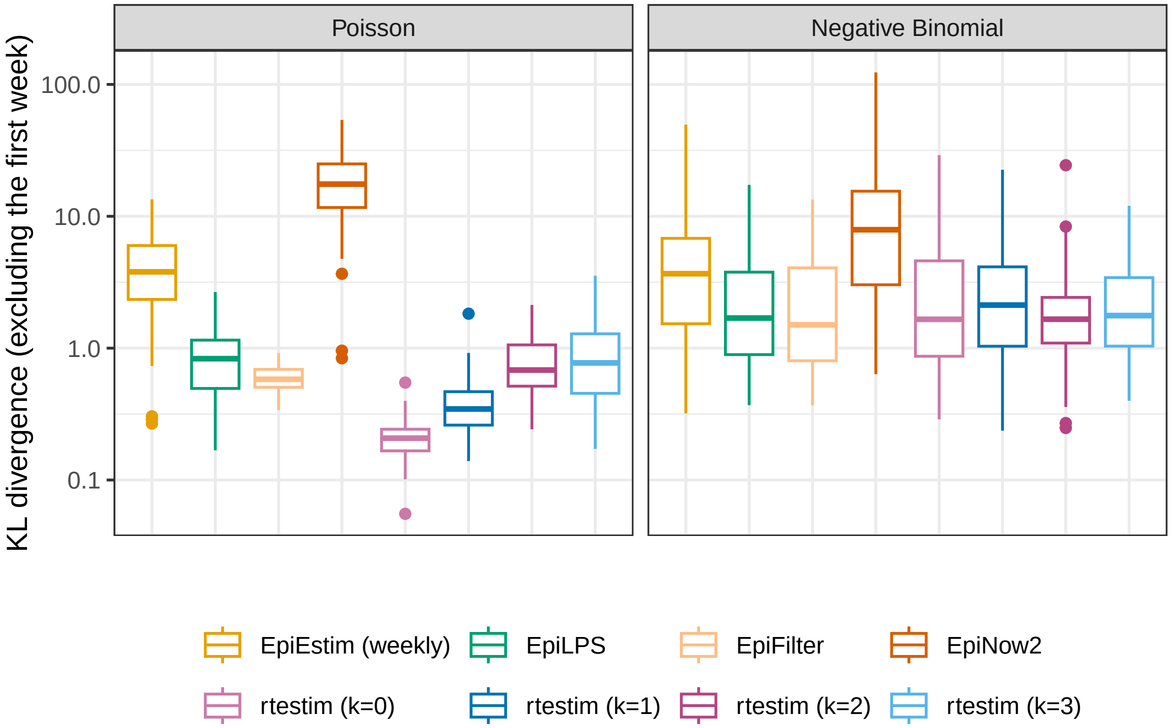

Figure A.3.3: The average KL divergence excluding the first week for flu epidemics, since EpiEstim with the weekly sliding window does not provide estimates for the first week. Y-axes are on a logarithmic scale.

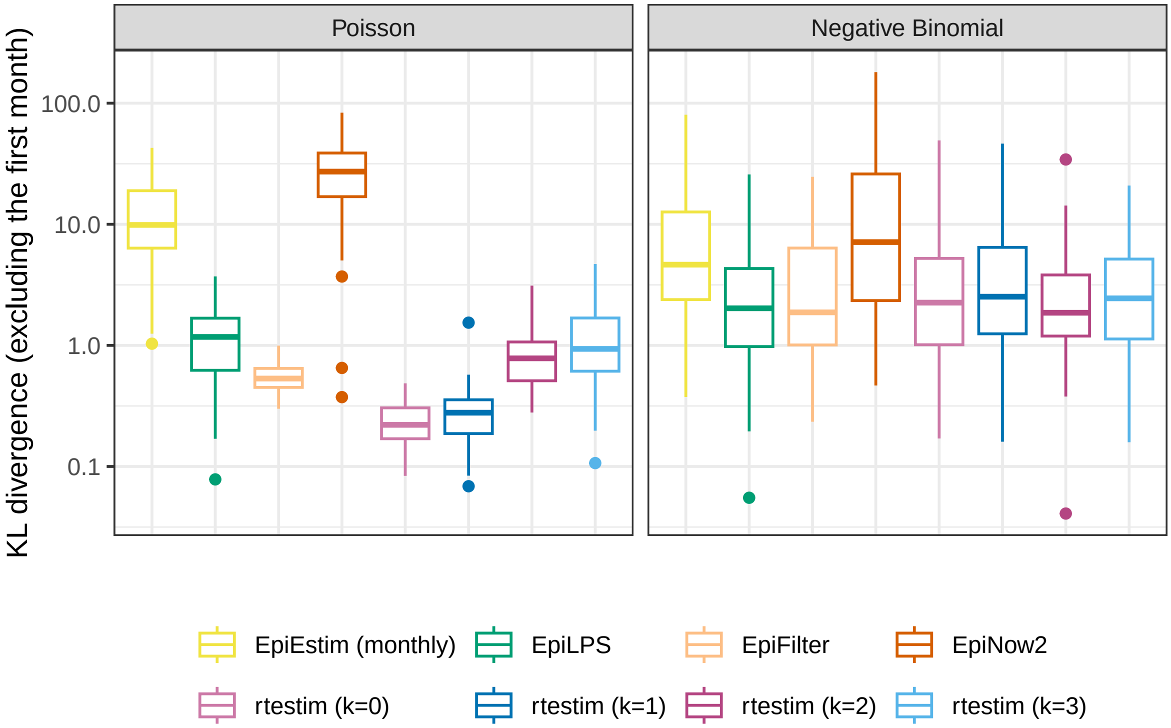

Figure A.3.4: The mean KL divergence excluding the first month for flu epidemics, since EpiEstim with the monthly sliding window does not provide estimates for the first month. Y-axes are on a logarithmic scale.

#### A.4 Experimental results under misspecification of the serial interval distributions

##### A.4.1 SI misspecification for long epidemics

Figures A.4.1 and A.4.2 display KL divergence (excluding the first week and the first month respectively) for all 8 methods with "mild" misspecification (using adjusted `measles` SI parameters) and "major" misspecification (using `SARS` SI parameters) for long `measles` epidemics across all settings. `rtestim` is reasonably robust to misspecification of the SI parameters: median KL error for each problem design is almost always the lowest with the lowest IQR. `EpiLPS` is a strong competitor under negative binomial incidence since it it uses the correct loss function. `EpiFilter` is also quite robust to SI misspecification under Poisson incidence.

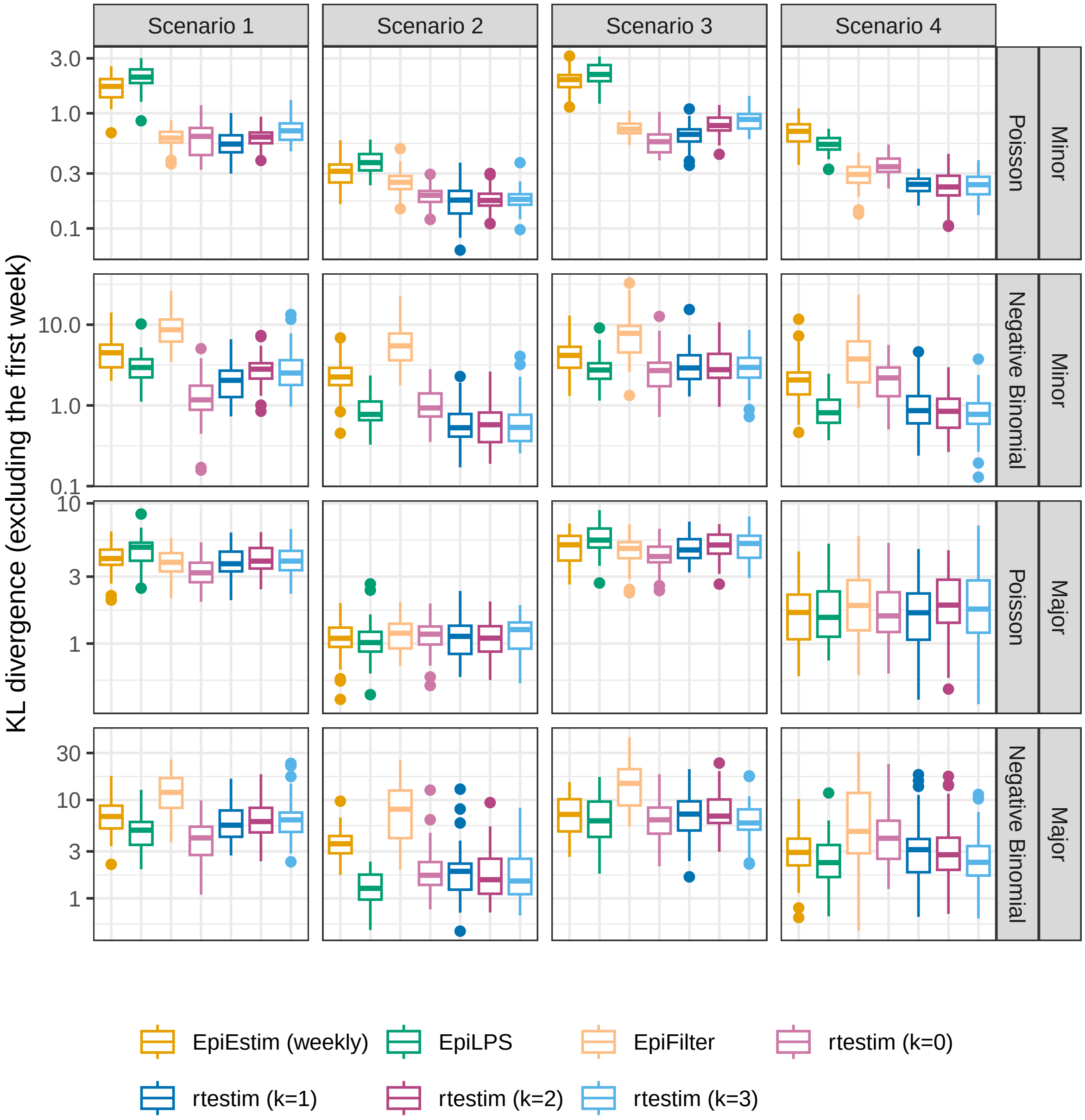

Figure A.4.1: The mean KL divergence excluding the first week for measles epidemics with SI misspecification. Y-axes are on a logarithmic scale.

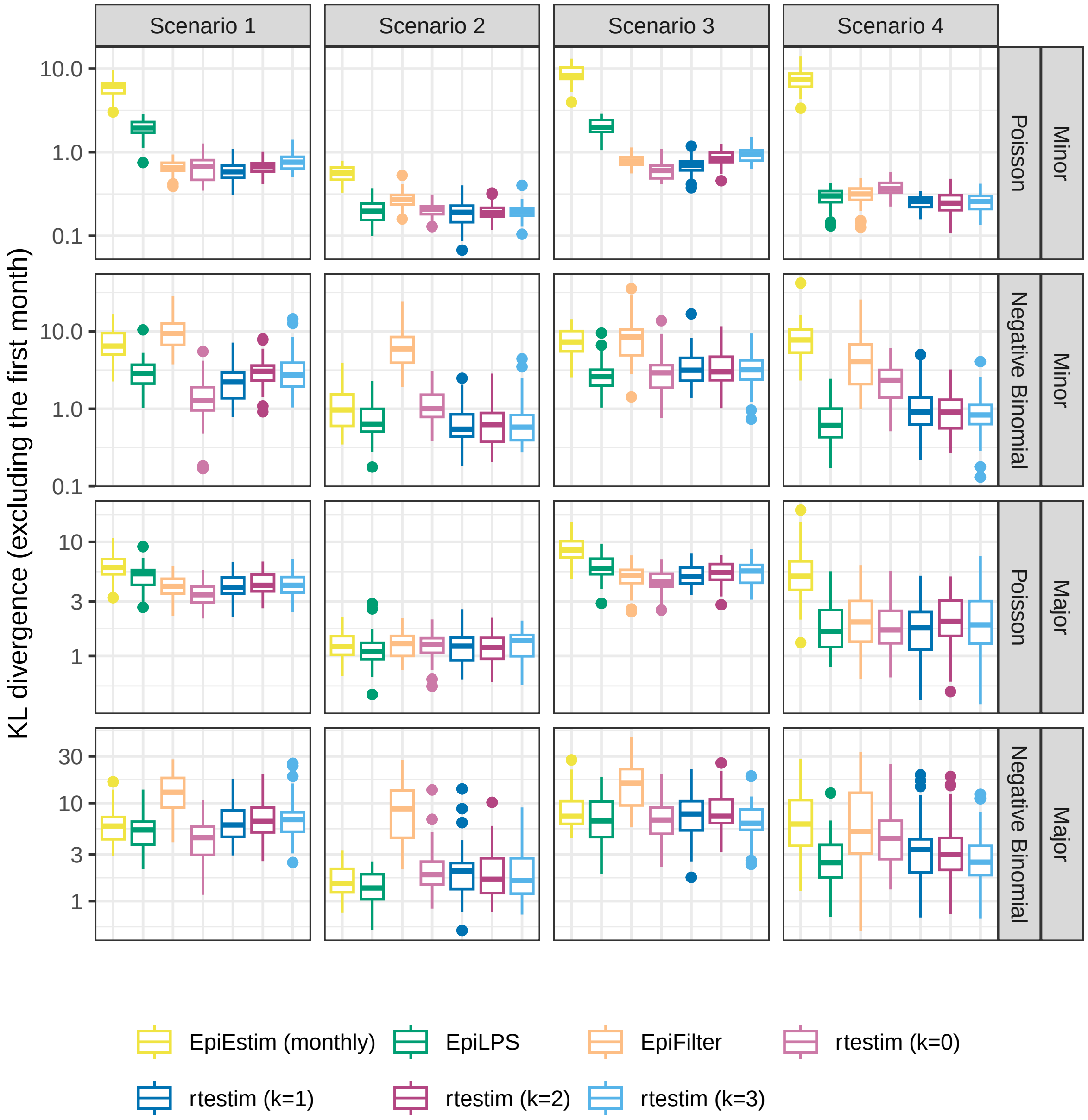

Figure A.4.2: The mean KL divergence excluding the first month for measles epidemics with SI misspecification. Y-axes are on a logarithmic scale.

##### A.4.2 SI misspecification for short epidemics

Figures A.4.3 and A.4.4 display KL divergence, excluding the first week and the first month respectively, for all 9 methods with “minor” misspecification (using slightly modified `flu` SI parameters) and “major” misspecification (using `measles` parameters) for short `flu` epidemics across all settings. Conclusions are similar to those for short epidemics. We also note that `EpiNow2` is quite robust to major misspecification in SI parameters, while `EpiLPS` is less satisfactory when only the first week is excluded. This may be due to the overly large estimates at the beginning of the epidemics persisting beyond the first week, but not the first month.

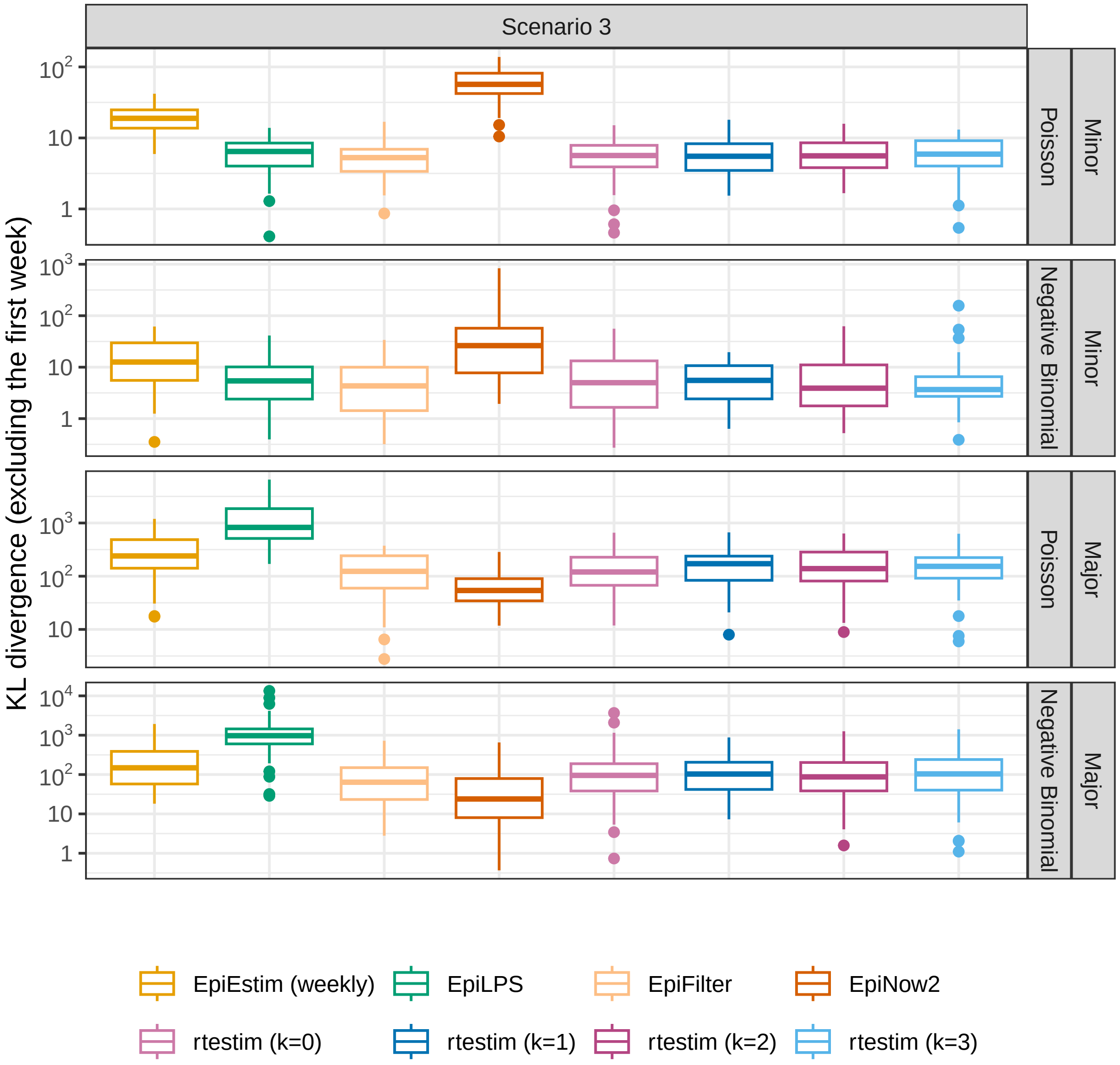

Figure A.4.3: The mean KL divergence excluding the first week for flu epidemics with SI misspecification. Y-axes are on a logarithmic scale.

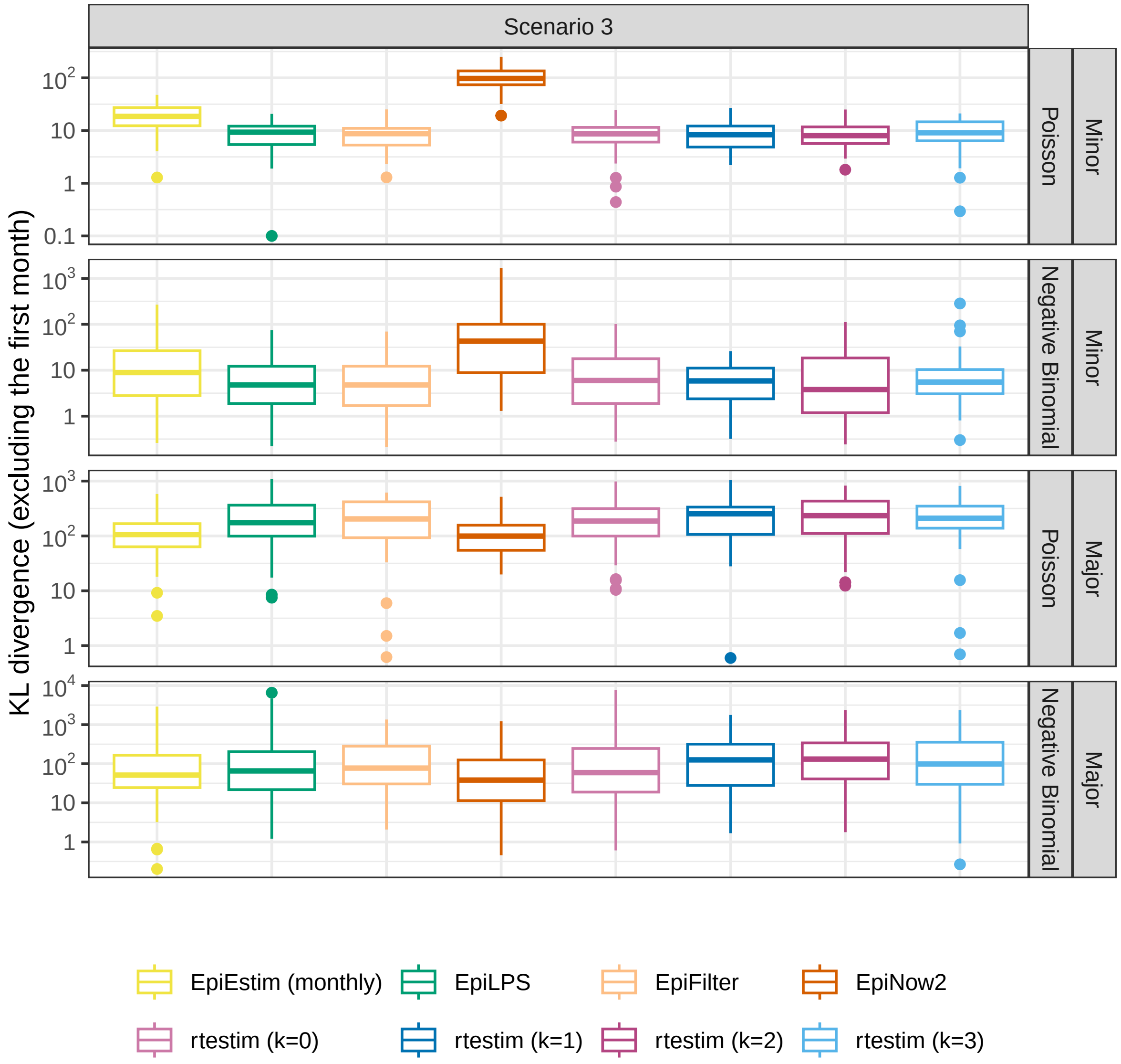

Figure A.4.4: The mean KL divergence excluding the first month for flu epidemics with SI misspecification. Y-axes are on a logarithmic scale.

#### A.5 Time comparisons of all methods

Figures A.5.1 show the time comparisons across all methods for long (`measles` and `SARS`) epidemics. `EpiEstim` with both sliding windows is very fast and converges in less than 0.1 seconds. Piecewise constant `rtestim` (with k=0) estimates can be generated within 0.1 seconds as well. `EpiLPS` is slightly slower, but still very fast and within 1 second for all experiments. `EpiFilter` is typically similar to our method with $k > 0$. Piecewise linear and cubic `rtestim` (with $k = 1$ and $k = 3$ respectively) are slower, but generally complete within 10 seconds. We also provide an alternative view with the running time of each case in a separate panel in Figures A.5.2 and A.5.3 for `measles` and `SARS` epidemics respectively. We find similar results as in Figure A.5.1.

It is remarkable that `rtestim` computes 50 lambda values with 10-fold CV for each experiment, which results in 550× the number of models estimated per experiment (including modelling for all folds). The running times are no more than 10 seconds for most of the experiments, which means the running time for each estimate is very fast: on average less than 0.02 seconds. The other methods only run once for a fixed set of hyperparameters for each experiment.

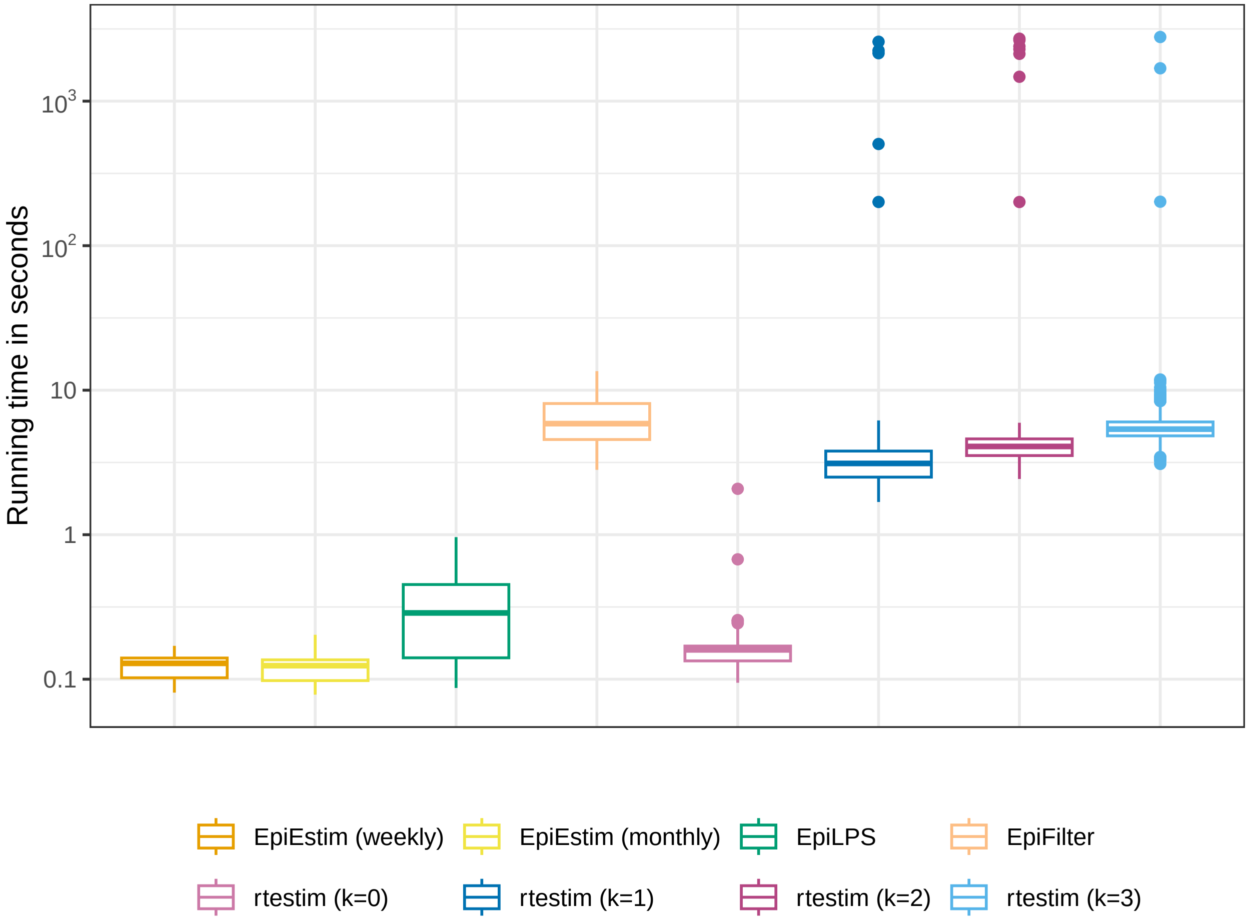

Figure A.5.1: Running time comparison of all methods for long (measles and SARS) epidemics across all cases. Y-axis is on a logarithmic scale.

Figure A.5.4 displays the running time of all methods for short (`flu`) epidemics. All methods except `EpiNow2` converge within $\sim 1$ second. Figure A.5.5 displays the running times for each setting separately, and finds

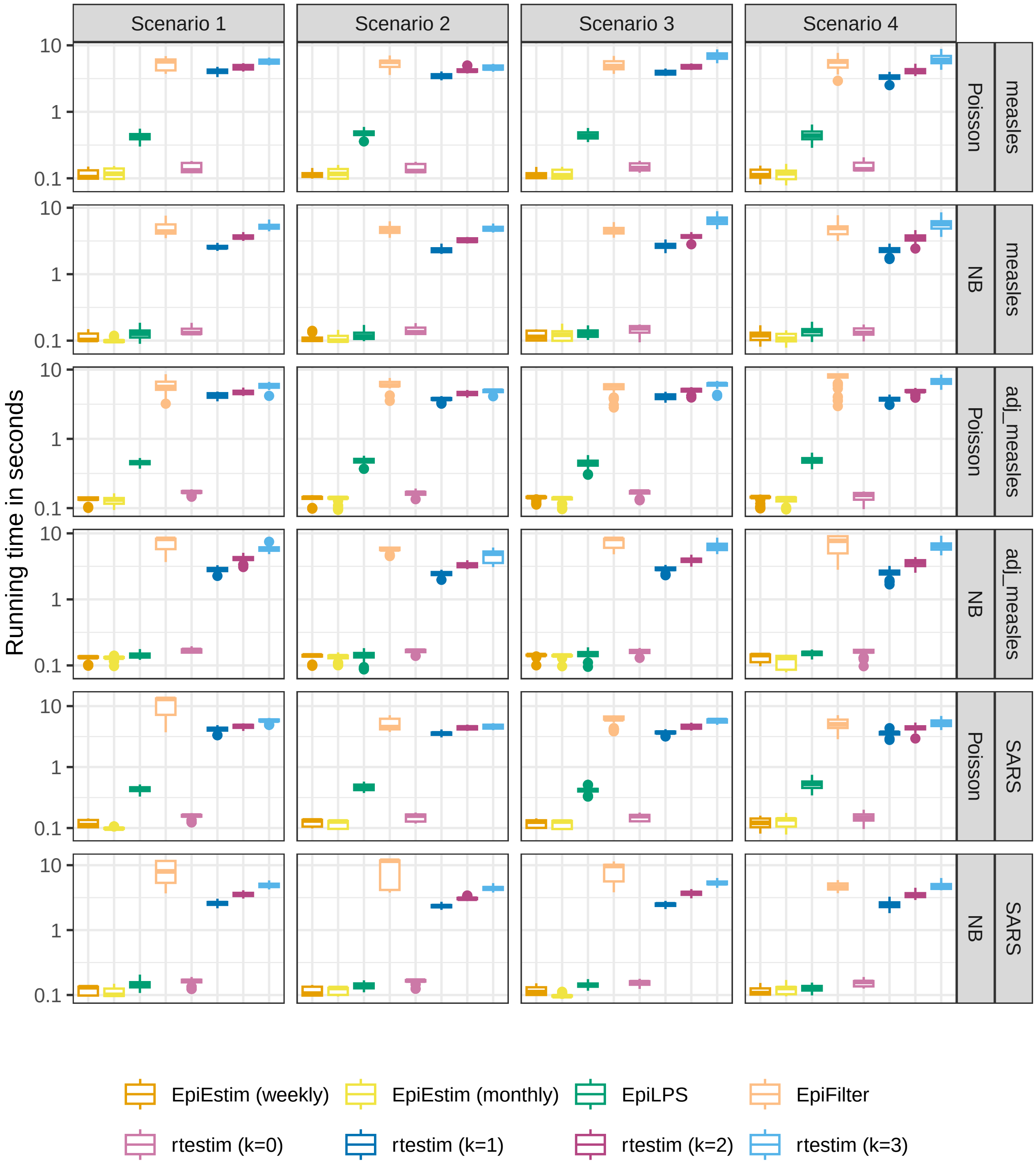

Figure A.5.2: Running time comparison of all methods for measles epidemics with each pair of SI parameters (measles, adjusted measles, and SARS; excluding outliers for better illustration). Y-axes are on a logarithmic scale.

similar results as in the overall running time comparison.

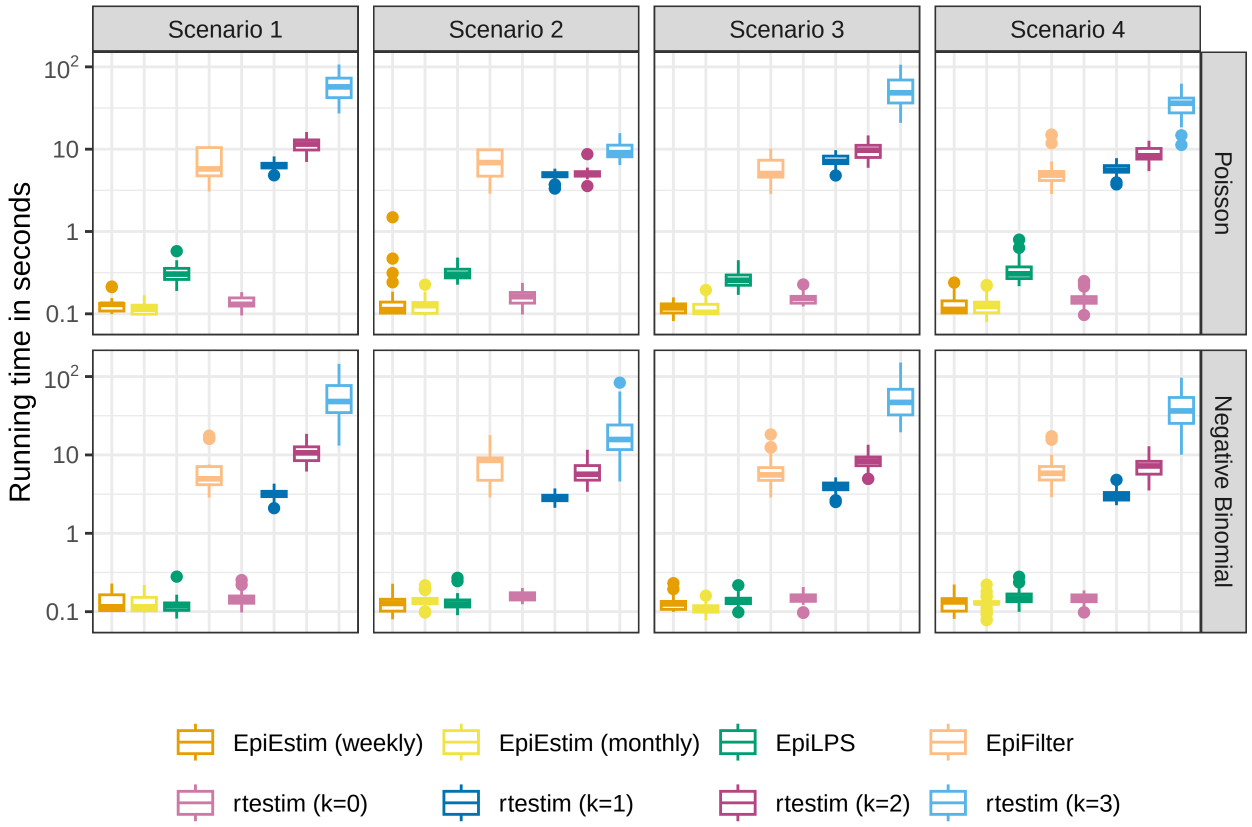

Figure A.5.3: Running time comparison of all methods for SARS epidemics with each choice of SI distribution. Y-axes are on a logarithmic scale.

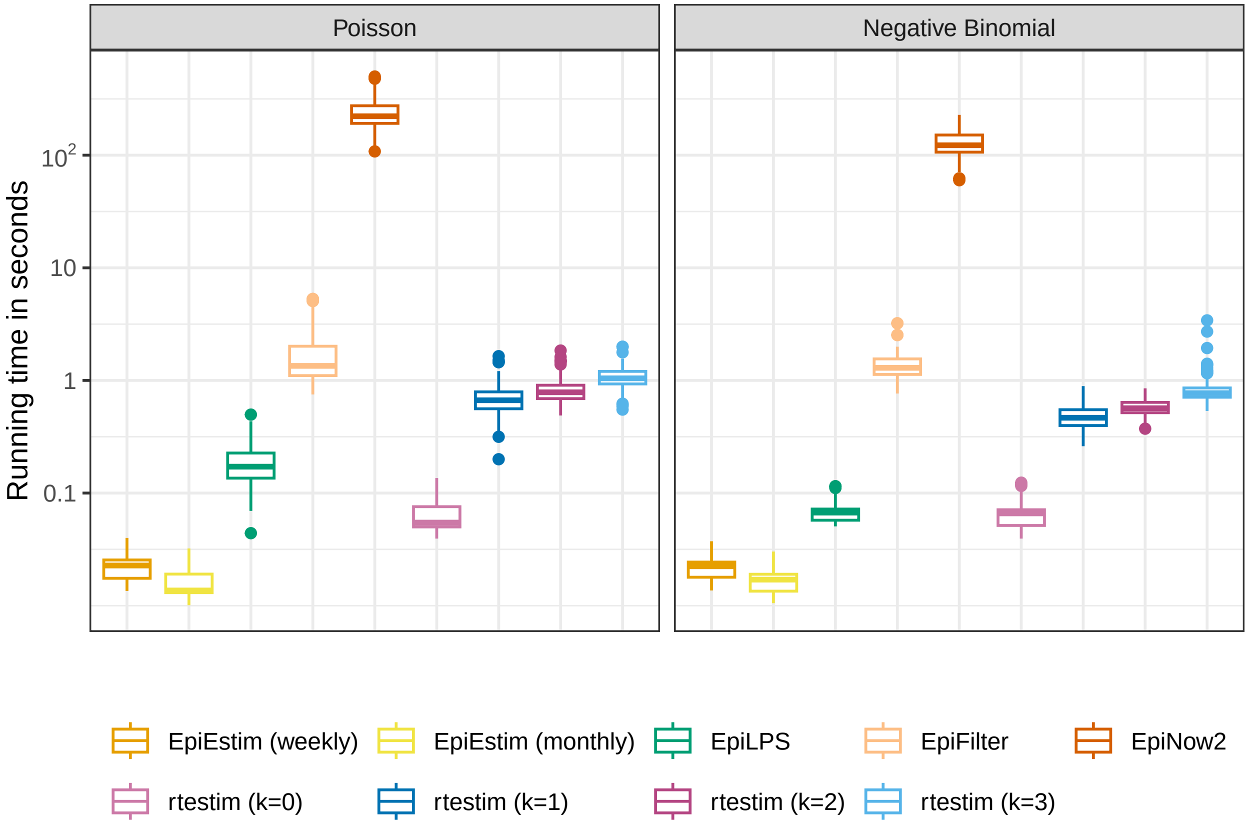

Figure A.5.4: Time comparisons of methods for short (flu) epidemics across all pairs of SI distribution. Y-axes are on a logarithmic scale.

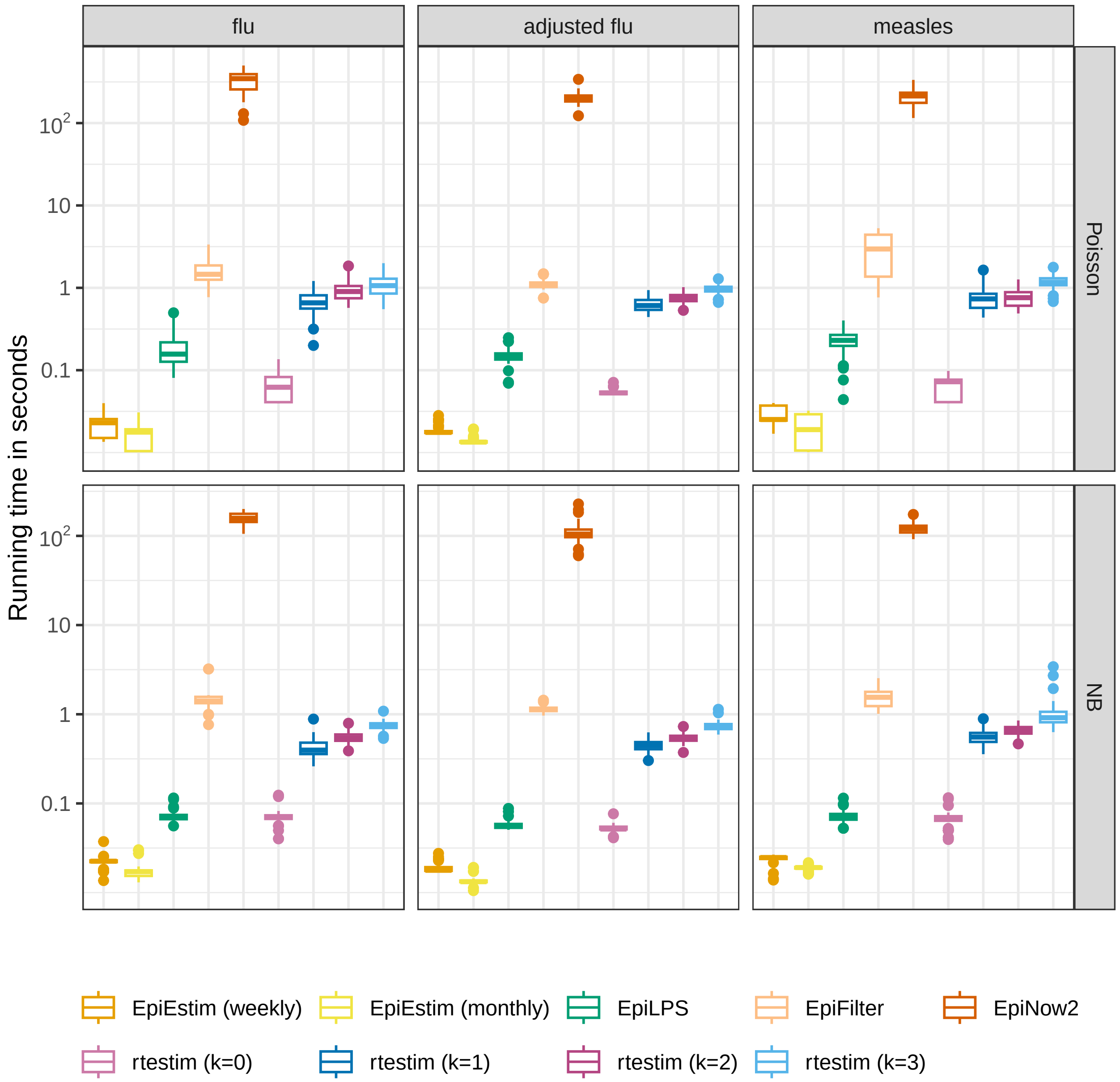

Figure A.5.5: Time comparisons of methods for short (flu) epidemics for pieciewise linear Rt (Scenario 3) for different pairs of SI parameters (flu, adjusted flu, and measles) and incidence distributions in different panels. Y-axes are on a logarithmic scale.

#### A.6 Confidence interval coverage

##### A.6.1 Estimates and confidence intervals for sample epidemics

Fig 5 and Fig 6 in the manuscript provided $\mathcal{R}_t$ estimates by all methods on sample `measles` epidemics with Poisson incidence and `SARS` epidemics with negative binomial incidence respectively. Figures A.6.1 and A.6.4 provide a clearer view of each method with its 95% confidence interval in a separate panel. The full display of sample epidemics for other settings are shown in Figures A.6.2 and A.6.3.

All methods (except `EpiEstim` with the monthly sliding window) fit the epidemics with Poisson incidence well with $\widehat{\mathcal{R}}_t$ close to the true $\mathcal{R}_t$ and 95% CI covering the true value at most timepoints. Under negative binomial incidence, `rtestim` with $k = 0$ fails to recover the curvature in $\mathcal{R}_t$, especially in the exponential and periodic scenarios. `EpiEstim` with weekly sliding windows and `EpiFilter` are more wiggly, and `EpiLPS` has wider confidence intervals given negative binomial incidence compared to Poisson incidence. For large incidence under the negative binomial distribution, `EpiFilter` is extremely wiggly, and it is difficult for `rtestim` (k=0) to recover many changepoints and the curvature especially in exponential and periodic scenarios. `EpiLPS` performs well overall, but returns large estimates at the beginning of the epidemics, estimates which remain inflated well after the first week. Overall, our method with different degrees can recover the changepoints and graphical curvature of $\mathcal{R}_t$ in all scenarios, except in the case of the periodic $\mathcal{R}_t$ curve with large negative binomial incidence, where `EpiLPS` has a clear advantage ignoring the inflated estimates at the early stage. The accuracy across different settings by different methods generally coincides with the findings in the KL divergence estimates.

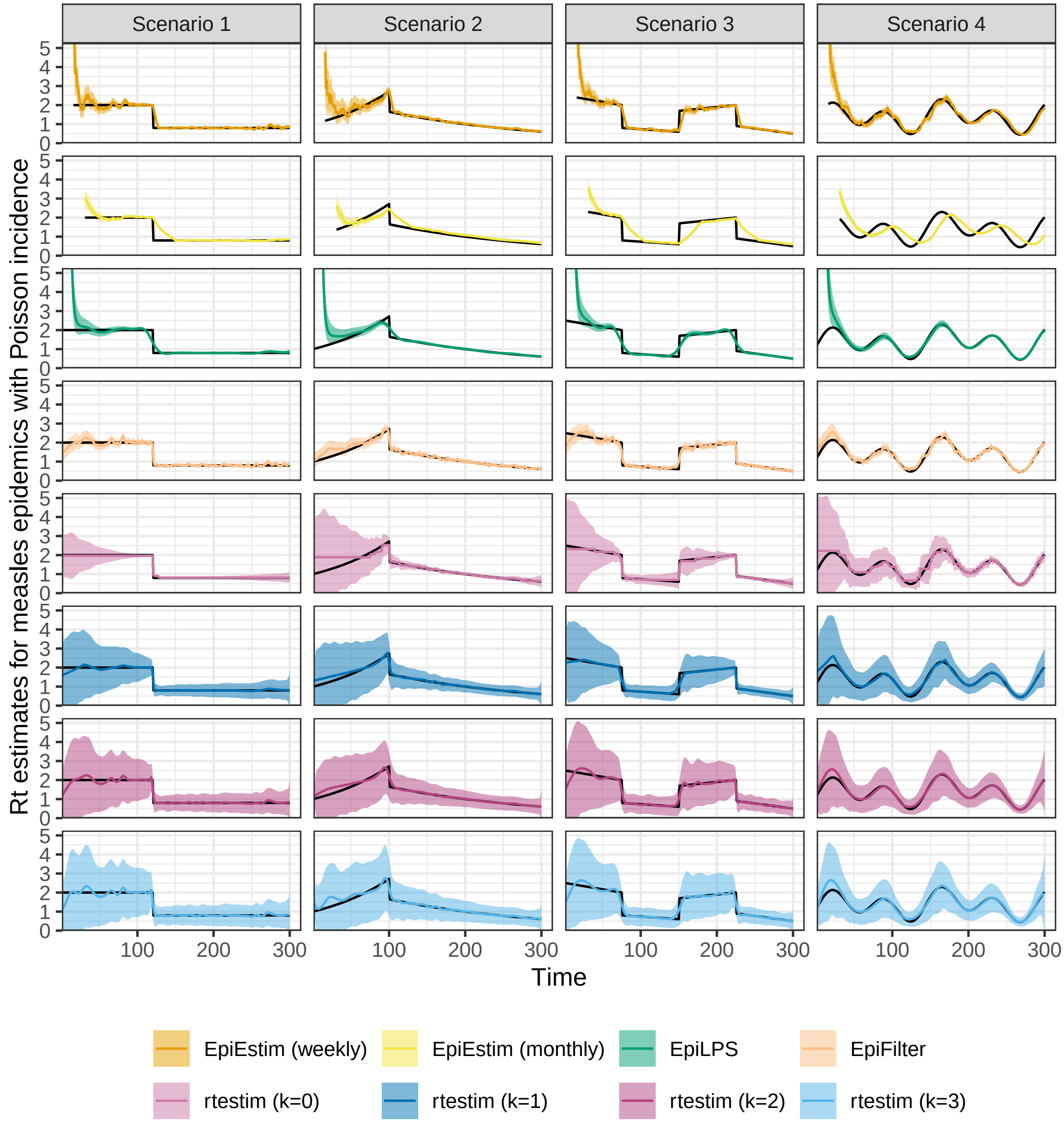

Figure A.6.1: Example measles epidemics with Poisson incidence. Y-axes beyond 5 are truncated for better illustration of small values.

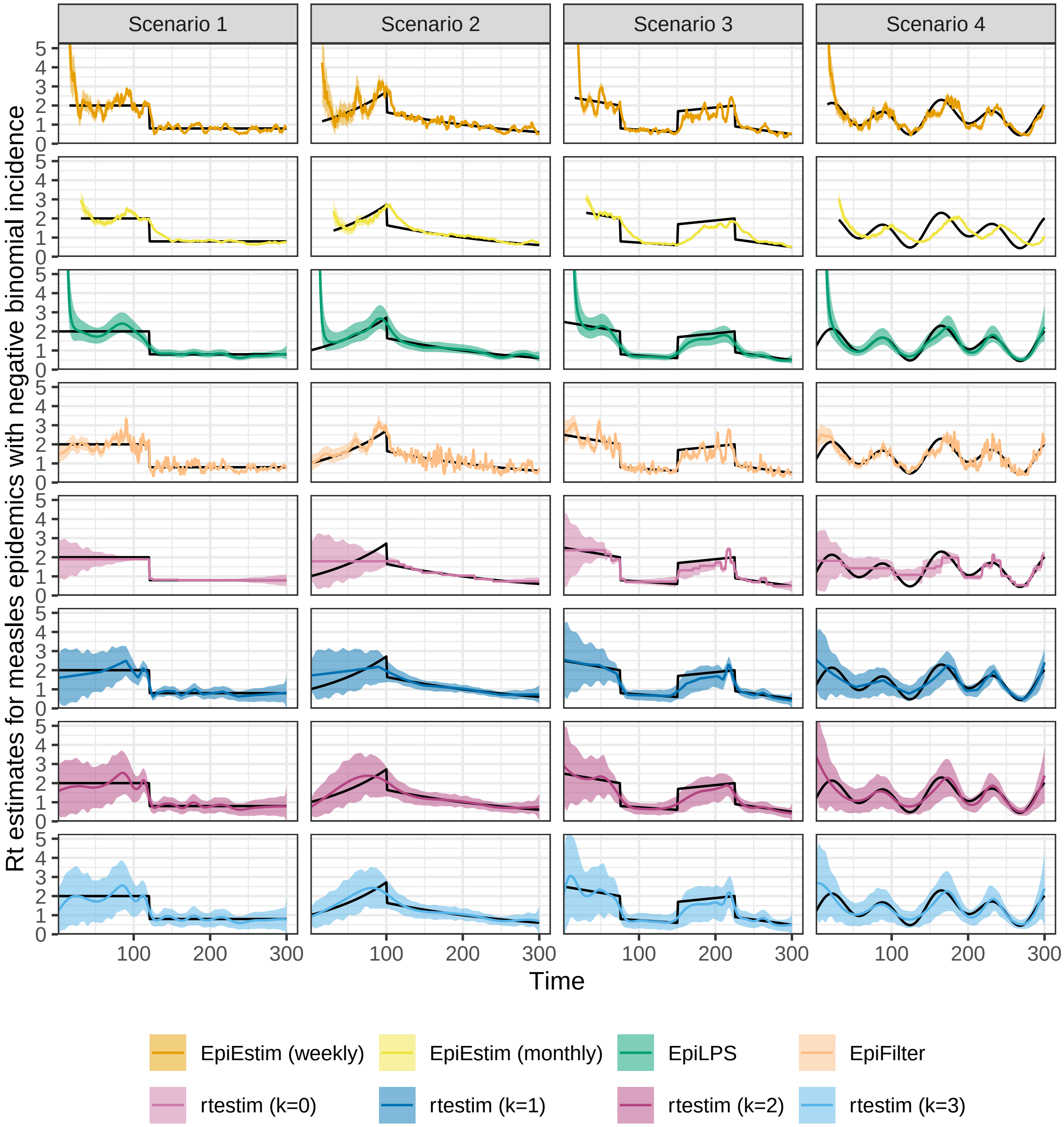

Figure A.6.2: Example measles epidemics with negative binomial incidence. Y-axes beyond 5 are truncated for a better illustration of small values.

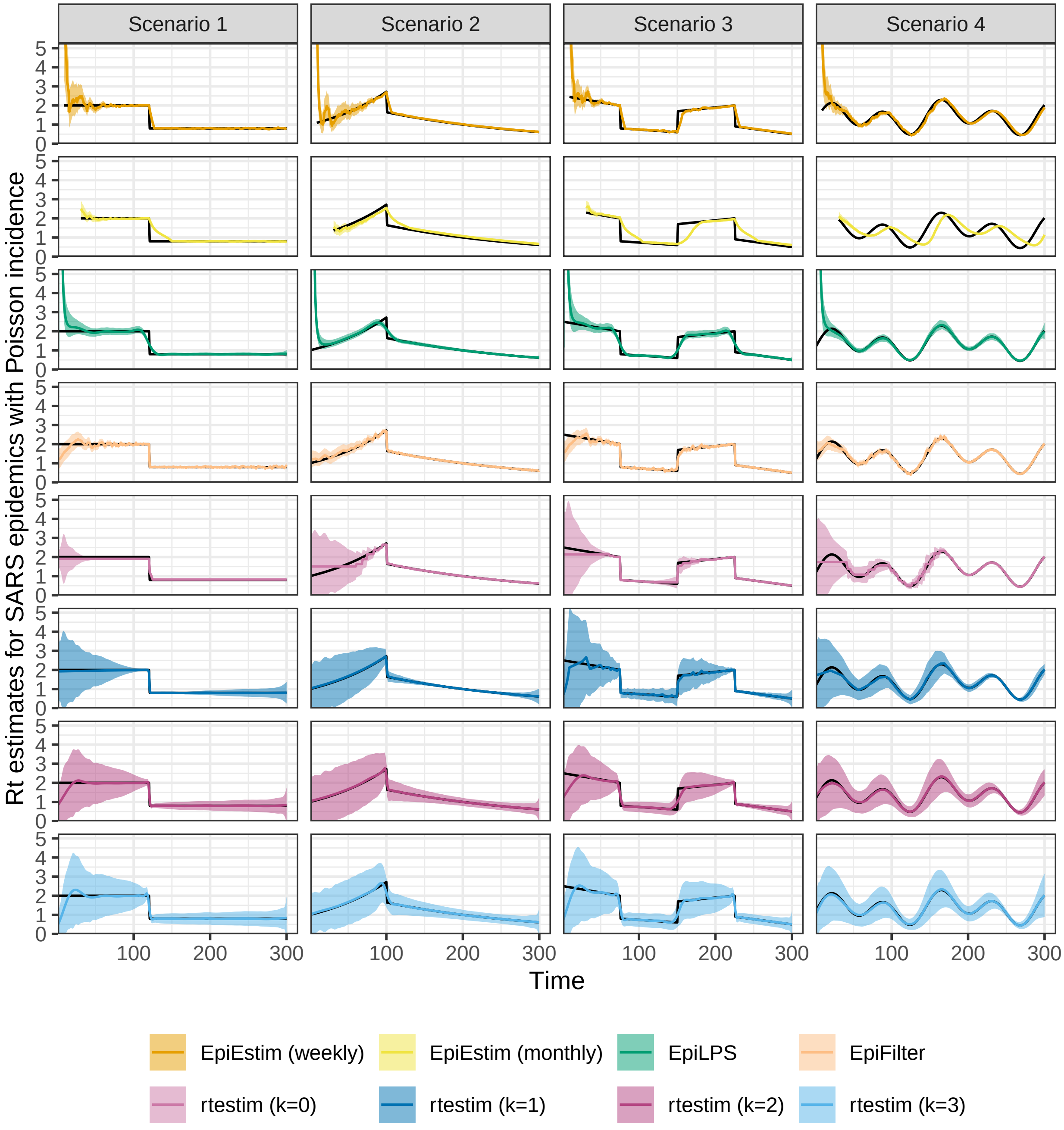

Figure A.6.3: Example SARS epidemics with Poisson incidence. Y-axes beyond 5 are truncated for a better illustration of small values.

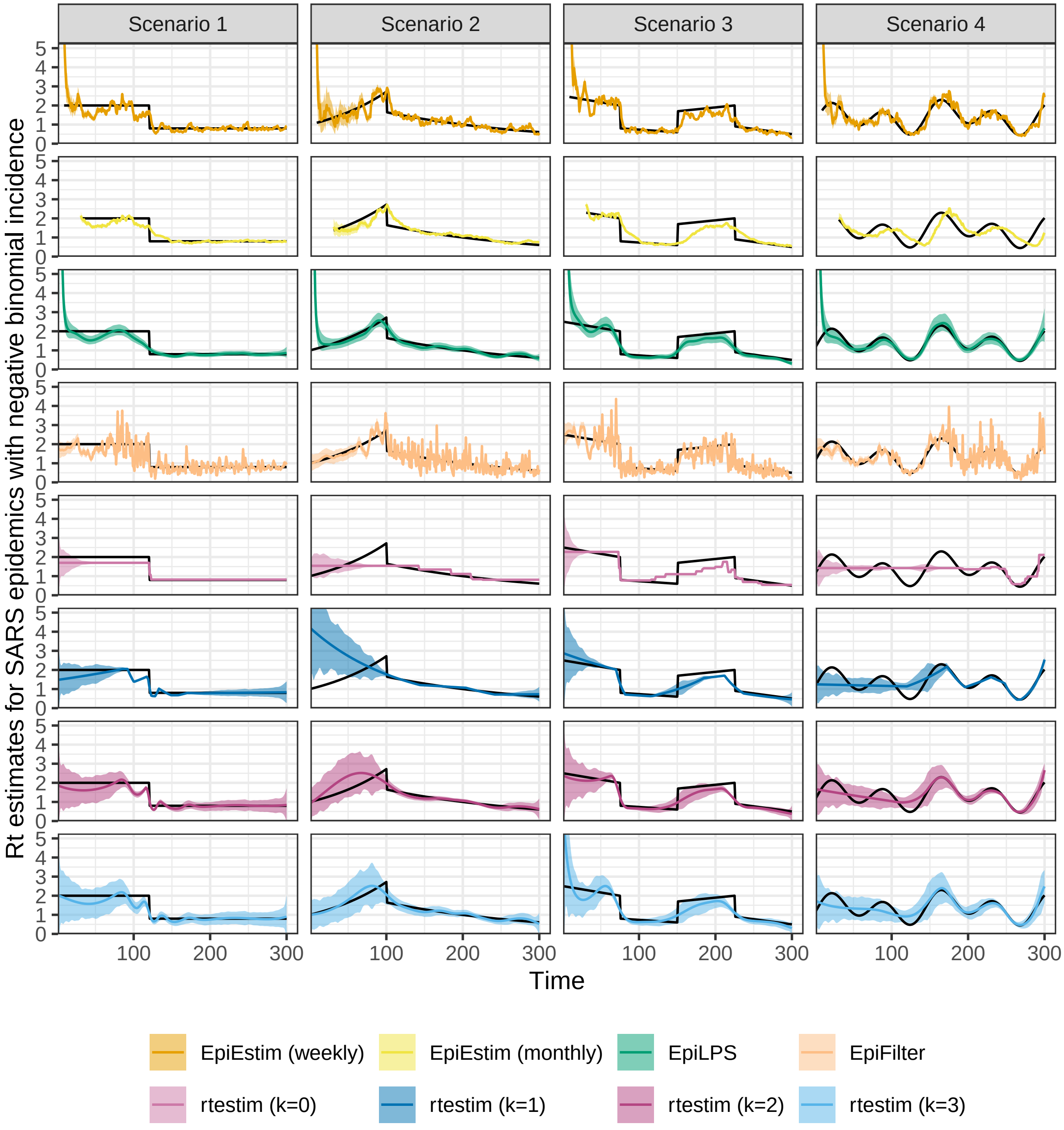

Figure A.6.4: Example SARS epidemics with negative binomial incidence. Y-axes beyond 5 are truncated for a better illustration of small values.

Table 2: Summary of experimental settings on coverage of confidence intervals

| Length | SI | Rt scenario | Incidence | SI for modelling | Method |
|---|---|---|---|---|---|
| 300 | measles | 3 | Poisson, NB | measles | 8 methods |
| 300 | SARS | 3 | Poisson, NB | SARS | 8 methods |

##### A.6.2 Coverage comparisons of confidence intervals

We focus on a specific $\mathcal{R}_t$ scenario, the piecewise linear case, and only long epidemics to compare the coverage of 95% confidence intervals across all 8 methods. We use the true serial interval distributions, those used to generate the synthetic epidemics, in this case. Table 2 summarizes the experimental settings. For each setting, we generate 50 synthetic epidemics.

We measure the coverage of 95% confidence intervals using three metrics:

1. percent coverage per coordinate (if available) across all synthetic data,
2. percent overall coverage over all available coordinates, and
3. interval score (Bracher et al. 2021) averaged over all available coordinates.

The first metric results in the percent coverage (across the 50 replicates) at each time point for each setting and method. In some cases, this is not available for every time point, for example, `EpiEstim` with a weekly sliding window does not provide estimates for the first week. The second metric aggregates across all time and replications. The third metric, interval score (Bracher et al. 2021), is defined as

$$\mathrm{IS}_\alpha(\mathcal{R}, u, l) = \frac{1}{n}\sum_{t=1}^{n}(u_t - l_t) + \frac{2}{\alpha}(l_t - \mathcal{R}_t)\mathbf{1}_{(\mathcal{R}_t < l_t)} + \frac{2}{\alpha}(\mathcal{R}_t - u_t)\mathbf{1}_{(\mathcal{R}_t > u_t)},$$

where $\alpha = 0.05$ is the significance level, $l, u$ are the lower and upper bounds and $\mathbf{1}_X$ is the indicator function of the condition $X$, taking the value 1 if $X$ and zero otherwise. A confidence band that covers the true values more frequently with shorter interval widths will have a lower interval score.

##### A.6.3 Experimental results on interval coverage

Figures A.6.5 and A.6.6 displays the percentages of coverage of 95% CIs per coordinate over 50 random samples for `measles` and `SARS` epidemics respectively. Low Poisson incidence is the easiest for all methods, with coverage near 100% at most timepoints and 0 at the change point. Large negative binomial incidence is the hardest: `EpiLPS` does the best here with averge coverage over all timepoints close to 1 (higher than nominal). This is consistent with the findings in the accuracy comparison (using KL values) and the illustration of sample epidemics in Figures A.6.1–A.6.4, where `EpiLPS` is the most accurate. Using `rtestim` with $k = 1, 2, 3$ has 100% coverage at most timepoints except the changepoints. The exception is the hardest case, where larger degrees tend to have higher percent coverge at most timepoints. On the other hand, `rtestim` with $k = 0$ tends to produce overly narrow intervals, leading to lower coverage. `EpiEstim` with weekly sliding windows fails to cover the true $\mathcal{R}_t$ more frequently under negative binomial incidence compared to Poisson, and performs worse for larger incidence. Its point estimates are quite accurate, but since its 95% confidence band is overly narrow, and the estimated curves are quite wiggly, it often fails to cover the true values. `EpiEstim` with monthly sliding windows has low percent coverage at more timepoints than other methods, especially under negative binomial noise. This is consistent with the findings in Section A.6.1, where the point estimates miss $\mathcal{R}_t$ frequently. It also has relatively narrow intervals. `EpiFilter` has lower percent coverage under negative binomial incidence than under Poisson incidence, which is consistent with its performance for point estimation and is to be expected given the misspecified data model.

Figures A.6.7 and A.6.8 display the percent coverage of 95% CIs averaged over all timepoints and 50 random replications of `measles` and `SARS` epidemics respectively. CIs of `rtestim` with $k = 1, 2, 3$ have nearly 100% coverage across all timepoints for all random samples except in the hardest problem, where the incidence is large and overdispersed. The coverage of `rtestim` $k = 0$ is lower than for other degrees, similar to the above. `EpiFilter` has better coverage under Poisson incidence compared to negative binomial incidence. `EpiEstim` with weekly sliding windows has higher coverage compared to monthly windows, while the percent coverage is less than the nominal 95% in most cases. `EpiLPS` is the closest to nominal in most cases, and even in the hardest problem, its empirical coverage is quite accurate.

Figures A.6.9 and A.6.10 display the interval scores of the 95% CIs averaged over 50 random `measles` and `SARS` epidemics respectively. Our method always has the lowest or close to the lowest interval scores. For Poisson, `EpiFilter` has the lowest interval scores, and the scores of `rtestim` are slightly higher. `EpiLPS` has very large interval scores due to its large estimates at the early stage of the epidemic. These large misses (much larger than the true values) are multiplied by $\frac{2}{\alpha}, \alpha = 0.05$ when computing the interval score, resulting in very poor performance on this metric.

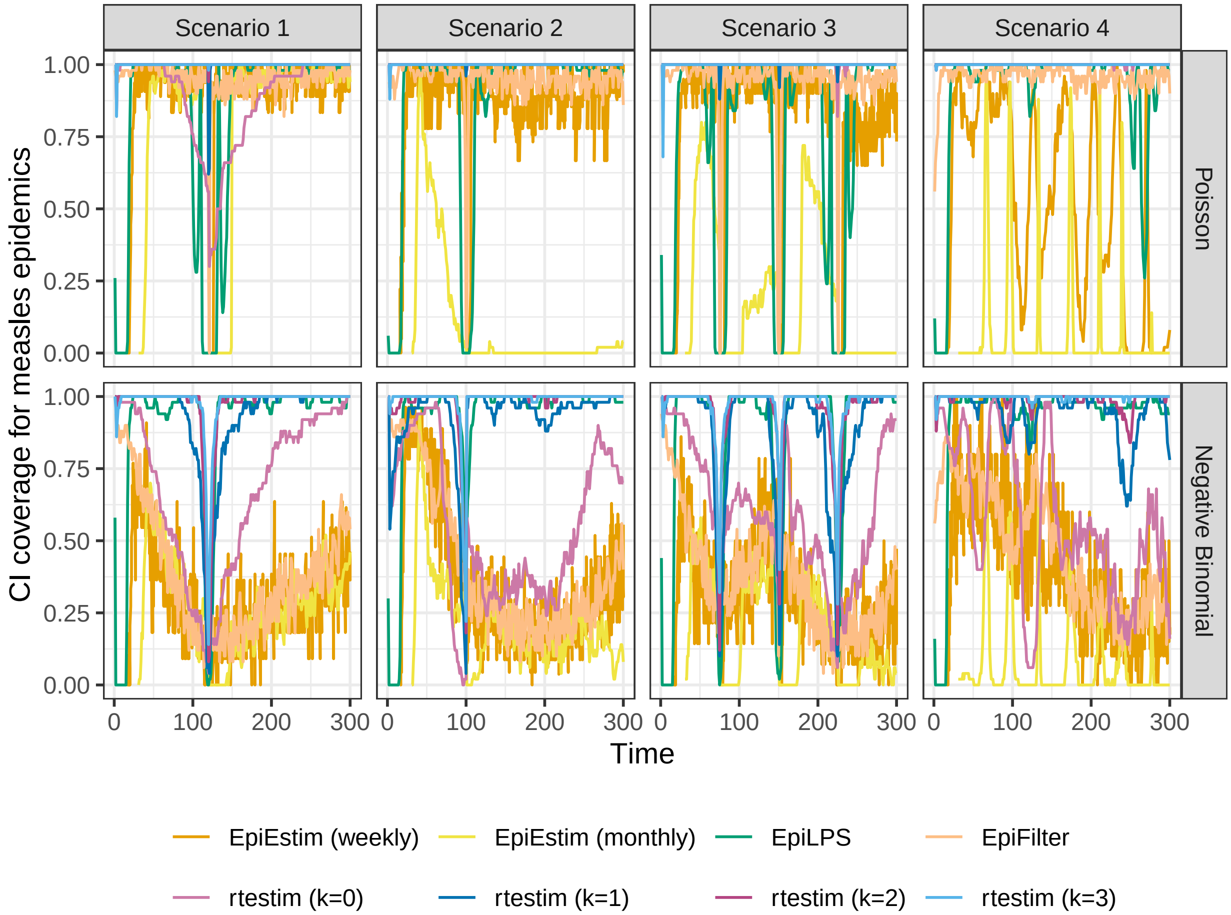

Figure A.6.5: Percent coverage of CIs per coordinate across 50 synthetic measles epidemics.

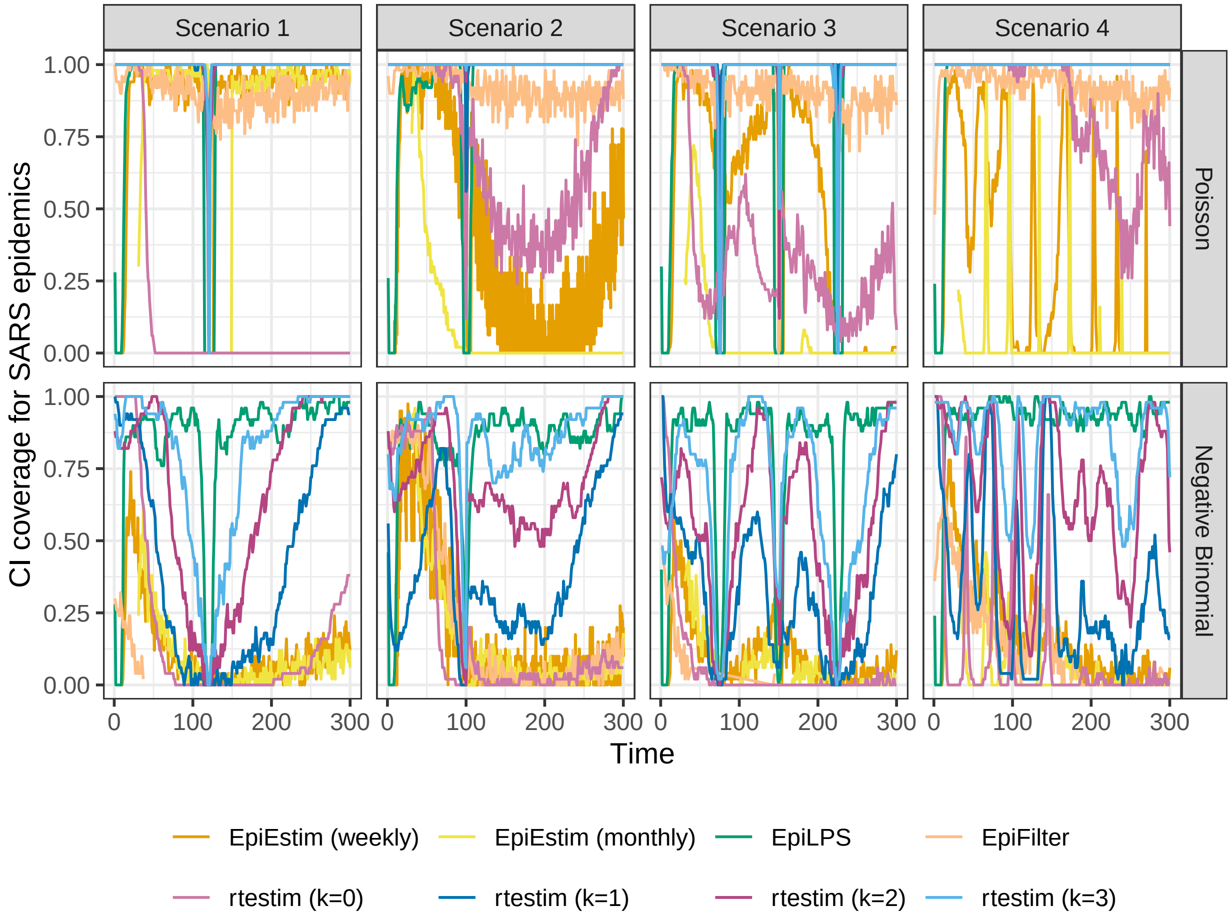

Figure A.6.6: Percent coverage of CIs per coordinate across 50 synthetic SARS epidemics.

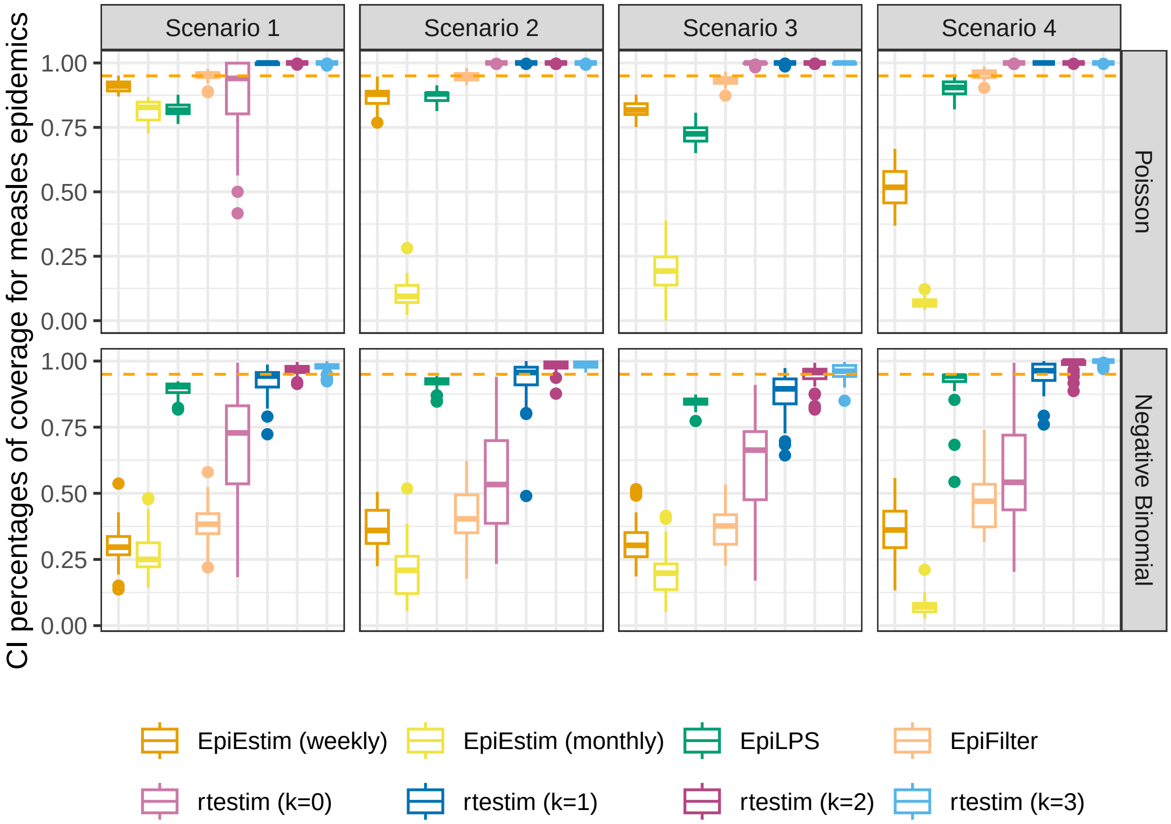

Figure A.6.7: Percent coverage of CIs over all timepoints for 50 synthetic measles epidemics. The orange dashed line represents 95% percentage of coverage across all timepoints.

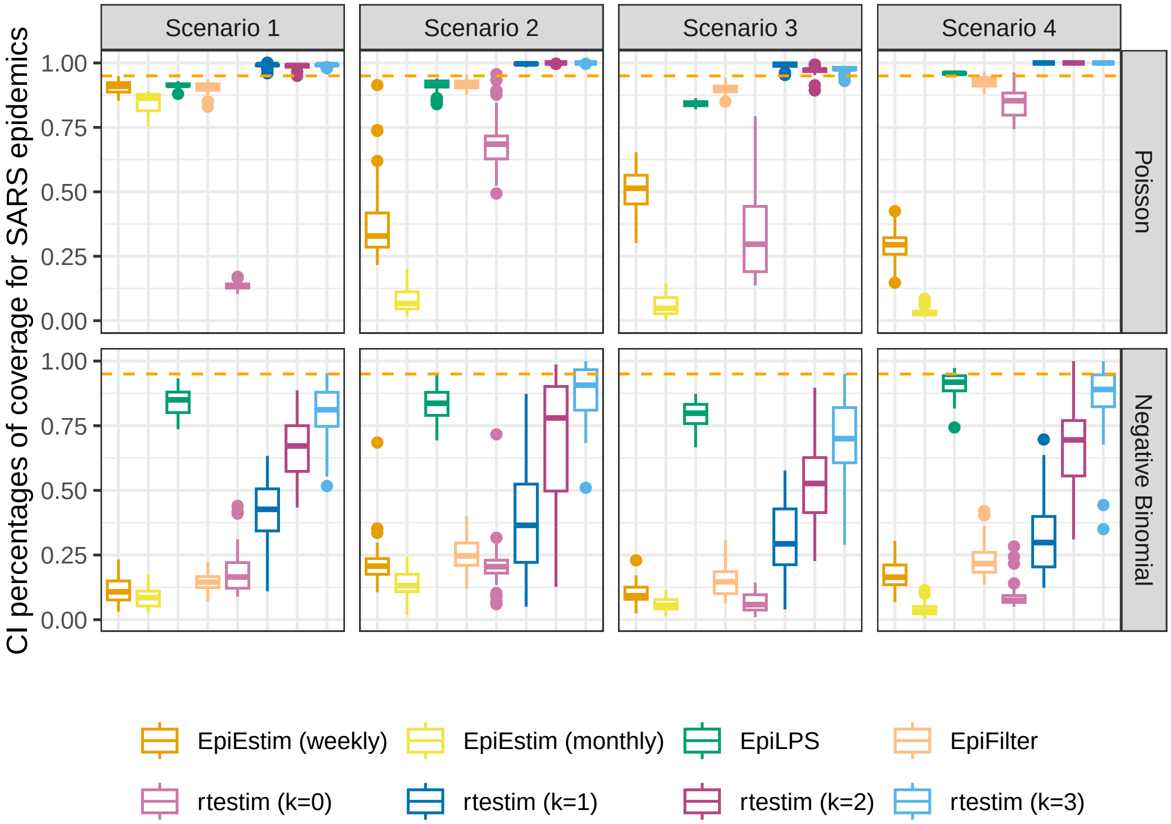

Figure A.6.8: Percent coverage of CIs over all timepoints for 50 synthetic SARS epidemics. The orange dashed line represents 95% percentage of coverage across all timepoints.

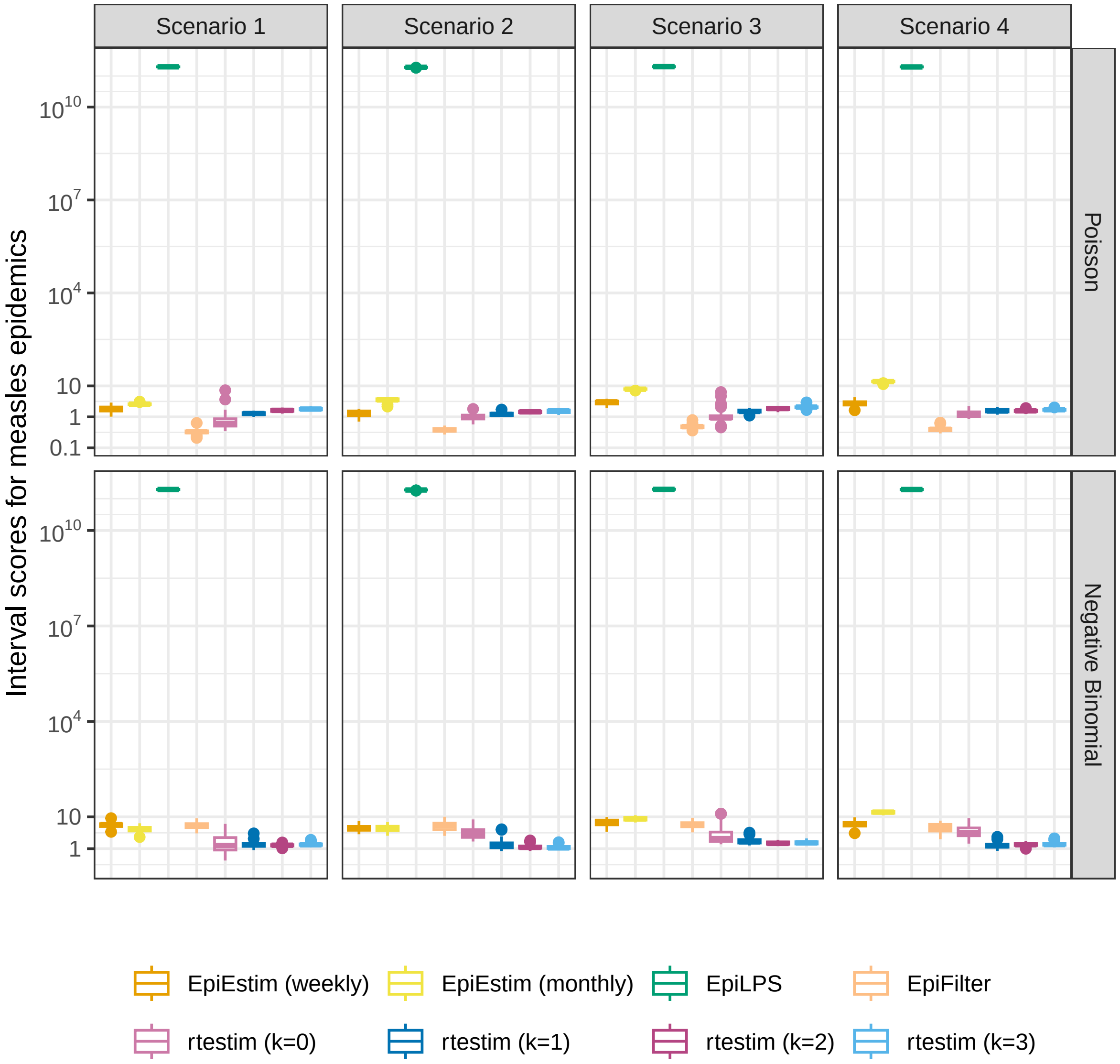

Figure A.6.9: Interval scores averaged over all coordinates for 50 synthetic measles epidemics.

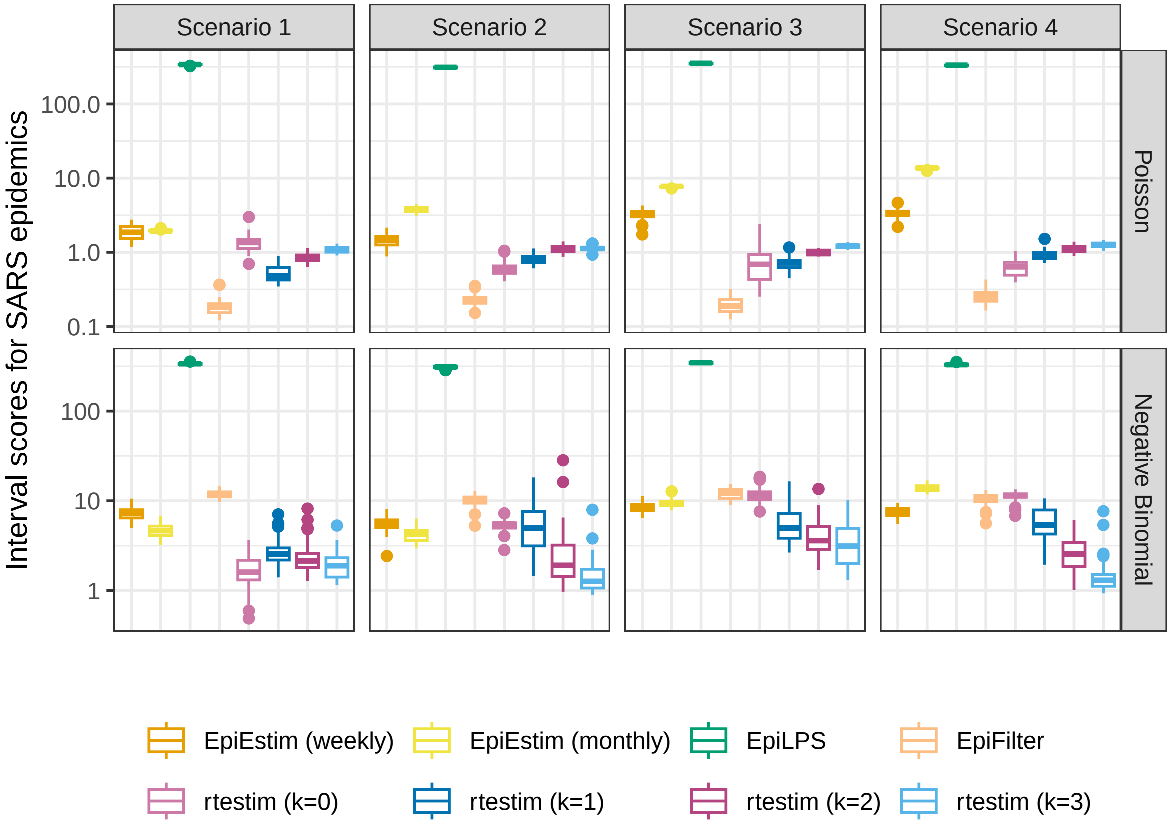

Figure A.6.10: Interval scores averaged over all coordinates for 50 synthetic SARS epidemics.

#### A.7 Data examples and alternative visualizations of Figs 5 and 6

##### A.7.1 More visualization of example epidemics

We generate `measles` and `SARS` epidemics using Poisson and negative binomial incidence distributions for each experimental setting. The condensed display of estimates for `measles` with Poisson incidence and `SARS` with negative binomial incidence are provided in Fig 5 and Fig 6 in the manuscript. A full visualization of each case is provided in Section A.6.1. Here, we provide the condensed visualization of the other cases in Figures A.7.1 and A.7.2. All methods provide accurate point estimates given large incidence from the Poisson distribution, while `EpiEstim` (with weekly sliding window) and `EpiFilter` are more wiggly under negative binomial incidence.

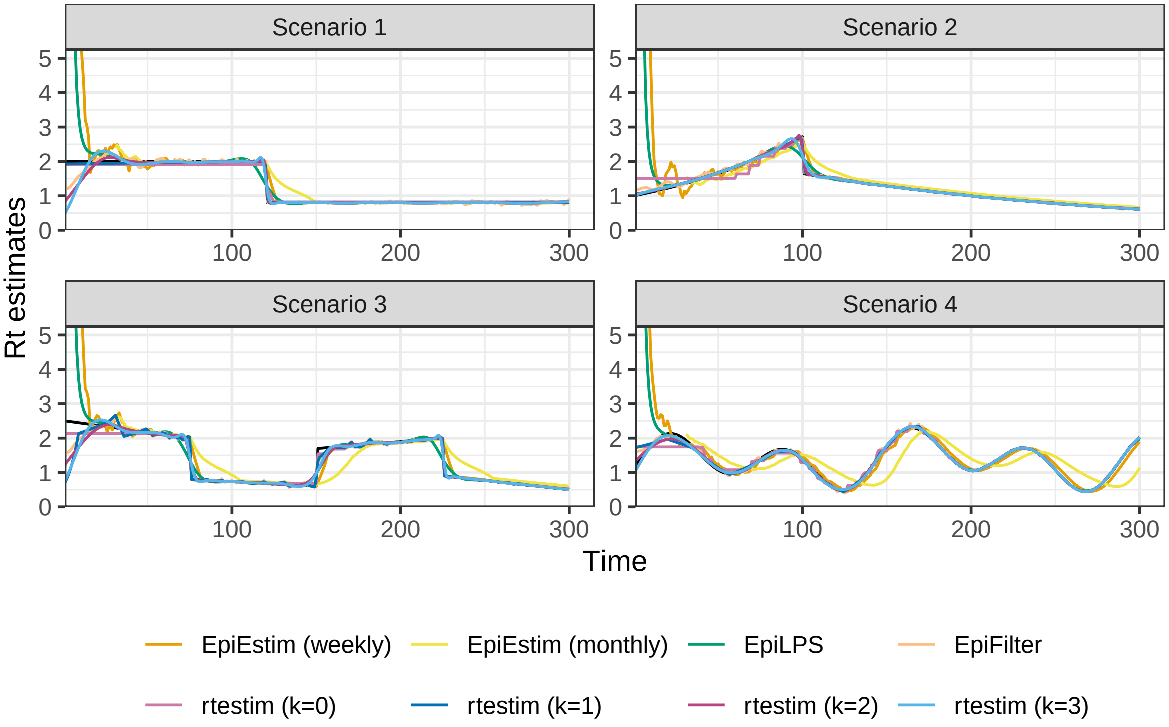

Figure A.7.1: Example of instantaneous reproduction number estimates for SARS epidemics with Poisson observations. Y-axes beyond 5 are truncated for a better illustration of small values.

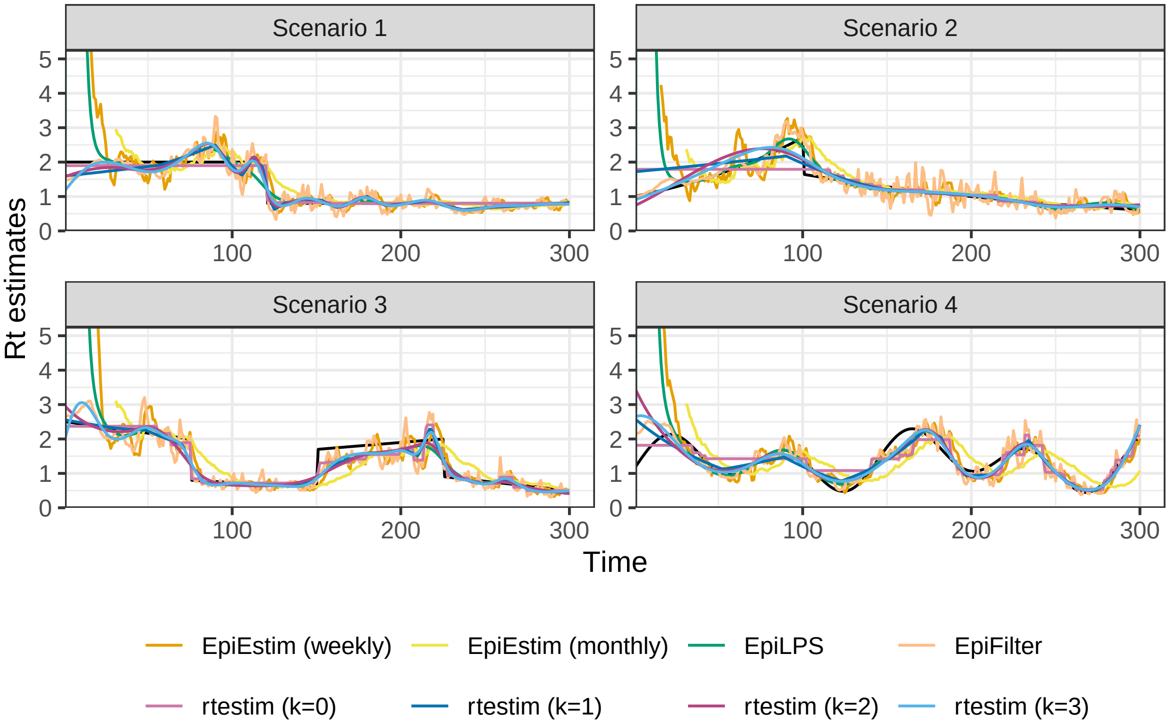

Figure A.7.2: Example of instantaneous reproduction number estimates for measles epidemics with negative binomial observations. Y-axes beyond 5 are truncated for a better illustration of small values.

##### A.7.2 Alternative view of the difference between fitted and true Rt estimates

We also provide an alternative view of Fig 5 & Fig 6 in the manuscript by plotting $\mathcal{R}_t - \widehat{\mathcal{R}}_t$ in Figures A.7.3 and A.7.4 respectively. Figures A.7.5 and A.7.6 provide the alternative view of A.7.1 and A.7.2 respectively. As is to be expected, the difference is largest at the changepoints for most methods. In the sinusoidal periodic scenario, the difference also displays a periodic pattern. This makes sense since $\mathcal{R}_t$ is sinusoidal, while most methods estimate curves to a fixed polynomial degree. Thus higher-order behaviour is missed.

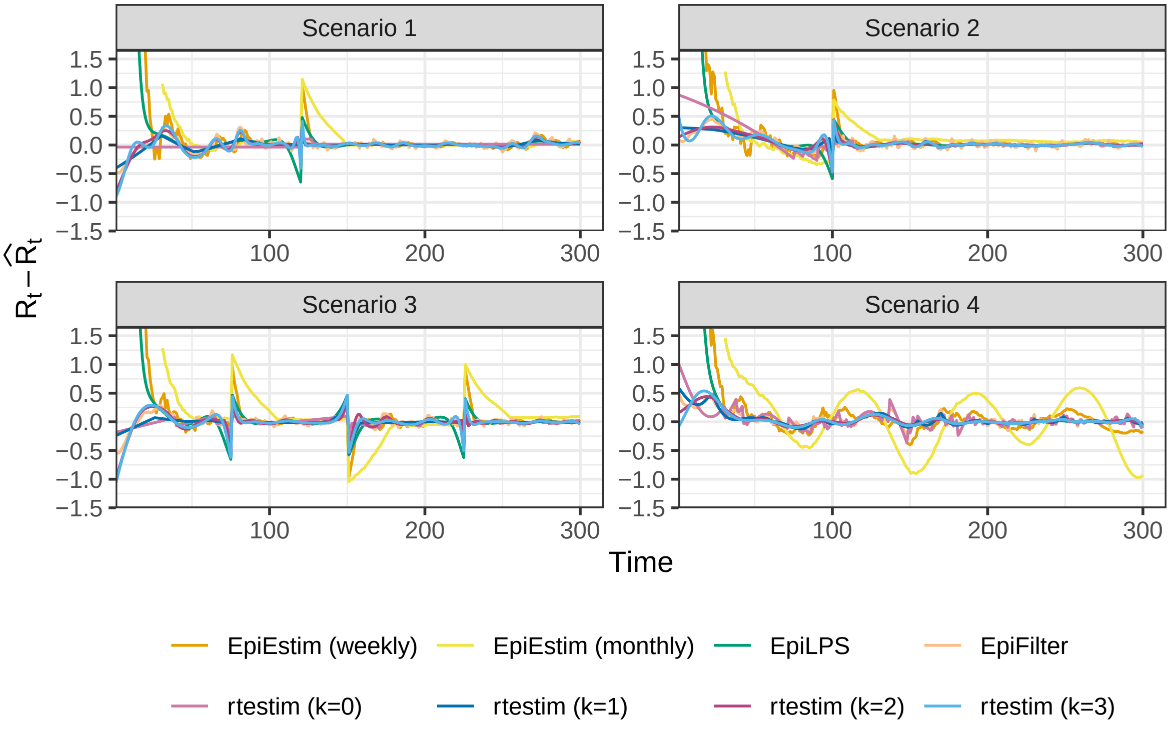

Figure A.7.3: Difference between the true and estimated instantaneous reproduction numbers for measles epidemics with Poisson observations. Y-axes beyond 1.5 are truncated for a better illustration of small values.

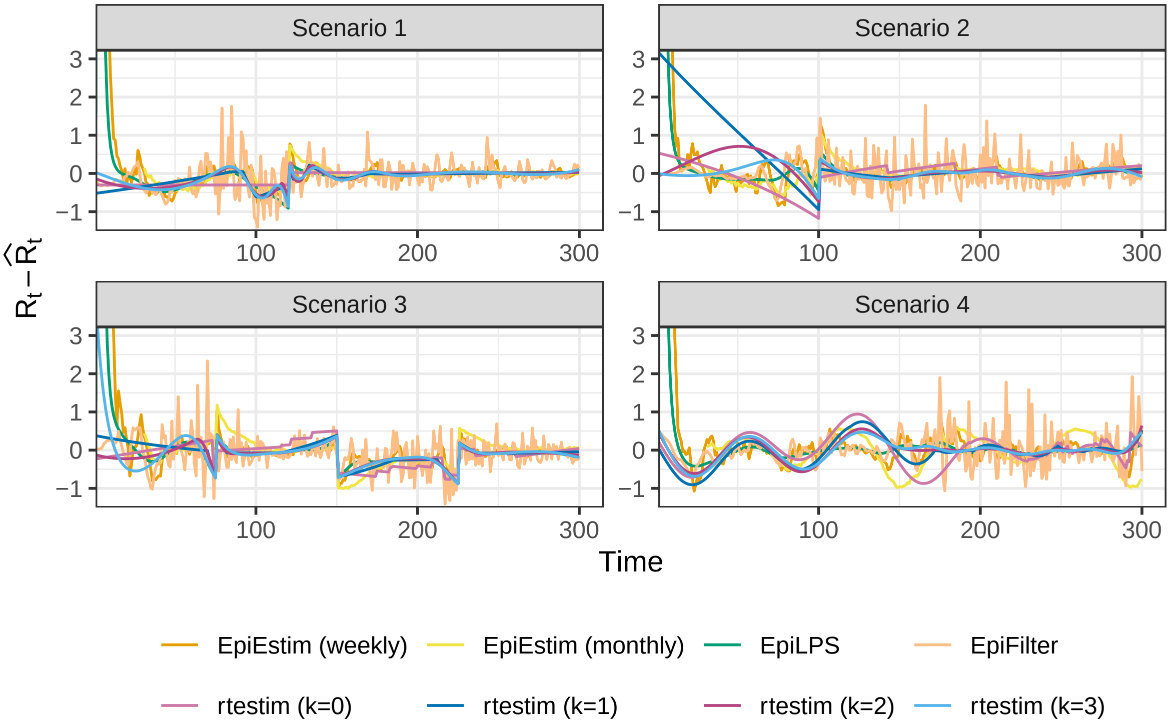

Figure A.7.4: Difference between the true instantaneous reproduction number and its estimation for SARS epidemics with negative binomial observations. Y-axes beyond 3 are truncated for a better illustration of small values.

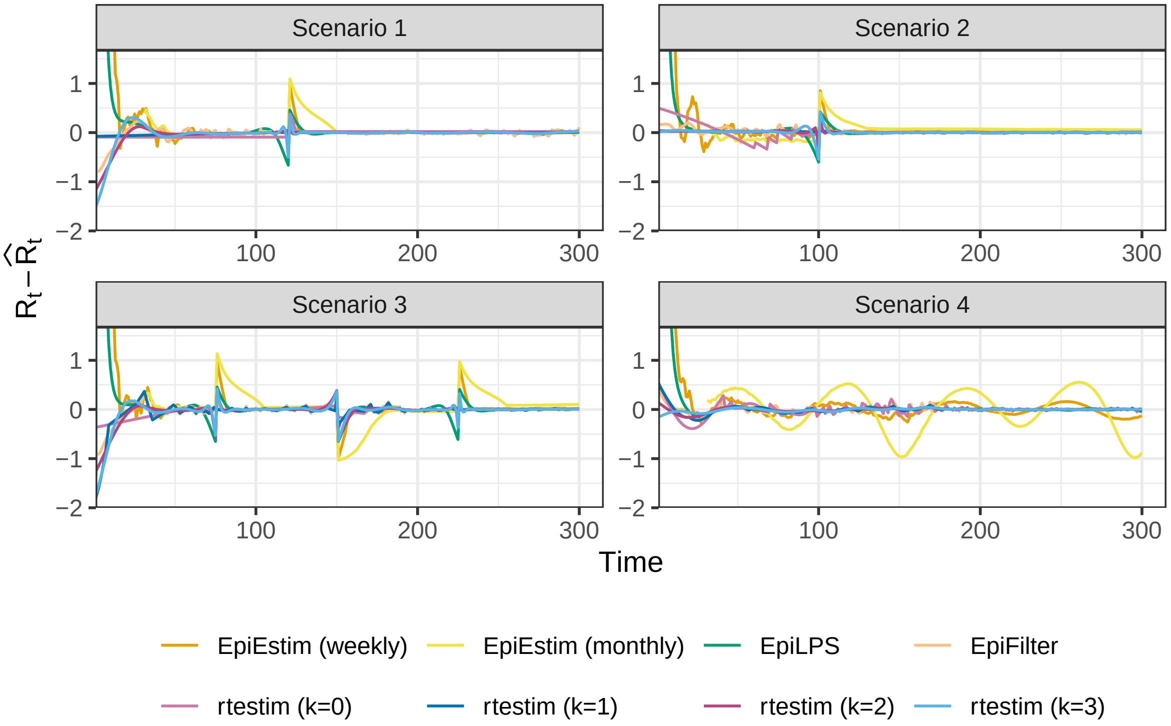

Figure A.7.5: Difference between the true and estimated instantaneous reproduction numbers for SARS epidemics with Poisson observations. Y-axes beyond 1.5 are truncated for a better illustration of small values.

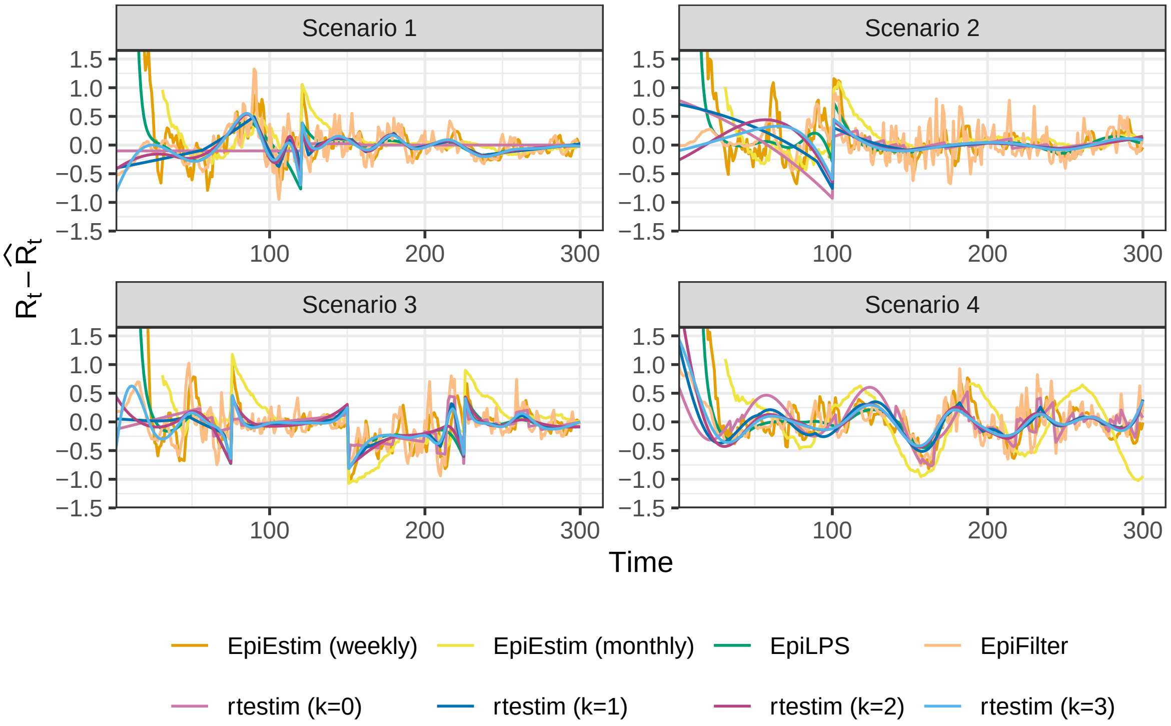

Figure A.7.6: Difference between the true and estimated instantaneous reproduction numbers for measles epidemics with negative binomial observations. Y-axes beyond 1.5 are truncated for a better illustration of small values.

#### A.8 Application of `rtestim` and all competitors on real epidemics

We apply all methods on Covid19 incidence in Canada, and the estimates are displayed in A.8.1. An alternative display which plots all estimated curves in one panel for an easier comparison is provided in A.8.2. All methods provide similar $\widehat{\mathcal{R}}_t$ curves beyond the early stage. Many methods, including `rtestim` ($k = 1, 2$), `EpiLPS`, and `EpiEstim` (weekly sliding window), all have large estimates (larger than 3) at the early stage of the epidemic. `EpiFilter` is much more wiggly than other estimates. All methods agree that the instantaneous reproduction number of Covid19 in Canada decreases to below 1 near June 2021 and reaches a small peak afterwards, and then decreases slowly until an outbreak at the end of 2021.

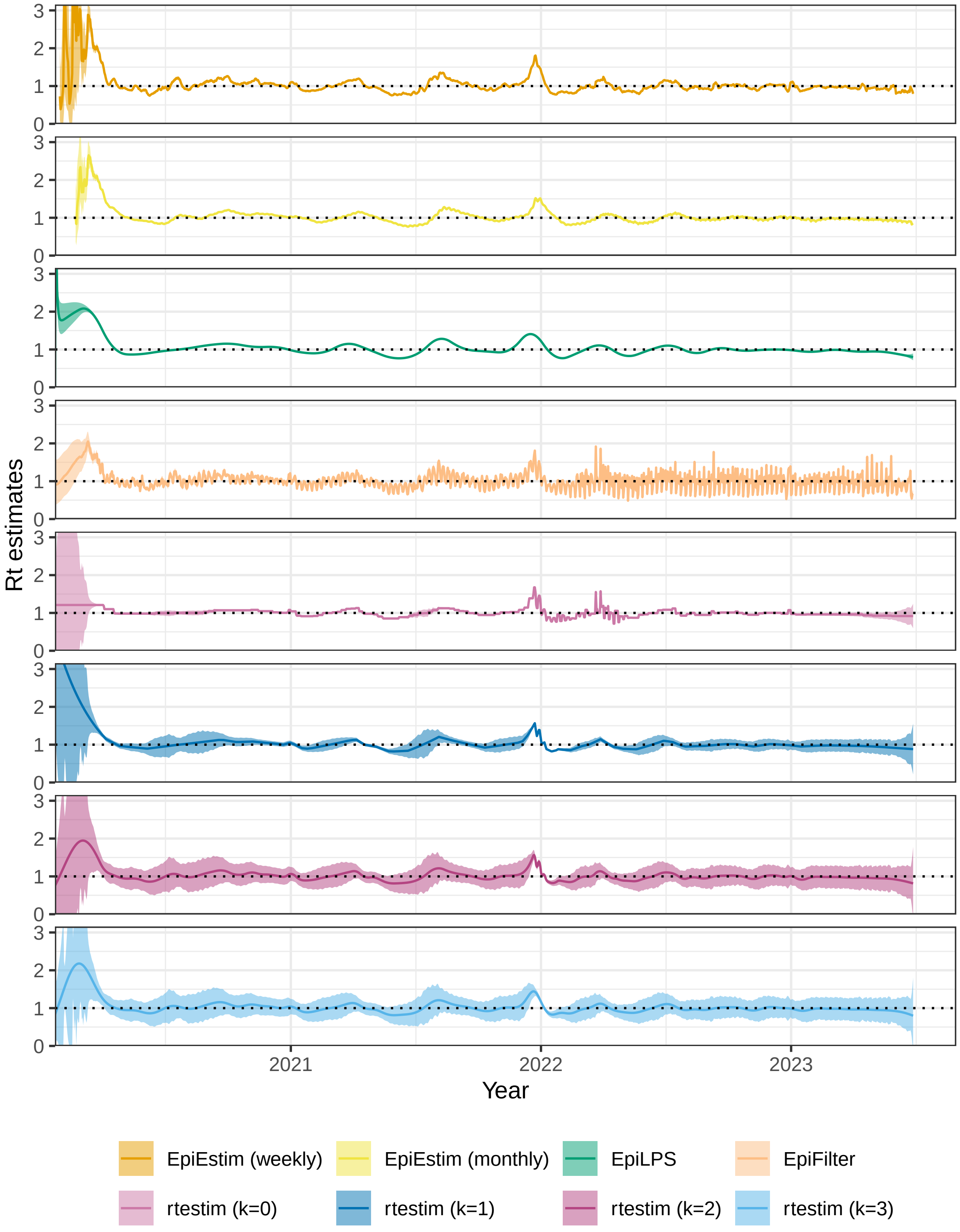

Figure A.8.1: Rt estimates with CIs for Covid19. Y-axes are truncated beyond 3 for a better display of the fluctuation in small values.

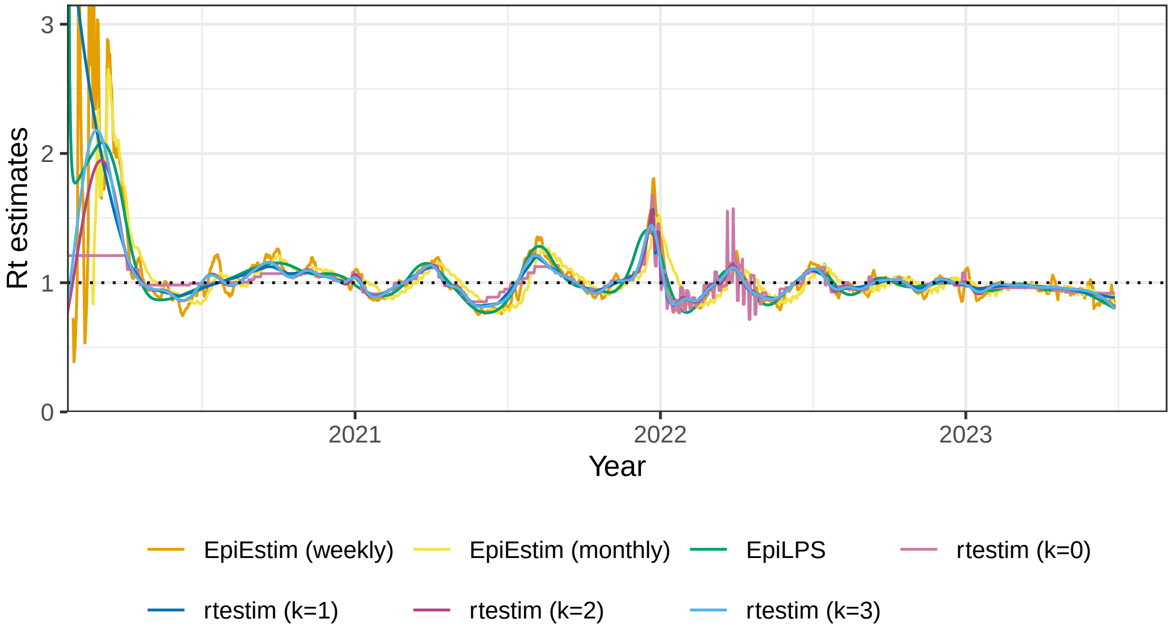

Figure A.8.2: Rt estimates for Covid19. Y-axis beyond 3 is truncated for a better display of the fluctuation in small values. EpiFilter is excluded here, because its estimates are too wiggly and make the plot less readable.

We also apply all methods on Flu in 1918. The results are visualized in Figures A.8.3 and A.8.4. `EpiEstim` with weekly sliding windows, `EpiFilter` and `rtestim` ($k = 0$) capture the peak of $\mathcal{R}_t$ (close to 3) at around day 30 since the start of the epidemic. While `EpiEstim` with monthly sliding windows, `EpiLPS`, `rtestim` ($k = 2, 3$) captures the increase around day 30, but have smaller estimates otherwise. Most methods agree that after day 50, the instantaneous reproduction number decreases to, and remains below, 1.

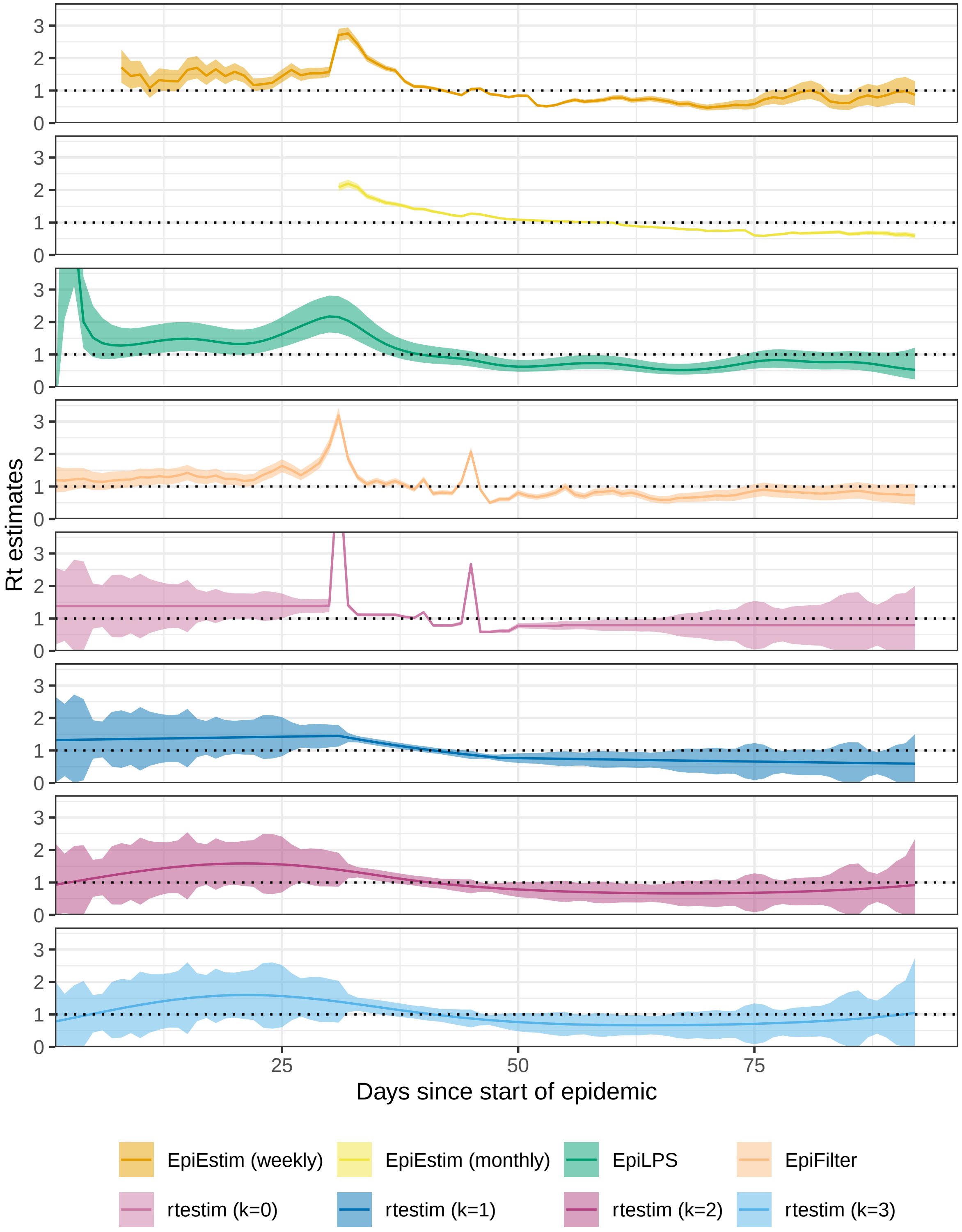

Figure A.8.3: Rt estimates with CIs for Flu 1918. Y-axes are truncated beyond 3.5 for a better display of the fluctuation in small values.

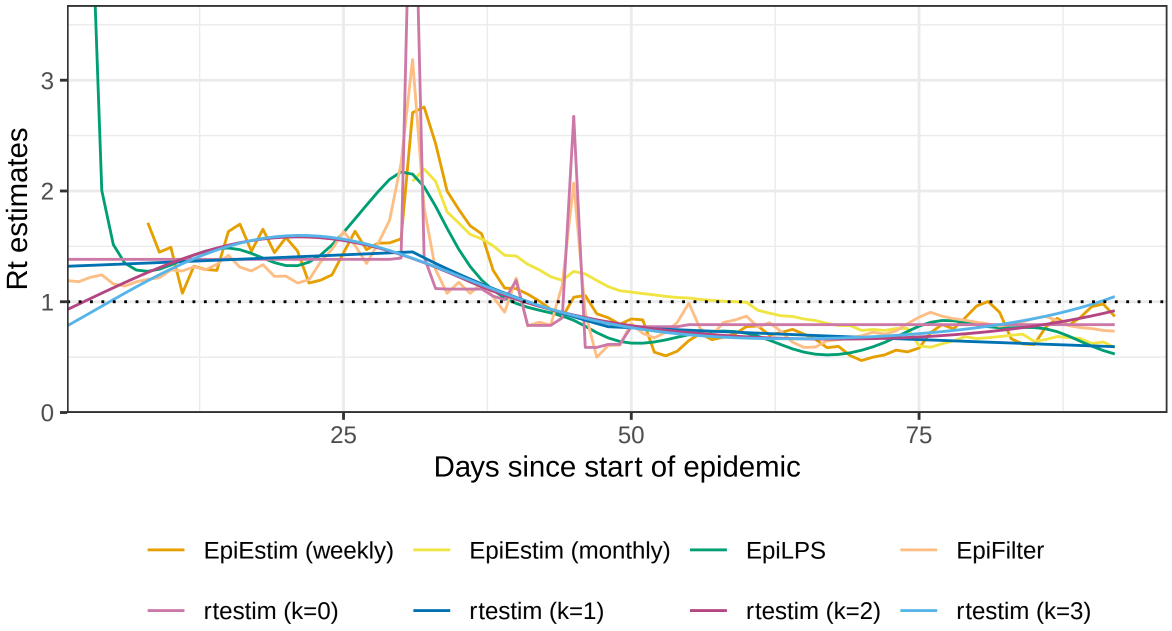

Figure A.8.4: Rt estimates for Flu 1918. Y-axis beyond 3.5 is truncated for a better display of the fluctuation in small values.

### References

Boëlle, Pierre-Yves, Severine Ansart, Anne Cori, and Alain-Jacques Valleron. 2011. "Transmission Parameters of the A/H1N1 (2009) Influenza Virus Pandemic: A Review." *Influenza and Other Respiratory Viruses* 5 (5): 306–16.

Bracher, Johannes, Evan L. Ray, Tilmann Gneiting, and Nicholas G. Reich. 2021. "Evaluating Epidemic Forecasts in an Interval Format." Edited by Virginia E. Pitzer. *PLoS Computational Biology* 17 (2): e1008618. https://doi.org/10.1371/journal.pcbi.1008618.

Cori, Anne, Neil M Ferguson, Christophe Fraser, and Simon Cauchemez. 2013. "A New Framework and Software to Estimate Time-Varying Reproduction Numbers During Epidemics." *American Journal of Epidemiology* 178 (9): 1505–12.

Ferguson, Neil M, Derek AT Cummings, Simon Cauchemez, Christophe Fraser, Steven Riley, Aronrag Meeyai, Sopon Iamsirithaworn, and Donald S Burke. 2005. "Strategies for Containing an Emerging Influenza Pandemic in Southeast Asia." *Nature* 437 (7056): 209–14.

Groendyke, Chris, David Welch, and David R Hunter. 2011. "Bayesian Inference for Contact Networks Given Epidemic Data." *Scandinavian Journal of Statistics* 38 (3): 600–616.

Lipsitch, Marc, Ted Cohen, Ben Cooper, James M Robins, Stefan Ma, Lyn James, Gowri Gopalakrishna, et al. 2003. "Transmission Dynamics and Control of Severe Acute Respiratory Syndrome." *Science* 300 (5627): 1966–70.

Wainwright, Martin J., and Michael I. Jordan. 2008. "Graphical Models, Exponential Families, and Variational Inference." *Foundations and Trends in Machine Learning* 1 (1–2): 1–305. https://doi.org/10.1561/2200000001.
